# Large-scale proteome and metabolome analysis of CSF implicates altered glucose metabolism and succinylcarnitine in Alzheimer’s disease

**DOI:** 10.1101/2021.09.02.21262642

**Authors:** Daniel J. Panyard, Justin McKetney, Yuetiva K. Deming, Autumn R. Morrow, Gilda E. Ennis, Erin M. Jonaitis, Carol A. Van Hulle, Chengran Yang, Yun Ju Sung, Muhammad Ali, Gwendlyn Kollmorgen, Ivonne Suridjan, Anna Bayfield, Barbara B. Bendlin, Henrik Zetterberg, Kaj Blennow, Carlos Cruchaga, Cynthia M. Carlsson, Sterling C. Johnson, Sanjay Asthana, Joshua J. Coon, Corinne D. Engelman

**Affiliations:** Department of Population Health Sciences, University of Wisconsin-Madison; 610 Walnut Street, 707 WARF Building, Madison, WI 53726, United States of America; National Center for Quantitative Biology of Complex Systems, University of Wisconsin-Madison; Madison, WI 53706, United States of America; Department of Biomolecular Chemistry, University of Wisconsin-Madison; Madison, WI 53506, United States of America; Wisconsin Alzheimer’s Disease Research Center, University of Wisconsin-Madison; 600 Highland Avenue, J5/1 Mezzanine, Madison, WI 53792, United States of America; Department of Medicine, University of Wisconsin-Madison; 1685 Highland Avenue, 5158 Medical Foundation Centennial Building, Madison, WI 53705, United States of America; Wisconsin Alzheimer’s Institute, University of Wisconsin-Madison; 610 Walnut Street, 9th Floor, Madison, WI 53726, United States of America; Department of Psychiatry, Washington University School of Medicine; St Louis, MO 63110, United States of America; NeuroGenomics and Informatics Center, Washington University School of Medicine; St Louis, MO 63110, United States of America; Hope Center for Neurological Disorders, Washington University School of Medicine; St Louis, MO 63110, United States of America; Roche Diagnostics GmbH; Penzberg, Germany; Roche Diagnostics International Ltd; Rotkreuz, Switzerland; William S. Middleton Memorial Veterans Hospital; 2500 Overlook Terrace, Madison, WI 53705, United States of America; Department of Psychiatry and Neurochemistry, Institute of Neuroscience and Physiology, the Sahlgrenska Academy at the University of Gothenburg; Mölndal, Sweden; Clinical Neurochemistry Laboratory, Sahlgrenska University Hospital Mölndal, Sweden; Department of Neurodegenerative Disease, UCL Institute of Neurology; London, UK; UK Dementia Research Institute at UCL; London, UK; Hong Kong Center for Neurodegenerative Diseases; Hong Kong, China; Morgridge Institute for Research; Madison, WI 53706, United States of America; Department of Chemistry, University of Wisconsin-Madison; Madison, WI 53506, United States of America

## Abstract

A major hallmark of Alzheimer’s disease (AD) is the aggregation of proteins (β-amyloid (A) and hyperphosphorylated tau (T)) in the brain, which makes the AD proteome in cerebrospinal fluid (CSF) of particular interest. Here, we conducted a CSF proteome-wide analysis among participants with and without AD pathology (n = 137 total participants: 56 A-T-, 39 A+T-, and 42 A+T+; 915 proteins analyzed), using a panel of 9 CSF biomarkers for neurodegeneration and neuroinflammation. We identified 61 proteins significantly associated with AT category (P < 5.46 x 10^-5^; strongest was SMOC1, P = 1.87 x 10^-12^) and 636 significant protein-biomarker associations (P < 6.07 x 10^-6^; strongest was a positive association between neurogranin and EPHA4, P = 2.42 x 10^-25^). Community network and pathway enrichment analyses highlighted three biomarker-associated protein networks centered around amyloid and tau measures, neurogranin, and the remaining biomarkers. Glucose metabolic pathways were enriched primarily among the amyloid- and tau-associated proteins, including malate dehydrogenase and aldolase A, both of which were associated with CSF phosphorylated tau levels in an independent replication cohort of 717 participants (P = 8.65 x 10^-56^ and P = 1.35 x 10^-45^). Follow-up interrogation of related CSF metabolite levels in the same samples as the discovery proteomics analysis identified increasing levels of succinylcarnitine with ptau and numerous other CSF biomarkers (P < 0.00056) that were replicated in an independent sample of 363 participants. Together, these results implicate glucose metabolic dysregulation and increased CSF succinylcarnitine levels as amyloid and tau pathology emerge in AD.

**One Sentence Summary:** Combining cerebrospinal fluid proteomics data with neurodegeneration and neuroinflammation biomarkers, genomics, and cerebrospinal fluid metabolomics, we identify and replicate a theme of altered glucose metabolism proteins and the metabolite succinylcarnitine across amyloid and tau progression in Alzheimer’s disease.

## INTRODUCTION

Despite much improvement in our understanding of it, Alzheimer’s disease (AD) continues to impose an enormous medical, social, and economic toll on society. An estimated 50 million people have dementia worldwide, with that number likely to increase to over 150 million by 2050(*1*). AD is the 6^th^ leading cause of death in the U.S. and costs an estimated $290 billion annually for healthcare(*2*). Part of the reason for this global impact of AD has been the lack of a cure or effective therapies for the disease, which is driven in part by an incomplete understanding of its causal mechanisms(*3*). The core pathological features of AD are well-described and center on the accumulation of two proteins, amyloid and tau, into amyloid plaques and neurofibrillary tangles(*4*), for which there are validated cerebrospinal fluid (CSF) biomarkers(*5*).

In order to better inform research on AD, there has been a shift in the conceptualization of the disease from a focus on clinical signs and symptoms(*6*) to AD biology measured *in vivo*. Using CSF assays related to amyloid deposition and hyperphosphorylation of tau protein (in addition to neuroimaging), it has become possible to leverage these biomarkers for identifying preclinical AD, mild cognitive impairment (MCI), and AD dementia(*7–10*). Most recently, in 2018, an explicit research framework for categorizing AD was proposed by the National Institute on Aging and Alzheimer’s Association (NIA-AA). This framework categorized individuals as amyloid positive (A+), tau positive (T+), and/or neurodegeneration positive (N+)(*11*). This so-called ATN framework—using ATN-based categorizations rather than more traditional clinical diagnoses as outcomes—provided nosological clarity in studying AD and other forms of dementia.

The use of these biomarker-defined categories is most relevant in multiomic approaches to studying AD pathophysiology, where molecular pathways are interrogated and clear case definitions are essential. Omics research offers immense promise for understanding complex disease by leveraging analyses of millions of molecular features spanning from the genome to the proteome, metabolome, phenome, and beyond(*12*). In the field of AD research, each of these individual omic approaches has already been applied extensively. Genomics research has highlighted a number of important loci, from the role of mutations in *APP*, *PSEN1*, and *PSEN2* in early-onset familial AD(*13*) to late-onset AD genetic risk factors like *APOE*, *CR1*, and *ABCA7*(*14–16*). CSF metabolomics studies have identified alterations in cholesterol, sphingolipid, norepinephrine, and other pathways(*17, 18*). In the CSF proteome, already known to include the amyloid and tau biomarkers for AD, studies have identified altered proteins related to the immune system and inflammation, carbohydrate metabolism, phospholipids, and the regulation of synapses(*19–24*).

Here, we combined the A and T of the ATN framework of AD with a novel CSF proteomics data set comprising 915 proteins generated for 137 participants, building on our recently published pilot study results in an independent sample(*25*). We comprehensively profiled the AD CSF proteome, its relationship to AT category, and its association with a diverse set of 9 AD CSF biomarkers covering measures of amyloid, tau, neurodegeneration, and neuroinflammation. These results were then extensively interrogated for pathway-level and network-based patterns, with top findings replicated in an independent AD proteomics cohort with an alternative proteomics modality and previously published AD proteomics associations with a similar mass spectrometry-based modality. The top-implicated biological pathway was then further explored with a focused metabolomics analysis using the same original participants and an independent metabolomics replication cohort of 363 participants. Finally, we combined the proteomics data set with previously generated genome-wide genotypes, 390 CSF metabolites, and demographic information to examine the relative utility of different omics data sets in predicting the AT-based categories. Elucidating the pathophysiology leading to the development of AD pathology and symptoms of AD dementia is expected to inform the identification of novel, effective drug targets.

## RESULTS

### Sample summary

CSF samples from 137 WRAP and ADRC participants were selected as described in the Methods, roughly evenly distributed across the three AT categories of interest (Table 1, Supplementary Figure 1, Supplementary Figure 2). Most (102, 74.5%) of the participants were cognitively unimpaired at the time of the sample, with 16 (11.7%) and 19 (13.9%) participants having an MCI or AD dementia diagnosis, respectively. The age and sex distributions across the AT categories varied, with worse AT pathology having a higher average participant age and a greater proportion of males. The amyloid and tau measures reflected the AT categorizations as expected. The remaining CSF biomarkers showed a general increase with increasing AT pathology with the exception of IL-6, which fluctuated across the groups.

**Table 1.**
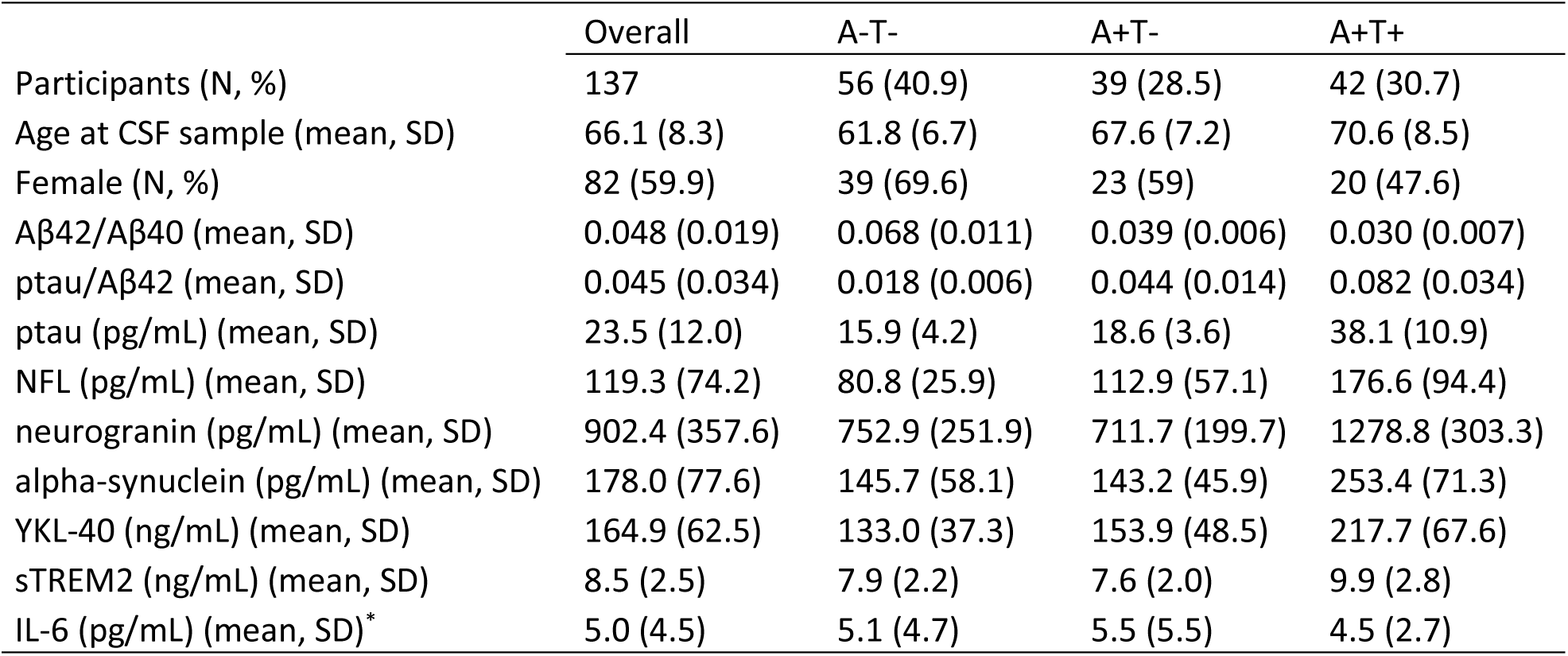
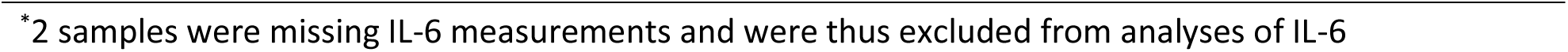
Summary of sample demographics and CSF biomarkers

|  | Overall | A-T- | A+T- | A+T+ |
| --- | --- | --- | --- | --- |
| Participants (N, %) | 137 | 56 (40.9) | 39 (28.5) | 42 (30.7) |
| Age at CSF sample (mean, SD) | 66.1 (8.3) | 61.8 (6.7) | 67.6 (7.2) | 70.6 (8.5) |
| Female (N, %) | 82 (59.9) | 39 (69.6) | 23 (59) | 20 (47.6) |
| Aβ42/Aβ40 (mean, SD) | 0.048 (0.019) | 0.068 (0.011) | 0.039 (0.006) | 0.030 (0.007) |
| ptau/Aβ42 (mean, SD) | 0.045 (0.034) | 0.018 (0.006) | 0.044 (0.014) | 0.082 (0.034) |
| ptau (pg/mL) (mean, SD) | 23.5 (12.0) | 15.9 (4.2) | 18.6 (3.6) | 38.1 (10.9) |
| NFL (pg/mL) (mean, SD) | 119.3 (74.2) | 80.8 (25.9) | 112.9 (57.1) | 176.6 (94.4) |
| neurogranin (pg/mL) (mean, SD) | 902.4 (357.6) | 752.9 (251.9) | 711.7 (199.7) | 1278.8 (303.3) |
| alpha-synuclein (pg/mL) (mean, SD) | 178.0 (77.6) | 145.7 (58.1) | 143.2 (45.9) | 253.4 (71.3) |
| YKL-40 (ng/mL) (mean, SD) | 164.9 (62.5) | 133.0 (37.3) | 153.9 (48.5) | 217.7 (67.6) |
| sTREM2 (ng/mL) (mean, SD) | 8.5 (2.5) | 7.9 (2.2) | 7.6 (2.0) | 9.9 (2.8) |
| IL-6 (pg/mL) (mean, SD)* | 5.0 (4.5) | 5.1 (4.7) | 5.5 (5.5) | 4.5 (2.7) |

\*2 samples were missing IL-6 measurements and were thus excluded from analyses of IL-6

### CSF proteomics descriptive analyses

The nLC-MS/MS analysis, MaxQuant identification, and LFQ quantification generated a total of 2,040 protein groups across the participants. After the proteomics quality control steps (Supplementary Table 1, Supplementary Figure 3, Supplementary Figure 4, Supplementary Figure 5), 915 proteins remained (Supplementary Table 2). Included in these proteins were YKL-40 (correlation with immunoassay measurement = 0.352, P = 2.40 x 10^-5^), sTREM2 (correlation with immunoassay measurement of sTREM2 = 0.490, P = 1.26 x 10^-9^), apolipoprotein E (APOE), amyloid precursor protein (APP; correlation with Aβ42 = 0.136, P = 0.114), amyloid-like protein-1 (APLP1), and APLP2. The tau protein was not reliably quantified by nLC-MS/MS in our samples. Little difference in protein missingness was seen by AT category (Supplementary Figure 3, Supplementary Figure 4, Supplementary Figure 5). The CSF proteome showed a rich correlation structure with both larger clusters and smaller pockets of highly correlated proteins (Figure 1a, Supplementary Table 3). Further interrogation with PCA underscored this complexity, with the first 4 PCs collectively explaining only half (49.89%) of the total variance (Supplementary Figure 6), with the top 2 PCs not explained by either AT or sex (Supplementary Figure 7). The top PC (PC1) was weakly correlated with age at sample (correlation = 0.17; P = 0.045), and its top 5 protein contributors (SEZ6L2, NFASC, L1CAM, PCDH1, and NRCAM) shared a theme of neuronal cell structure and adhesion. The second PC (PC2) was not correlated with age (correlation = -0.019; P = 0.82), and its top 5 protein contributors (C4orf48, DAF [CD55], MEGF10, FBLN3, and CNTFR) shared a theme of neuropeptides and glial function.

**Figure 1:**
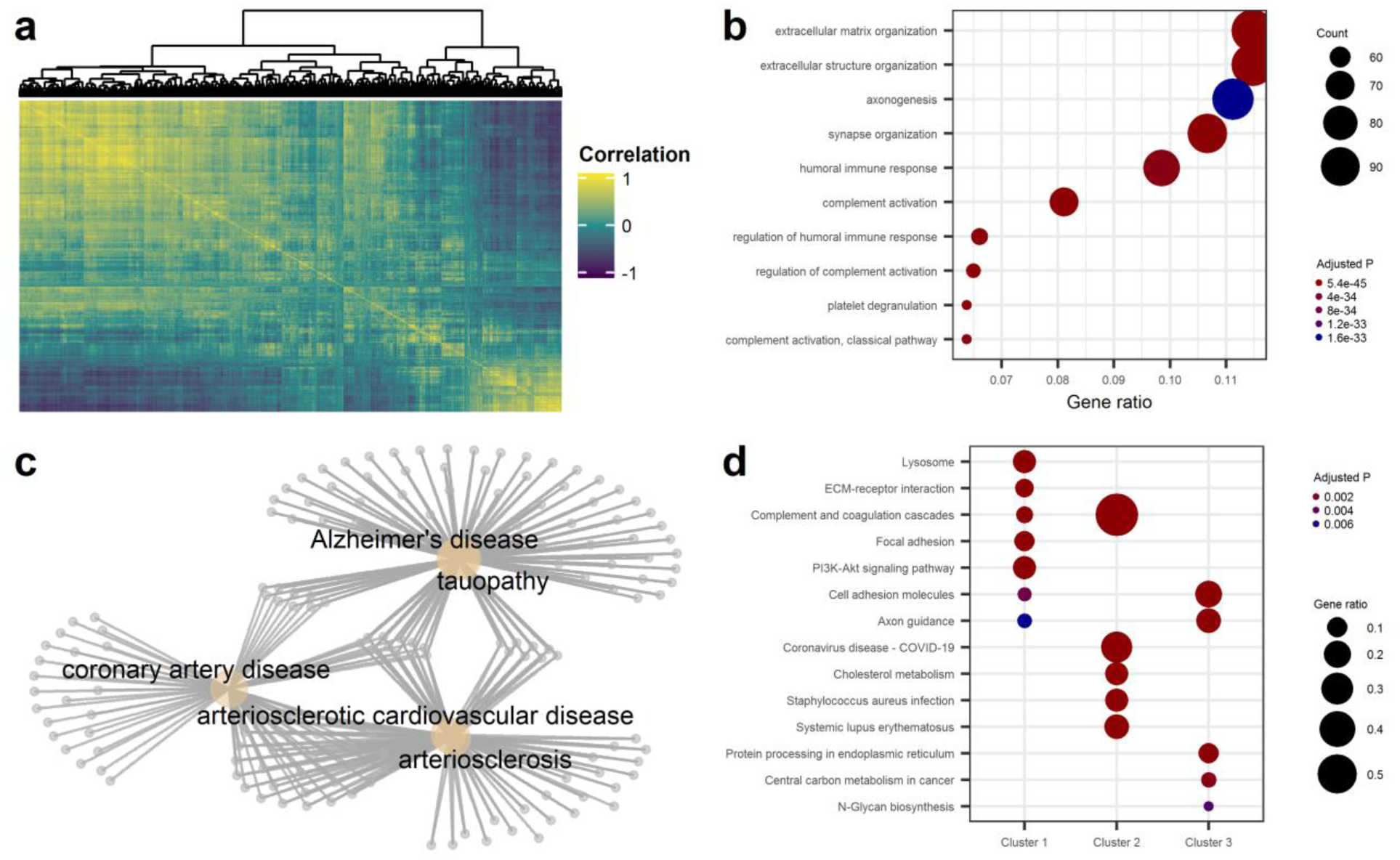
CSF proteomics descriptive analyses **a)** A correlation heatmap of the CSF proteins (n = 915) across all participants (n = 137) is shown. The dendrogram above the heatmap shows the results of hierarchical clustering of the proteins. An intricate set of correlation patterns can be seen, with both large clusters of proteins (e.g., top-left and bottom-right of the plot) and small local clusters seen throughout. **b)** An enrichment plot of the GO biological process terms among quantified proteins in the CSF compared to the entire human genome is shown. Significantly enriched processes included extracellular processes, processes involving axons and synapses, and immune system processes. **c)** A network representation of enriched DO terms among the CSF proteome is shown. The tan nodes represent significantly enriched disease terms, and the gray nodes represent proteins whose genes were associated with those terms. Three clusters of terms emerged: Alzheimer’s disease and tauopathy; coronary artery disease; and arteriosclerotic cardiovascular disease and arteriosclerosis. **d)** A cluster comparison plot of KEGG pathways between the three main clusters of CSF proteins is shown. The clusters were generated by Gaussian mixture modelling. Cluster one shows an enrichment of extracellular matrix and cell cycle pathways; cluster two shows enrichment of immune system-related pathways; and cluster three shows enrichment of a handful of other pathways.

Pathway enrichment analysis comparing the proteins quantified in the CSF to the entire human proteome revealed significant enrichment of terms related to extracellular, neuronal, immune system, and platelet pathways (Figure 1b, Supplementary Table 4). Significantly enriched DO pathways among the cohort included three groups of proteins with some small overlap: Alzheimer’s disease and tauopathy; coronary artery disease; and arteriosclerotic cardiovascular disease and arteriosclerosis (Figure 1c).

Given the apparent presence of clusters of proteins based on the correlation structure, the CSF proteome was divided into 3 clusters based on a Gaussian mixture model (Supplementary Figure 8). These three clusters were then compared to each other for the differential enrichment of biological pathways (Supplementary Table 5). The KEGG terms revealed a pattern where the smallest cluster (2) was enriched for immune system and cholesterol pathways while the other two larger clusters were enriched for extracellular and metabolism-related pathways (Figure 1d).

### Protein-AT associations

The ANCOVA tests revealed 61 statistically significant associations between proteins and AT category after multiple testing correction (P < 5.46 x 10^-5^), with a total of 496 (54.2%) of the proteins nominally associated (P < 0.05) (Supplementary Table 6). The differences in distribution of the top ten proteins revealed a number of different patterns in relation to amyloid and tau pathology (Figure 2a). Based on both the box plots and logistic regressions, some proteins increased (FABP3, SMOC1) consistently as AT pathology increased. Other proteins did not change from A- to A+ but did change from T- to T+ (GOT1, ALDOA, GDA, DDAH1, CRYM, and ACVR1B). Overall, there was an enrichment of statistical signal across the proteome that was not seen in the permutation sensitivity analysis (Figure 2b). Controlling for the *APOE* ε4 allele count did not substantially change the results, with 53 of the 61 proteins remaining significantly associated when the *APOE* variable was added to the ANCOVA models (Supplementary Figure 9). Among the set of significantly associated proteins, a number of biological pathways were enriched relative to the full human proteome (Supplementary Table 7), including extracellular matrix, secretory granule, and vesicle lumen GO cellular component terms; peptidase regulation GO molecular function terms; and glucose metabolic KEGG pathways (Figure 2c).

**Figure 2:**
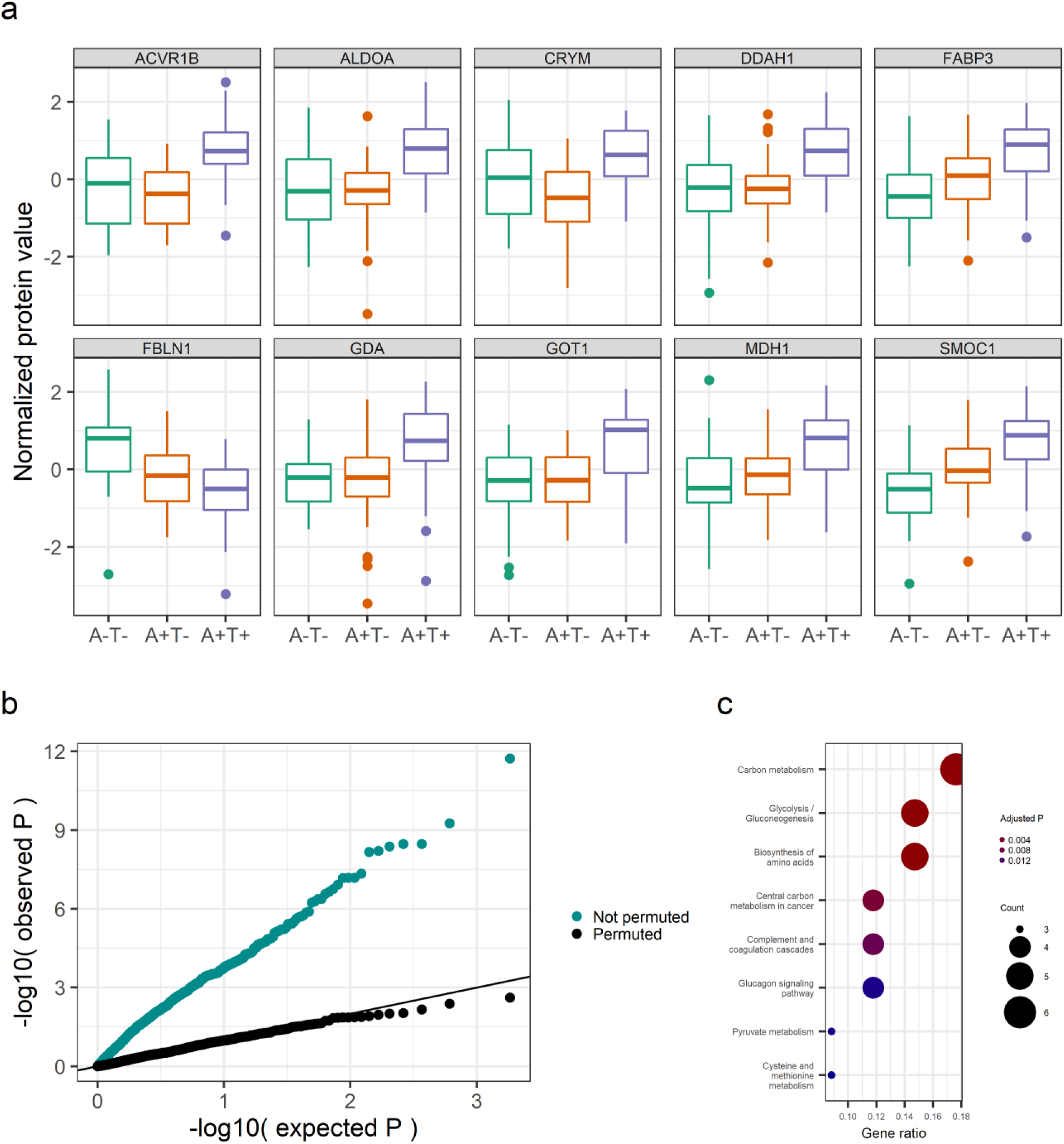
Proteins associated with **a)** The distributions of the top 10 most significantly associated proteins with AT category are shown. A number of different patterns were seen across increasing AT pathology, including proteins that increased consistently, decreased consistently, or increased only with tau positivity. **b)** The quantile-quantile (Q-Q) plots of the protein-AT ANCOVA association tests are shown above, with the distribution of P values shown separately for the original (“Not permuted”) and permuted data sets. Substantial signal enrichment was seen across the CSF proteome, with that signal absent in the permuted data set. **c)** The enriched KEGG pathways among the AT-associated proteins are shown, revealing a general perturbation of metabolic pathways.

When the logistic regression model was used to test for the direction of effect between proteins and A+T+ (vs. A-T-), only 9 proteins were significantly associated with being A+T+, and all of these proteins were also significantly associated in the ANCOVA model (Supplementary Figure 10). All but 1 (FBLN1) of the proteins significantly associated with A+T+ were increased in A+T+ relative to A-T- (Supplementary Figure 11).

### Protein-CSF biomarker associations

When each of the 9 CSF biomarkers (Aβ42/Aβ40, ptau, ptau/Aβ42, NFL, alpha-synuclein, neurogranin, YKL-40, sTREM2, and IL-6) was regressed on each CSF protein, a total of 636 protein-biomarker associations were statistically significant after Bonferroni correction (P < 6.07 x 10^-6^; Supplementary Table 8). As with the protein-AT associations, there was widespread association signal across the proteome with the CSF biomarkers that was not seen in the permutation test, except for IL-6, which had no significantly associated proteins (Supplementary Figure 12). The top 3 significantly associated proteins per biomarker are summarized in Table 2, with the full list of all statistically significant protein associations available in Supplementary Table 8. A total of 119 significantly enriched pathways among the biomarker-specific sets of significantly associated proteins was observed, with glucose metabolic pathways noted to be enriched among amyloid-related biomarkers (Supplementary Table 9).

**Table 2.** Top 3 significantly associated proteins for each biomarker

| Biomarker | UniProt ID | Entrez ID | Protein | Effect size | Effect size<br>(std. biomarker) | P value |
| --- | --- | --- | --- | --- | --- | --- |
| AB42/AB40 | Q9H4F8 | 64093 | SMOC1 | -0.010 | -0.54 | 4.55E-14 |
| AB42/AB40 | P40925 | 4190 | MDH1 | -0.009 | -0.46 | 1.44E-09 |
| AB42/AB40 | S4R371 | 2170 | FABP3 | -0.008 | -0.43 | 2.69E-08 |
| alpha-synuclein | Q16270 | 3490 | IGFBP7 | -50.56 | -0.65 | 1.04E-18 |
| alpha-synuclein | E7EUF1 | 5168 | ENPP2 | -49.04 | -0.63 | 1.96E-15 |
| alpha-synuclein | P12259 | 2153 | F5 | -45.88 | -0.59 | 6.30E-15 |
| log<sub>10</sub> NFL | Q16270 | 3490 | IGFBP7 | -0.092 | -0.43 | 4.28E-13 |
| log<sub>10</sub> NFL | P23142-4 | 2192 | FBLN1 | -0.091 | -0.43 | 5.55E-12 |
| log<sub>10</sub> NFL | P12259 | 2153 | F5 | -0.084 | -0.39 | 5.11E-11 |
| neurogranin | E9PG71 | 2043 | EPHA4 | 266.13 | 0.74 | 2.42E-25 |
| neurogranin | O00451 | 2675 | GFRA2 | 266.78 | 0.75 | 1.12E-23 |
| neurogranin | Q8TAG5-2 | 222008 | VSTM2A | 259.95 | 0.73 | 8.53E-23 |
| ptau | A0A087WZS0 | 66004 | LYNX1 | 6.70 | 0.56 | 1.05E-14 |
| ptau | P40925 | 4190 | MDH1 | 6.78 | 0.57 | 3.29E-14 |
| ptau | E9PG71 | 2043 | EPHA4 | 6.62 | 0.55 | 3.81E-14 |
| ptau/AB42 | Q9H4F8 | 64093 | SMOC1 | 0.019 | 0.57 | 1.83E-15 |
| ptau/AB42 | S4R371 | 2170 | FABP3 | 0.018 | 0.53 | 1.68E-12 |
| ptau/AB42 | P18669 | 5223 | PGAM1 | 0.017 | 0.50 | 4.72E-11 |
| sTREM2 | Q16270 | 3490 | IGFBP7 | -1.68 | -0.67 | 3.67E-19 |
| sTREM2 | E7EUF1 | 5168 | ENPP2 | -1.68 | -0.67 | 5.54E-17 |
| sTREM2 | P09486 | 6678 | SPARC | -1.47 | -0.59 | 3.01E-13 |
| YKL-40 | Q16270 | 3490 | IGFBP7 | -28.22 | -0.45 | 4.54E-12 |
| YKL-40 | E9PG71 | 2043 | EPHA4 | 24.60 | 0.39 | 3.45E-09 |
| YKL-40 | P12259 | 2153 | F5 | -24.43 | -0.39 | 4.63E-09 |

The network plot and subsequent community analysis of the protein-biomarker associations revealed three communities (modularity = 0.256) among the network (Figure 3). One community largely comprised the more traditional AD biomarkers of ptau, ptau/Aβ42, and Aβ42/Aβ40; a second community centered around the proteins associated with neurogranin; and the third community included the remaining biomarkers of alpha-synuclein, YKL-40, NFL, and sTREM2 (IL-6 had no significant protein associations and was not included). The largest number of shared associations across the biomarkers occurred among neurogranin, ptau, and alpha-synuclein, which shared 103 protein associations between at least two of those biomarkers (Supplementary Figure 13).

**Figure 3:**
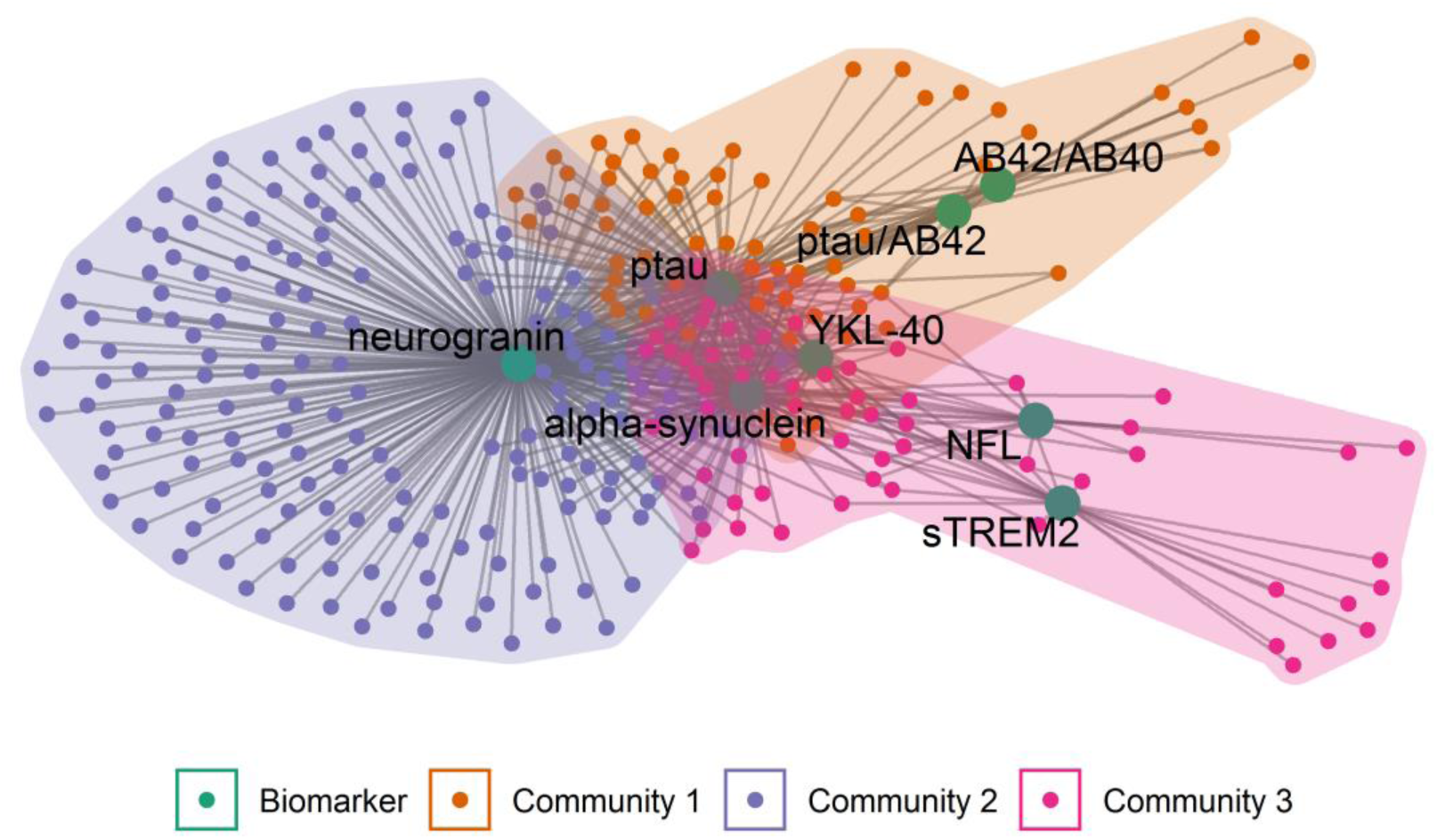
Network analysis of protein-biomarker associations A bipartite graph representation of the proteins significantly associated with the CSF biomarkers after Bonferroni correction is shown. The nodes representing the biomarkers are larger and in green. Proteins are represented as smaller nodes, with an edge to a biomarker representing a significant association between a protein and a biomarker. The colors of the protein nodes and underlying shaded regions correspond to three distinct communities identified with the fast greedy modularity optimization algorithm. Three such communities were identified. Community 1 included proteins associated with the more traditional AD biomarkers, such as ptau, ptau/Aβ42, and Aβ42/Aβ40. Community 2 centered around neurogranin, including many proteins uniquely associated with neurogranin. Community 3 included proteins associated with the remaining biomarkers of alpha-synuclein, YKL-40, NFL, and sTREM2. No proteins were significantly associated with IL-6. Many proteins were shared in the center of the network, particularly among neurogranin, ptau, and alpha-synuclein.

### Replication of top pathway results

The glucose metabolism pathway (REACTOME ID R-HSA-70326) was significantly enriched among all three of the amyloid and tau measures, so the results of the 9 proteins from this pathway that were significantly associated with one of the amyloid or tau biomarkers were chosen for replication in the Knight ADRC, which used an aptamer-based instead of an MS-based proteomics platform. These proteins included MDH1, ALDOA, PGK1, TPI1, PGAM1, PKM, GOT1, ALDOC, and ENO1. Of these 9 proteins, 5 of them (MDH1, ALDOA, PGK1, TPI1, PGAM1) were measured in the CSF in the Knight ADRC. For the protein associations with CSF amyloid levels, there was statistically significant (P < 0.0056) and directionally concordant replication of the associations of MDH1, ALDOA, and TPI1 with both levels of CSF ptau/Aβ42 and CSF ptau (Table 3). Associations of PGAM1 with CSF ptau/Aβ42 and CSF ptau were nominally significant in the replication analysis with P values closer to the replication significance threshold. For CSF amyloid levels, no signals were statistically significantly replicated, which might be in part due to a difference in amyloid outcome (Aβ42/40 vs. Aβ42). PGK1 was statistically significantly associated with both CSF Aβ42 and CSF ptau/Aβ42 levels, but in the opposite direction as observed, which may be related to technical differences in the underlying proteomics platforms for this protein. Looking at plasma levels of these same 9 proteins and their associations with these biomarkers, only 2 of the 9 proteins were analyzed in the Knight ADRC data set (TPI1 and PGAM1), but neither showed convincing evidence of association with the CSF biomarkers (Supplementary Table 10).

**Table 3.** Replication of implicated glucose metabolism-related proteins in the Knight ADRC

| Protein | Discovery (Wisconsin ADRC) |  |  |  |  |  | Replication (Knight ADRC) |  |  |  |  |  |
| --- | --- | --- | --- | --- | --- | --- | --- | --- | --- | --- | --- | --- |
|  | CSF AB42/40 |  | CSF ptau/AB42 |  | CSF ptau |  | CSF AB42 |  | CSF ptau/AB42 |  | CSF ptau |  |
|  | Beta | P value | Beta | P value | Beta | P value | Beta | P value | Beta | P value | Beta | P value |
| MDH1 | -0.0088 | 1.44E-09 | 0.016 | 3.21E-10 | 6.78 | 3.29E-14 | -4.53 | 7.61E-01 | 0.043 | 4.13E-22 | 19.13 | 8.65E-56 |
| ALDOA | -0.0070 | 1.75E-06 | 0.014 | 7.34E-08 | 6.59 | 1.26E-13 | -3.73 | 8.02E-01 | 0.040 | 3.75E-19 | 17.50 | 1.35E-45 |
| PGK1 | -0.0068 | 5.67E-06 | 0.013 | 5.08E-07 | 4.13 | 2.05E-05 | 82.96 | 1.49E-08 | -0.016 | 5.21E-04 | -1.12 | 4.04E-01 |
| TPI1 | -0.0071 | 9.01E-07 | 0.012 | 4.51E-06 | 4.43 | 1.71E-06 | 22.07 | 1.36E-01 | 0.032 | 1.86E-12 | 15.80 | 1.78E-36 |
| PGAM1 | -0.0081 | 4.35E-08 | 0.017 | 4.72E-11 | 5.38 | 1.16E-08 | -39.12 | 8.06E-03 | 0.014 | 3.32E-03 | 3.57 | 7.90E-03 |
| ALDOC | -0.0048 | 1.44E-03 | 0.008 | 2.25E-03 | 4.42 | 2.53E-06 | . | . | . | . | . | . |
| ENO1 | -0.0068 | 4.72E-06 | 0.011 | 3.86E-05 | 2.86 | 3.12E-03 | . | . | . | . | . | . |
| GOT1 | -0.0072 | 5.67E-07 | 0.014 | 2.25E-08 | 6.58 | 5.88E-14 | . | . | . | . | . | . |
| PKM | -0.0066 | 6.93E-06 | 0.014 | 5.75E-08 | 6.65 | 5.14E-14 | . | . | . | . | . | . |

A "." indicates a protein that was not present in the QC'd Knight ADRC data set.

When checked for replication in the CSF proteomics results of Higginbotham et al. 2020 (see their Table S2A(*26*)), 8 of these glucose metabolism-related proteins (MDH1, ALDOA, PGK1, TPI1, PGAM1, ALDOC, ENO1, and GOT1) showed a nominally significant increase in AD participants relative to controls (P < 0.05), and all of these but TPI1 had an FDR < 0.05. In the brain proteomics results reported by Johnson et al. 2020 (see their Supplementary Table 2A(*24*)), 5 proteins (MDH1, PGK1, TPI1, ENO1, and PKM) showed at least a nominally statistically significant difference (P < 0.05) from the ANOVA analysis across controls, asymptomatic AD, and AD participants. Of these, TPI1 showed a statistically significant increase between asymptomatic AD and AD participants (P = 0.016) while the other 4 proteins (MDH1, PGK1, ENO1, and PKM) were statistically significantly increased in AD participants relative to controls (P range from 0.019 to 9.5 x 10^-5^).

### Secondary analysis of insulin-related proteins

Of the insulin-related proteins of interest that had not been already included in the QC’d data set, only IGF-1 and AKT1 were identified in the proteomics workflow. AKT1 was only quantified in one subject and was thus not suitable for further analysis, but IGF-1 was quantified in 82 samples (59.9%) and analyzed further using only the non-imputed measurements. A trend in missing values by AT category was noted: 44.6% of samples were missing IGF-1 in A-T-, 41.0% in A+T-, and 33.3% in A+T+. The ANCOVA analysis of IGF-1 did not show a statistically significant difference of the protein across AT categories (P = 0.170), though the distribution of the protein appeared to increase with amyloid positivity (Supplementary Figure 14). The association analysis between IGF-1 and the CSF biomarkers revealed a nominally significant negative association with Aβ42/Aβ40 (P = 0.011) and positive association with ptau/Aβ42 (P = 0.009) (Supplementary Table 11, Supplementary Figure 15).

### Secondary analysis of glycolysis and TCA cycle metabolites

Of the 10 glycolysis and TCA cycle-related metabolites examined in the discovery analysis (Supplementary Table 12), only one (succinylcarnitine (C4-DC)) was statistically significantly associated with AT category after multiple testing correction (P = 1.57 x 10^-6^;Supplementary Table 13; Supplementary Figure 16). This metabolite was also statistically significantly associated with CSF neurogranin, alpha-synuclein, ptau, sTREM2, YKL-50, NFL, and ptau/Aβ42, in each case indicating that succinylcarnitine increases in the CSF along with increasing levels of the biomarker (Supplementary Table 14). In the independent replication cohort (n = 363) (Supplementary Table 15), succinylcarnitine showed a similar trend to what was seen in the discovery cohort. This association was weaker than that seen in the discovery cohort (P = 0.014), though the replication cohort distribution across the AT categories was strongly skewed to include more A-T-participants than the other two categories. More convincing were the succinylcarnitine-biomarker associations (i.e., with neurogranin, alpha-synuclein, ptau, sTREM2, YKL-50, and NFL), which were replicated for all biomarkers except for ptau/Aβ42.

### Multiomic prediction models for amyloid and tau

The results of the multiomic amyloid and tau prediction models revealed a consistent pattern where the CSF proteome outperformed the other individual omic data sets in predicting positivity based on the core biomarkers of Aβ42/Aβ40, ptau/Aβ42, and ptau (Figure 4, Supplementary Table 16). The predictive model based on the CSF proteome (number of predictors selected ranged from 75-105) achieved a high AUC across for all three biomarkers (Aβ42/Aβ40 AUC = 0.839, ptau/Aβ42 AUC = 0.920, and ptau AUC = 0.954), performing slightly better than even the integrative model when predicting ptau or ptau/Aβ42. For Aβ42/Aβ40 and ptau positivity, the sensitivity and specificity values further demonstrated the relative superiority of the proteomics model. For Aβ42/Aβ40 positivity, the sensitivity of the CSF proteome model was 0.800 compared to much lower values (0.050–0.450) from the other models. For ptau positivity, the specificity of the CSF proteome model (0.800) was much higher than the other models (0.000–0.200). In all cases, the genome-based model performed poorly (AUCs ranged from 0.525–0.679) (Figure 4a). The 2D histograms showing the performance of the CSF proteome relative to the raw biomarker values highlighted the effective classification by the proteomic models with effective delineation between positive and negative amyloid statuses (Figure 4b–d).

**Figure 4:**
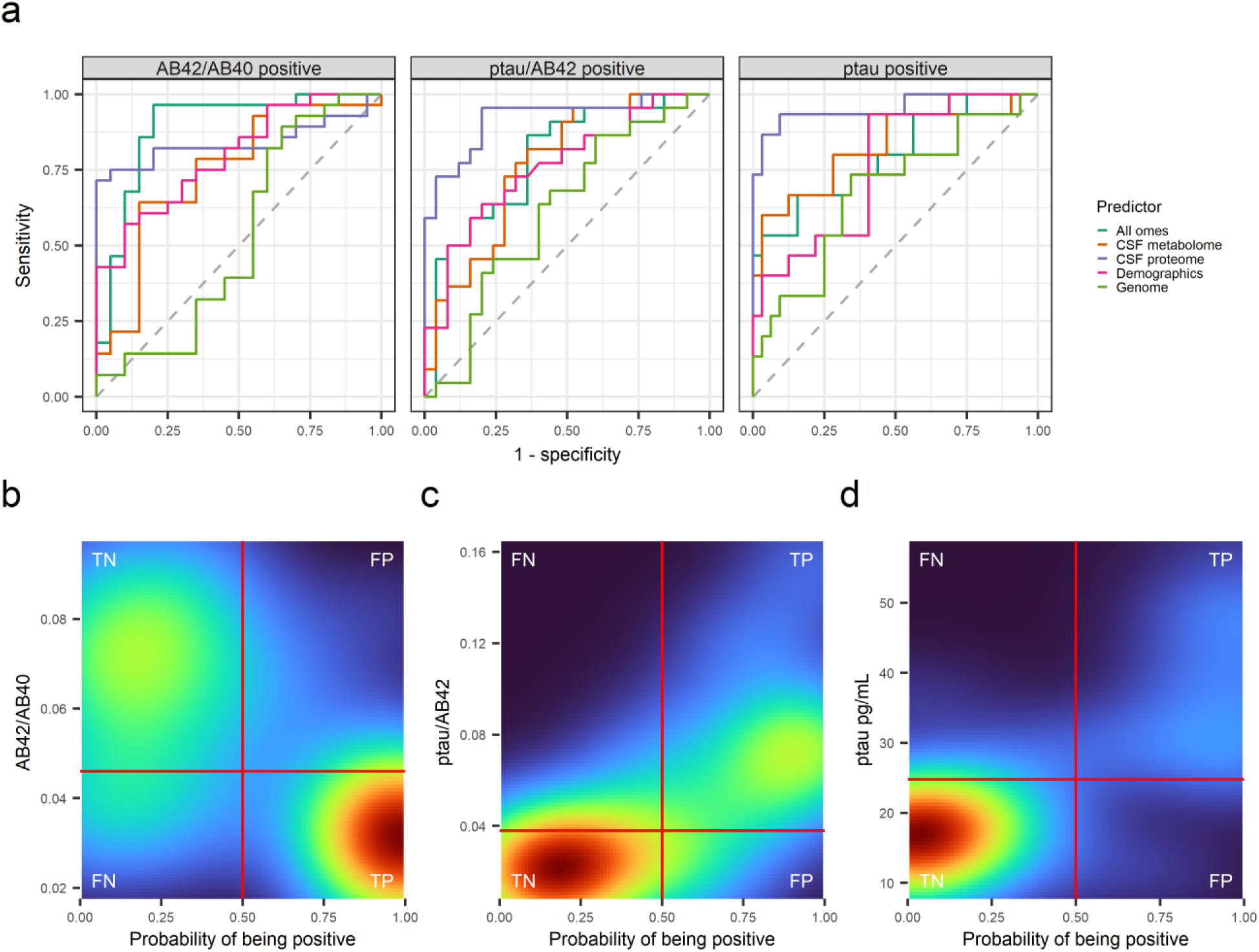
Multiomic predictive model performance for amyloid and tau The ability of different omic data sets to predict the key AD CSF biomarkers for amyloid and tau is summarized. **a)** The receiver operator characteristic (ROC) curves for Aβ42/Aβ40, ptau/Aβ42, and ptau are shown for the held-out testing data set predictions. Each omic prediction set is plotted with a different line. **b-d)** A 2D density plot summarizes the actual values and predicted probability of positivity for each CSF biomarker (Aβ42/Aβ40, ptau/Aβ42, and ptau, respectively) from the proteomic predictor models applied to the held-out testing data set. The text labels in the corners refer to the prediction categories: TN = true negative, FP = false positive, TP = true positive, and FN = false negative. The vertical red line indicates the threshold for a hard classification of biomarker positivity from the prediction models. The horizontal red line indicates the binary threshold for the CSF biomarker determined from previous work (see Methods).

## DISCUSSION

In this study, we completed four main CSF proteomics analyses with multiple and diverse sources of independent replication: 1) a characterization of the CSF proteome in an AD-focused cohort; 2) a differential proteomics analysis centered on the ATN framework rather than clinical diagnoses; 3) an interrogation of the individual and pathway-level associations of the proteome with a robust set of neurodegeneration and neuroinflammation biomarkers; and 4) a comparative multiomic predictive analysis of amyloid and tau using different omic data sets on the same set of participants. We found that the results of the proteomics discovery and replication analyses highlighted altered (namely increased) levels of glucose-metabolism-related proteins in the CSF. Further triangulation with CSF metabolomics data on the same participants from the discovery proteomics cohort and independent replication identified a positive association of succinylcarnitine with ptau and markers of neurodegeneration and neuroinflammation. These results provide new insight into the specifics and timing of glucose metabolic dysregulation in AD.

Using the pipeline we developed from the pilot study, we successfully quantified 915 proteins that passed our QC metrics across 137 participant CSF samples. Our participant population covered the spectrum of amyloid and tau positivity in a largely preclinical cohort that, in combination with the rich set of standard and novel CSF biomarkers of neuroinflammation and neurodegeneration, gave us a window into the proteomic changes occurring as amyloid and tau accumulate. Among the CSF proteins we quantified, significantly enriched functional annotation included extracellular matrix, axonogenesis, humoral immune system, complement system, and platelet pathways (Figure 1b), which was similar to previous work despite the difference in cohort (here, AD-focused, compared to typical or healthy CSF samples)(*27–32*). Also similar to previous work was our finding that there was no discernible difference by sex among the top 2 PCs of our CSF proteome (Supplementary Figure 7), echoing a previous study where unsupervised hierarchical clustering failed to distinguish samples by sex(*29*), though we note that such results do not preclude sex differences in individual AD-related proteins. When our CSF proteome was examined for significantly enriched disease-related proteins, we identified enriched clusters of proteins related to AD and tauopathy. This finding underscored results from the Zhang et al. study of 2,513 proteins from 14 CSF samples that showed an enrichment of proteins related to neurological disease in the CSF(*30*), though we also identified enriched clusters of proteins related to cardiovascular disease among the CSF proteome (Figure 1c), which could potentially reflect differences in the studied population.

We identified a total of 61 AT-associated CSF proteins after multiple testing correction (Supplementary Table 6), with 43 of these proteins having been previously implicated in AD. The protein here with the strongest association with AT category was the SMOC1 protein, which increased in the CSF with increasing pathology along the AT categories. The increase of SMOC1 with increasing pathology or disease severity has been noted before(*26, 33, 34*), and the protein has also been found to partly colocalize with amyloid plaques(*35*). Fatty acid binding protein 3 (FABP3), which was found to increase across AT categories in this study, is another example. FABP3 has been commonly associated with AD across CSF proteomics studies(*34, 36, 37*). However, several proteins commonly associated with AD were not significantly associated with AT category here, including APOE, clusterin, and secretogranin(*37*). Although the reason for this lack of association is unclear, it could be due to the present cohort being largely preclinical. On the other hand, we identified several novel protein associations, including ectonucleotide pyrophosphatase/phosphodiesterase family member 5 (ENPP5, noted to possibly be involved in neuronal cell communications(*38*)), heparin cofactor2 (SERPIND1, previously associated with multiple sclerosis(*39*)), extracellular matrix protein 2 (ECM2, jointly associated with iron along with APOE(*40*)), and glycoprotein endo-alpha-1,2-mannosidase-like protein (MANEAL, where variants in both *MANEAL* and *OSTM1* have been observed in connection with a neurodegenerative disorder(*41*)). The enriched KEGG pathways among the AT-associated proteins were carbon metabolism (hsa01200), biosynthesis of amino acids (hsa01230), and glycolysis/gluconeogenesis (hsa00010), and all three of these enriched pathways were enriched whether the full human proteome or only our 915 CSF proteins were used as the background distribution. The enriched GO terms varied but included multiple pathways related to the regulation of peptidases, some of which have been shown to be related to AD and amyloid metabolism(*42, 43*). The differential expression of proteins related to metabolism has been seen in other CSF proteomics cohorts(*24, 26, 36, 44*), but although protein peptidases are known to be present in a large number in the CSF proteome(*28*), there is comparatively less proteomics work highlighting the role of peptidases beyond a study by Whelan et al. that found up-regulated endopeptidases in AD patients relative to controls(*23*).

One of the major trends in AD research has been the movement toward a biomarker-based definition of AD(*11*). Recent CSF proteomics work in AD has explored the relationship between CSF protein levels and various AD biomarkers, especially measures of amyloid and tau pathology(*34, 45*). We replicated numerous previously reported protein associations with CSF levels of ptau (e.g., SMOC1, BASP1, and GAP43)(*34, 45*), but failed to replicate any of the proteins significantly associated with CSF amyloid, though we note that our analysis used Aβ42/Aβ40 as the outcome rather than Aβ42 since Aβ42/Aβ40 is considered to be a better biomarker for AD than Aβ42 alone(*46*).

Unique to this study was the inclusion of a more comprehensive set of CSF biomarkers relevant to AD, neurodegeneration, and neuroinflammation. Each biomarker had its own unique set of significantly associated proteins, but the network community analysis revealed that the affected proteins for the biomarkers tended to separate between the traditional AD biomarkers of amyloid and tau, neurogranin, and the remaining biomarkers. Notably, IL-6—a cytokine that has been explored as a possible marker of AD-related neuroinflammation—was alone in lacking statistically significant protein associations, supporting a meta-analysis that found no significant difference in peripheral IL-6 levels between AD cases and controls(*47*). Collectively, our results identify broadly different protein networks associated with the classical AD biomarkers compared to more general markers of neurodegeneration and neuroinflammation.

The pathway enrichment analysis of biomarker-associated proteins underscored the differences among these protein groups (Supplementary Table 9). Among the amyloid and tau biomarkers, the associated proteins shared a common theme of enrichment of glucose metabolism pathways, including two proteins (MDH1 and ALDOA) that showed evidence of association with AD diagnosis in the Knight ADRC replication data set and in one or both of the AD MS proteomics data sets from Higginbotham et al. 2020(*26*) or Johnson et al 2020(*24*). MDH1 levels have been associated with AD in the past, although the direction of effect has not been consistent(*48–50*). Here, MDH1 was observed to increase with amyloid and tau pathology. ALDOA has been previously positively associated with CSF tau levels(*34*), as was the case here, and also associated with the *APOE* ε4 allele(*51*). Glucose metabolism, which is the major source of energy for the brain, has long been known to show signs of dysfunction in AD even before the emergence of symptoms(*52–56*). Our findings here, where 74% of participants were cognitively unimpaired, support the emergence of glucose metabolic dysregulation presymptomatically as amyloid and tau begin to show alterations. Further underscoring potential abnormalities in energy metabolism as AD develops is the observation that 18 of the 61 proteins associated with AT here have been previously connected with insulin resistance, including pyruvate kinase (*PKM*), alpha-enolase (*ENO1*), and triosephosphate isomerase (*TPI*), which in a previous study were found to be elevated in participants with type 1 diabetes(*57*). These proteins were statistically significantly associated with one or more of the three amyloid and tau biomarkers. Importantly, the enrichment of glucose metabolic pathways among the amyloid and tau-related proteins in this study was not seen among the other two communities of biomarkers, which instead tended to be enriched for extracellular matrix, cell junctions and adhesion, and other pathways. Taken together, these results provide evidence for a dysregulation of the glucose metabolic proteome that is more specific to amyloid and tau than to biomarkers of neuroinflammation or neurodegeneration.

The secondary analysis of IGF-1 further suggests possible abnormalities in insulin signaling. IGF-1, which can bind insulin receptors(*58*), has been previously implicated in AD in studies that found decreased *IGF1* expression in brain tissue and evidence of IGF-1 resistance(*59–61*). Here, we found increased levels of CSF IGF-1 with increasing amyloid pathology, though the lower sample size in the IGF-1 analysis warrants caution in the interpretation. We also observed a statistically significant decrease in IGFBP7 levels from A-T- to A+T+ and a negative correlation with sTREM2, alpha-synuclein, neurogranin, ptau, NFL, and YKL-40. IGFBP7 can bind to IGF1, and the levels of IGFBP7 have been noted to increase in brain tissue in both mouse models of AD and AD patients(*62*). The difference in the observed direction of effect might be related to differences between the brain tissue and CSF, though it is worth further investigation.

The secondary analysis of specific metabolites in the glycolysis and TCA cycle pathways provided a greater depth of insight into the proteomic changes detected in the main analysis. Among the participants whose CSF samples were included in the proteomics analysis, only one of the 10 CSF metabolites showed a significant association with AT category—succinylcarnitine (C4-DC)—which increased in the A+T+ group relative to both the A-T- and A+T-groups. A similar trend was observed in the independent replication cohort, though with only nominal significance (P < 0.05;Supplementary Figure 16). Succinylcarnitine is a type of acylcarnitine, which typically play a role in metabolism by transporting fatty acids from the cytoplasm into the mitochondria for beta-oxidation(*63*).

Succinylcarnitine may also play a role in the transfer of succinyl groups between the mitochondria and the cytosol through the carnitine shuttle(*64*). With regard to the TCA cycle, succinyl-CoA (one of the TCA cycle intermediates) may be esterified into succinylcarnitine. In mouse models of aging and AD, succinylcarnitine levels were found to increase significantly in the brain hippocampus along with multiple other changes to metabolite levels from glycolysis and the TCA cycle(*65*). In another mouse model of aging, succinylcarnitine increased significantly in the hippocampi of older mice, implicating a diversion from succinate in the TCA cycle and broader issues with TCA cycle metabolism. Our results here provide evidence in humans that implicates succinylcarnitine in AD in conjunction with changing tau levels, complementing previous AD genomics findings relating a nearby enzyme (succinyl-CoA ligase, *SUCLG2*) with CSF amyloid levels(*66*). Moreover, recent AD metabolomics work in the blood and brain identified general acylcarnitine dysregulation and specifically found decreased levels of short-chain acylcarnitines(*67*) (the kind of acylcarnitine that succinylcarnitine is), which potentially indicates that either the CSF or succinylcarnitine is an exception to this general trend. More broadly, our proteomics and follow-up metabolomics analyses reveal a situation where the protein machinery of glucose metabolism is upregulated as amyloid levels change, while succinylcarnitine levels increase only when tau begins to change, potentially signaling problems with the metabolic pathways trying to keep up with the demands imposed by AD. Furthermore, given the known increases in acylcarnitines more generally in aging(*68, 69*), it is possible that increasing succinylcarnitine could be connected with oxidative stress in the brain as well. Additional joint multiomics work will be needed to pinpoint the timing and context of these changes in succinylcarnitine levels and their implications.

A potential consequence of altered glucose metabolism is a disruption in autophagy and proteostasis(*52*), which has been observed in AD(*70–73*). Although many proteasome-relevant proteins were not quantified in our CSF data set, several heat shock proteins, which often help regulate protein folding and degradation, were, including the chaperone protein HSP90AA1. We observed a statistically significant negative association of HSP90AA1 levels with Aβ42/Aβ40 and a significant positive association with ptau/Aβ42, consistent with worse pathology. Interestingly, cathepsin D (CTSD), a lysosomal protease that has been targeted by drugs seeking to modulate autophagy in AD mouse models(*73, 74*), was statistically significantly associated with sTREM2, alpha-synuclein, neurogranin, NFL, and YKL-40, but not with amyloid or tau (Supplementary Table 8). Moreover, we observed an enrichment of peptidase-related pathways among proteins associated with AT category, ptau, and neurogranin (Supplementary Table 7, Supplementary Table 9). Further potential evidence implicating proteostasis was the substantial enriched association signal across the proteome, with 54.2% (496/915) of CSF proteins being nominally associated with AT category and substantial deviation from the expected null distribution across the proteome (Figure 2b). These nominally associated proteins included many proteins known to form intracellular or extracellular deposits in disease(*71*), including APP, TTR, B2M, APOA1, APOA2, APOA4, APOC2, APOC3, LYZ, CST3, SOD1, IGHG1, IGHG2, IGHG3, and HBB (Supplementary Table 6). Although individual protein associations and pathway enrichment have tended to be the focus of previous work, several studies have reported similar widespread enrichment among the proteome, including a case-control study of AD diagnosis with 487 of 1,968 (24.7%) of proteins nominally associated with AD diagnosis(*75*) and a study of protein correlations with CSF total tau, ptau, and Aβ42 where 63 out of 106 proteins (59.4%) were associated with at least one of the biomarkers(*45*). Indeed, one core feature of AD proteomics work historically has been the identification of numerous AD-associated proteins that nonetheless do not replicate upon further study(*36, 37*). There are technical and study design reasons why such protein associations may not replicate easily(*36*), and unmeasured confounding could certainly explain some of the signal enrichment, but another potential reason could be dysregulated proteostasis which leads to greater variation in the levels of many proteins. Finally, the relative importance of the CSF proteome over the CSF metabolome, genome, and demographic information in predicting relevant AD biomarkers, seen not just here but in multiple studies where high prediction performance was achieved from the CSF proteome(*23, 31, 76*), further supports the unique relevance of the proteome in AD pathophysiology.

A few limitations deserve mention. First, the sample size and studied population were both limited. Though our sample size of 137 was comparable to other CSF proteomics work in AD, analysis in a larger sample would provide more precision, particularly for the novel joint multiomic analyses presented here. Our study was also limited to individuals of European ancestry, which limits its generalizability to broader populations. Another limitation of our study was the lack of the N (neurodegeneration) category in our main analyses. Expanding the categories from AT only to all relevant combinations of ATN would allow for more nuanced analysis. In a similar vein, having comparison groups for other non-AD causes of dementia would allow for better triangulation of AD-specific proteomic changes. We note as well that the Knight ADRC replication data set used a different proteomics platform from the discovery data set (aptamer-based vs. MS-based) and that the correlation between the MS-based and immunoassay-based measurements of YKL-40 and sTREM2 were only moderate. One the one hand, technical differences between the platforms could have introduced different biases or issues in protein identification that might have impacted the observed results. On the other hand, one way to deal with potential differences in proteomics platforms across cohorts is to seek replication through different technologies(*77, 78*). Here we used such a cross-platform approach. The presence of concordant replication of association for 3 out of 5 overlapping proteins despite the difference in platform shows the robustness of these CSF signals to the choice of proteomics platform. Furthermore, the additional replication of 8 CSF proteins in the Higginbotham et al. and 5 brain proteins in the Johnson et al. data sets provide further evidence of the robustness of the associations reported here. As additional large AD proteomics data sets develop with a variety of proteomics platforms and outcome measures, further replication of our results will be useful. Finally, instead of using the entire omic data sets (or outcome-blind predictor reductions, as was the case with the genetic data set), additional filtering steps that narrowed down to predictors more likely to be associated with AD would provide even better prediction for the amyloid and tau measures.

Nevertheless, our study provides a thorough investigation of the CSF proteome and its relationship to AD, AD biomarkers, and other omics. Among the numerous proteins associated with AD and CSF biomarkers of neurodegeneration and neuroinflammation, we demonstrated that the CSF proteome associated with amyloid and tau was enriched for glucose metabolic pathways in contrast to the other biomarkers whose associated proteins were enriched for more extracellular and structural pathways. Follow-up independent proteomic replication analyses and CSF metabolomics interrogation provided robust support for the increase in the CSF of these proteins and highlighted succinylcarnitine as a relevant metabolite, corroborating several previous lines of AD experimental and genomic evidence. In total, these results showcase the power of multiomic analyses and provide a new look at the CSF proteome in AD in relation to amyloid and tau.

## MATERIALS AND METHODS

### Experimental design

The data in this study came from two longitudinal AD cohorts of middle- and older-aged adults: the Wisconsin Registry for Alzheimer’s Prevention (WRAP)(*79*) and the Wisconsin Alzheimer’s Disease Research Center (ADRC)(*80*) (Table 1, Supplementary Figure 1). Briefly, WRAP includes participants enriched for a parental history of AD dementia who were largely between the ages of 40 and 65 at the time of enrollment, fluent in English, able to perform neuropsychological testing, without a diagnosis or evidence of dementia at baseline, and without any health conditions that might prevent participation in the study. The ADRC study includes participants from one of several subgroups: mild late-onset AD, MCI, cognitively unimpaired middle-aged adults enriched for a parental history of presumed AD dementia, and age-matched healthy older controls (age > 65). Briefly, the ADRC participants were over the age of 45, with decisional capacity, and without a history of certain medical conditions (like congestive heart failure or major neurologic disorders other than dementia) or any contraindication to biomarker procedures. Participants in both the WRAP and ADRC cohorts were given diagnoses of AD, MCI, cognitively unimpaired, and others that were reviewed by a consensus review committee that included dementia-specialist physicians, neuropsychologists, and nurse practitioners(*79*). The National Institute of Neurological and Communicative Disorders and Stroke and Alzheimer’s Disease and Related Disorders Association (NINCDS-ADRDA)(*6*) and NIA-AA(*7*) criteria were used in defining the clinical diagnoses without reference to the participants’ CSF biomarker status. This study used the STROBE cohort reporting guidelines(*81*) and was performed as part of the GeneRations Of WRAP (GROW) study, which was approved by the University of Wisconsin Health Sciences Institutional Review Board. Participants in the ADRC and WRAP studies provided written informed consent.

### CSF biomarkers

The CSF samples used for the biomarker analyses were acquired from lumbar punctures (LPs) using a uniform preanalytical protocol between 2010 and 2018 as previously described(*82*). Samples were collected in the morning using a Sprotte 24- or 25-gauge atraumatic spinal needle, and 22 mL of fluid was collected via gentle extraction into polypropylene syringes and combined into a single 30 mL polypropylene tube. After gentle mixing, samples were centrifuged to remove red blood cells and other debris. Then, 0.5 mL CSF was aliquoted into 1.5 mL polypropylene tubes and stored at -80 degrees Celsius within 30 minutes of collection.

All CSF samples were assayed between March 2019 and January 2020 at the Clinical Neurochemistry Laboratory at the University of Gothenburg. CSF biomarkers were assayed using the NeuroToolKit (NTK) (Roche Diagnostics International Ltd, Rotkreuz, Switzerland), a panel of automated Elecsys® and robust prototype immunoassays designed to generate reliable biomarker data that can be compared across cohorts. Measurements with the following immunoassays were performed on a cobas e 601 analyzer (Roche Diagnostics International Ltd, Rotkreuz, Switzerland): Elecsys β-amyloid (1–42) CSF (Aβ42), Elecsys Phospho-Tau (181P) CSF (ptau), and Elecsys Total-Tau CSF, β-amyloid (1–40) CSF (Aβ40), and interleukin-6 (IL-6). The remaining NTK panel was assayed on a cobas e 411 analyzer (Roche Diagnostics International Ltd, Rotkreuz, Switzerland), including markers of synaptic damage and neuronal degeneration (neurogranin, neurofilament light protein [NFL], and alpha-synuclein) and markers of glial activation (chitinase-3-like protein 1 [YKL-40] and soluble triggering receptor expressed on myeloid cells 2 [sTREM2]).

A total of nine established CSF biomarkers for AD were analyzed in this study: the Aβ42/Aβ40 ratio, ptau, the ptau/Aβ42 ratio, NFL, alpha-synuclein, neurogranin, YKL-40, sTREM2, and IL-6. Since the CSF biomarker measurements were to be used as outcomes, each biomarker was assessed for skewness using the skewness function of the R package moments (version 0.14)(*83*). Any biomarker with a skewness ≥ 2 was transformed with a log10-transformation to better meet the normality assumption of regression. The outcomes that were log10-transformed were NFL and IL-6.

Samples used in this study were then assigned to pathological categories from the NIA-AA ATN research framework(*11*) using binary cut-offs for CSF amyloid and tau positivity. The development of these research cut-offs is described in detail elsewhere(*82*). Briefly, cut-offs were estimated via ROC analysis on a subsample of n = 185 participants (cognitively impaired and unimpaired) who underwent [11C] PiB-PET imaging within two years of an LP. Using the Matlab perfcurve function(*84*) with an equally weighted cost function(*85*), the optimal Aβ42/Aβ40 threshold was 0.046 and the optimal ptau/Aβ42 threshold was 0.038. Thresholds for ptau181 were determined by establishing a reference group of 223 CSF amyloid (Aβ42/Aβ40) negative, cognitively unimpaired younger participants (ages 40-60 years).

Biomarker positivity thresholds for these analytes were set at +2 standard deviations (SD) above the mean of this reference group (ptau threshold = 24.8 pg/mL). In this study, A+ and T+ were defined based on the CSF Aβ42/Aβ40 and ptau thresholds, respectively. The final pathological categories for this study included amyloid negative and tau negative (A-T-); amyloid positive and tau negative (A+T-); and amyloid positive tau positive (A+T+). The fourth possible category of amyloid negative and tau positive (A-T+) was not included in this study as these samples are considered to represent non-AD pathological change(*11*).

### CSF metabolomics

All samples used in this study had CSF metabolomics data available from the WRAP and ADRC cohorts that had been generated in previous work. The details of the CSF sample collection, handling, and metabolomics profiling have been previously described(*86, 87*). Briefly, fasting CSF samples were drawn from study participants in the morning through LP and then mixed, centrifuged, aliquoted, and stored at -80 degrees Celsius. Samples were kept frozen until they were shipped overnight to Metabolon, Inc. (Durham, NC), which similarly kept samples frozen at -80 degrees Celsius until analysis. Metabolon used Ultrahigh Performance Liquid Chromatography-Tandem Mass Spectrometry (UPLC-MS/MS) to conduct an untargeted metabolomics analysis of the CSF samples. The metabolites were then annotated with metabolite identifiers, chemical properties, and pathway information. Metabolite measurements were divided by the median measurement for that metabolite across all samples. Missing values for xenobiotic metabolites were imputed to 0.0001, while missing values for non-xenobiotic metabolites were imputed to half of the minimum value among all other measured samples for that metabolite.

The initial data set contained 412 metabolites from 1,172 CSF samples across 687 unique individuals. A total of 13 metabolites that were missing for ≥ 50% of samples were removed. One sample was removed for missing ≥ 40% of metabolite values. A total of 9 metabolites with low variance (interquartile range = 0) were then removed. A log10 transformation was applied to all metabolite values. A total of 220 samples from a clinical trial were excluded from analysis. The processed data set contained 390 metabolites quantified on 951 CSF samples from 609 unique individuals, including all but one of the CSF samples on which proteomics data were generated for this study.

### Genome-wide genotyping

Genome-wide genotypes were also available for all samples in this study. The genotyping in both the WRAP and ADRC had been previously conducted(*88*). For the WRAP cohort, DNA from whole blood samples were genotyped with the Illumina Multi-Ethnic Genotyping Array at the University of Wisconsin Biotechnology Center(*87*). Pre-imputation quality control (QC) steps included removing samples and variants with a high missingness (> 5%) or inconsistent genetic and self-reported sex. Samples from individuals of European descent were imputed using the Michigan Imputation Server(*89*) and the Haplotype Reference Consortium (HRC) reference panel(*90*). Variants with poor quality (R^2^ < 0.8) or out of Hardy-Weinberg equilibrium (HWE) were removed after imputation, leaving a total of 1,198 samples with 10,499,994 single nucleotide polymorphisms (SNPs). In the ADRC, whole blood samples were genotyped by the Alzheimer’s Disease Genetics Consortium (ADGC) at the National Alzheimer’s Coordinating Center (NACC) using the Illumina HumanOmniExpress-12v1_A, Infinium HumanOmniExpressExome-8 v1-2a, or Infinium Global Screening Array v1-0 (GSAMD-24v1-0_20011747_A1) BeadChip assay. Initial quality control was conducted on each chip’s data separately, removing variants or samples with high missingness (> 2%), out of HWE (P < 1×10^-6^), or with inconsistent genetic and self-reported sex. The remaining samples were then imputed with the Michigan Imputation Server, phased using Eagle2(*91*), and imputed to the HRC reference panel. As before, variants of low quality (R^2^ < 0.8) or out of HWE were removed. The data sets from the different chips were then merged together, leaving a data set with 377 samples of European descent and 7,049,703 SNPs. The WRAP and ADRC data sets were then harmonized to each other and to the 1000 Genomes Utah residents with Northern and Western European ancestry (CEU)(*92*) data set, using the GRCh37 genome build. Ambiguous SNPs were removed, and then the remaining SNPs were aligned to the same strand and allele orientations as the ADRC data set.

The 137 samples from this study were then extracted from this combined genetic data set and further processed using PLINK(*93*) (v1.90b6.3). To ensure sufficient data were available for use in the prediction models, only SNPs with no missing data and with a minor allele count of 20 or greater among the 137 samples were retained. Linkage disequilibrium (LD) pruning was then applied using a window size of 1000 kb, an R^2^ threshold of 0.1, and the 1000 Genomes CEU samples as the reference data set. The pruning resulted in a data set of 38,652 SNPs.

### APOE genotyping

Each sample was additionally assigned an *APOE* genotype based on the participant’s combination of the ε2, ε3, and ε4 alleles for *APOE* from a separate set of genotyping. DNA was extracted from whole blood samples, which was then genotyped for the *APOE* alleles using competitive allele-specific PCR-based KASP genotyping for rs429358 and rs7412(*86*).

### Proteomics sample selection

Based on the results of our pilot study for CSF proteomics(*25*), we had estimated *a priori* that a sample of approximately 150 would be sufficient for 80% power to detect most of the observed protein-AD diagnosis associations from the original matched case-control analyses in the pilot using the R package pwr (version 1.3-0)(*94*), though we note that the final study design used here differed from the pilot in that three participant groups were used and age and sex were controlled for in the analyses instead of using a matched design (Supplementary Figure 2). The process of selecting samples for CSF proteomics generation began by considering all CSF samples from fasted, successful LPs (n = 1,440) from 823 unique participants across WRAP and ADRC. From there, each CSF sample was matched to its closest set of CSF biomarker data, CSF metabolomics data, and consensus conference diagnosis. Samples were excluded if there was insufficient material for proteomics analysis, if they were part of a clinical trial, or if they had been used already in our pilot study. To simplify the downstream analyses, only one sample (the most recent) per participant was considered when there were multiple samples. An approximately equal number of samples per AT-defined subgroup (A-T-, A+T-, A+T+) was selected, prioritizing samples with available genomic data and metabolomic data. A total of 140 samples were selected to have proteomics data generated.

### Protein extraction and digestion

CSF protein concentration was determined by protein BCA assay (Thermo Scientific). CSF aliquots were moved to 96-plates and dried down using a SpeedVac Concentrator (Thermo Scientific) before being resuspended in a lysis buffer consisting of 10 mM TCEP, 40 mM CAA, 100 mM Tris pH 8, and 8M urea. The sample solution was then diluted to 25% strength using 100 mM Tris pH 8 before the addition of protease. Trypsin was added to the protein solution at an approximate ratio of 50:1 w/w and digested overnight at ambient temperature. The digestion reaction was quenched by acidification using TFA. Digested peptides were desalted using Strata-X Polymeric Reverse Phase plates (Phenomenex) before being dried down in the SpeedVac Concentrator overnight. Dried down samples were resuspended in 0.2% FA and peptide concentration was determined using a peptide BCA (Thermo Scientific). Peptide samples were injected directly from the 96-well plates.

### Offline fractionation

In order to increase proteomic depth and protein identifications, offline chromatographic fractionation of a set of pooled representative samples was performed. Pooled samples for each of the three disease groups were created by combining 10 µL of CSF from each sample in that disease group. These three pooled samples were then prepared using the extraction and digestion protocol described above. The three desalted, digested peptide solutions were then fractionated using high-pH reverse-phase liquid chromatography. Separation was performed using an Agilent Infinity 2000 HPLC with a 150 mm C18 reverse-phase column (Waters, XBridge Peptide BEH, particle size 3.5 µm). Mobile phase buffer A was a freshly prepared mixture 10 mM ammonium formate pH 9.5, and mobile phase buffer B was a freshly prepared mixture of 80% MeOH, 10 mM ammonium formate pH 9.5. The gradient method was 20 minutes in length with fractions collected from minute 5 to minute 20, with a flow a rate of 800 nL/min across the entire method. The method initiated with a concentration of 5% B before increasing to 35% by minute 2. Percent B increased to 100% by 13 minutes. From 5 to 20 minutes, 32 fractions were collected in round-bottom 96 well plates in a time-based manner. Fractions were concatenated into a total of 16 by combining every other column in the collection plate. Fractionated samples were injected directly from the collection plate.

### Online chromatography

To quantify the proteins in the individual CSF samples, we used a method previously developed in our pilot study(*25*): a single-shot nano-liquid chromatography-tandem mass spectrometry (nLC-MS/MS) method for quantitative and fast analysis of CSF protein extracts. Reverse phase columns used online with the mass spectrometer were packed using an in-house column packing apparatus described previously(*95*). In brief, 1.5 µm BEH particles were packed into a fused silica capillary purchased from New Objective (PicoTip, Stock # PF360-75-10-N-5) at 30,000 psi. During online LC separations, capillary was heated to 50° C and interfaced with mass spectrometer via an embedded emitter. For online chromatography, a Dionex UltiMate 300 nanoflow UHPLC was used with mobile phase A consisting of 0.2% FA and mobile phase B consisted of 70% ACN, 0.2% FA. A flow rate of 310 nL/min was used throughout with the method increasing from 0% to 7% B over the first four minutes. Percent B then increased to 49% B by 59 minutes before a wash step of 100% B from 62 to 67 minutes. Method finished with an equilibration step from minute 68 to 78 of 0% B.

### Tandem mass spectrometry

Peptides eluting from the column were ionized by electrospray ionization and analyzed using a Thermo Orbitrap Eclipse hybrid mass spectrometer. Survey scans were collected in the Orbitrap at a resolution of 240,000 with a normalized AGC target of 250% (1e6) with Advanced Precursor Determination engaged across the range of 300–1400 m/z. Precursors were isolated for tandem MS scans using a window of 0.5 m/z, with a dynamic exclusion duration of 22 seconds and a mass tolerance of 15 ppm. Precursors were dissociated using HCD with a normalized collision energy of 25%. Tandem scans were taken over the range 130–1350 m/z using the “rapid” setting with a normalized AGC target of 300% (3e4) and a maximum injection time of 18 ms.

The resulting raw data files were searched in MaxQuant(*96, 97*) using fast LFQ and a full human proteome with isoforms downloaded from UniProt (downloaded June 14, 2017). Oxidation of methionine and acetylation of the N terminus were allowed as variable modifications, and carbamidomethylation of cysteine was set as a fixed modification. Proteins were searched using a false discovery rate (FDR) of 1% with a minimum peptide length of 7 and a 0.5 Da MS/MS match tolerance. Matching between runs was utilized, applied with a retention time window of 0.7 minutes. Protein abundance data were extracted in the form of LFQ Intensity from the “proteinGroups.txt” output file. Throughout this manuscript, each protein group is referred to by the first listed majority protein from its annotation from MaxQuant. The protein data were annotated with Entrez IDs (via R package org.Hs.eg.db(*98*), version 3.11.4), UniProt(*99*) IDs, and gene information (GENCODE(*100*), version 37, and the HUGO Gene Nomenclature Committee, HGNC, database(*101*)). When the gene annotations conflicted or were absent from one of these databases for a given UniProt ID, the gene identifiers were taken in the order of resources listed. Although the LC-MS analysis was applied to individual samples as well as the three pooled and fractionated samples, only the individual samples were used for quantitative analysis and statistical investigations described hereafter.

### Proteomics quality control

After removing several samples with injection or other technical issues, the proteomics data set included 2,040 proteins across 137 samples. These data underwent a strict quality control pipeline: proteins that were missing for 33% or more of samples (either overall or within an AT category) were removed; samples missing 33% or more of proteins were removed; and proteins with an interquartile range of 0 were removed (Supplementary Figure 3, Supplementary Figure 4). A total of 137 samples with 915 proteins remained (Supplementary Table 1). The label-free quantification (LFQ) values for each protein were then log2-transformed. The remaining missing values were then randomly imputed based on a normal distribution derived from the lower end of the observed values for that protein (the observed distribution mean was shifted by -1.8 and the SD shrunk by a factor of 0.3) (Supplementary Figure 5). This imputation was performed separately within each AT category. Finally, each protein was Z-score transformed.

### Statistical analysis

### Proteomics descriptive analysis

The first step in understanding how the CSF proteome changes in AD is to first understand what its contents are and how it compares to the entirety of the human proteome. Thus, our first main objective was to extensively profile the set of proteins quantified in the CSF in this cohort (Supplementary Table 2). The pairwise correlation of all proteins was calculated (nominally significant results with correlation P < 0.05 shown in Supplementary Table 3) and then visualized with a heatmap with hierarchical clustering to show the underlying patterns of covariation (R package ComplexHeatmap(*102*), version 2.4.3) (Figure 1a). The structure was further examined with a principal components analysis (PCA), scree plot (Supplementary Figure 6), and plot of the first two PCs by AT category (R package factoextra(*103*), version 1.0.7) (Supplementary Figure 7) to assess the presence of independent signals among the CSF proteome and their relationship to the AT categories. The associations of the top two PCs with age were also examined with a correlation analysis. A pathway analysis was then performed to examine the differences between the set of proteins quantified in the CSF and the overall human proteome. The enrichment of Gene Ontology (GO) terms(*104*), Kyoto Encyclopedia of Genes and Genomes (KEGG) pathways(*104, 105*), and Disease Ontology (DO) gene sets(*106*) among the CSF proteome against the entire human proteome was calculated using the R packages clusterProfiler(*107*) (version 3.16.1), DOSE(*108*) (version 3.14.0), and ReactomePA(*109*) (version 1.32.0) (Figure 1b, Figure 1c, Supplementary Table 4). To summarize the major constituents of the observed CSF proteome in these cohorts, the presence of clusters was then assessed with Gaussian mixture modeling using the R package mclust(*110*) (version 5.4.6). The number of clusters (3) was chosen based on the elbow of the plot of the Bayesian Information Criterion (BIC) (Supplementary Figure 8). Enrichment of gene set ontologies across the clusters was repeated with the GO, KEGG, and DO sets and plotted (Figure 1d, Supplementary Table 5).

### Protein-AT category associations

The second main objective of this study was the identification of differentially expressed proteins across the three AT categories in order to understand how the CSF proteome changes across the AD trajectory. This analysis was performed using an analysis of covariance (ANCOVA) model comparing each protein across the three groups, controlling for age at LP and sex (Supplementary Table 6). Additionally, to test the difference in protein level between each pair of AT categories, a logistic regression model was used for each pair of categories (controlling for age at LP and sex) (Supplementary Table 6). A Bonferroni correction for the number of proteins tested (P = 0.05 / 915 = 5.46 x 10^-5^) was used for reporting significant results. The distributions of the top-associated proteins across the AT spectrum were plotted (Figure 2a). To assess whether signal enrichment was likely due to an artifact, the ANCOVA analyses were repeated with randomly permuted AT category labels. A quantile-quantile (Q-Q) plot was generated to assess the presence of signal enrichment across the proteome for AT-related differences and to compare the permuted and non-permuted analyses (Figure 2b). Since the *APOE* gene is known to have a significant effect on AD risk, we examined whether *APOE* genotype was driving the observed AT-protein associations. The ANCOVA analyses were repeated but with the count of *APOE* ε4 alleles included as an additional covariate. The same Bonferroni correction was used as before. The resulting AT-associated proteins were compared to the results from the original ANCOVA analyses (Supplementary Figure 9). The set of associated proteins from the non-permuted analysis was then assessed for enriched GO, KEGG, and DO gene sets against the human proteome as before (Figure 2c, Supplementary Table 7).

To examine the direction of effect of each protein, a logistic regression was performed with A+T+ (vs. A-T-) as the outcome and a protein as the main predictor, controlling for age at LP and sex and using the same Bonferroni threshold for significance as the ANCOVA analyses. The sample size for the logistic regression was smaller (n = 98) due to the exclusion of the A+T-samples. The overlap between the set of significantly associated proteins and the set of significantly associated proteins from the ANCOVA analysis was displayed in a Venn diagram (R package ggVennDiagram(*111*), version 1.0.7) (Supplementary Figure 10), and the odds ratios were presented in a volcano plot (Supplementary Figure 11).

### Protein-CSF biomarker associations

While understanding the relationships of the proteome with amyloid and tau status can provide a coarse view of the changing AD proteome, it is useful to investigate more nuanced connections with markers of neurodegeneration and neuroinflammation in part to be able to identify proteins that may be related to dementia but not necessarily specific to AD. To this end, our third main objective was conducting a comprehensive set of protein-CSF biomarker analyses using the NTK panel described above. Each protein was tested for association with each of the 9 CSF biomarkers (the Aβ42/Aβ40 ratio, ptau, the ptau/Aβ42 ratio, NFL, alpha-synuclein, neurogranin, YKL-40, sTREM2, and IL-6). Linear regression models were used to regress each CSF biomarker on each protein, controlling for age at CSF sample and sex and using a Bonferroni correction for the total number of tests (9 x 915 = 8,234 tests; P = 0.05 / 8,234 = 6.07 x 10^-6^) (Table 2, Supplementary Table 8). For the sake of interpretation, effect estimates where the biomarker outcome was additionally Z-score normalized were also provided (“Effect size (std. biomarker)” columns). The results were summarized with a Q-Q plot showing the signal enrichment for each biomarker along with a sensitivity analysis where the regression models were repeated with the biomarker values randomly permuted to test the robustness of each biomarker’s signal enrichment (Supplementary Figure 12). The cross-biomarker relationships among the significantly associated proteins were then visualized as a bipartite graph using the R package tidygraph (version 1.2.0)(*112*) and the Fruchterman-Reingold algorithm. A community structure network analysis was performed using the greedy hierarchical agglomeration algorithm(*113*) implemented in igraph (version 1.2.5)(*114*) to identify clusters of proteins among the protein-biomarker associations. An upset plot was then created showing the set of significantly associated proteins unique to each subset of biomarkers (Supplementary Figure 13) using the UpSetR package (version 1.4.0)(*115*). Pathway enrichment analyses were performed as before comparing each biomarker’s set of significantly associated proteins (Supplementary Table 9).

### Replication of top pathway results in other AD proteomics cohorts

A replication data set from the Knight ADRC was used first to validate findings from the main analyses. The Knight ADRC data set included samples from CSF (n = 717) and plasma (n = 490). The recruited individuals from the Knight ADRC cohort were evaluated by Clinical Core personnel of Washington University. For individuals with CSF and plasma data, cases received a clinical diagnosis of AD in accordance with standard criteria, and AD severity was determined using the clinical dementia rating (CDR) scale(*116*) at the time of lumbar puncture (for CSF samples) or blood draw (for plasma samples). CSF was collected by LP after overnight fasting, centrifuged, and frozen at −80 °C as described previously(*117–119*). Blood was collected at the time of LP, and serum or plasma was obtained by centrifugation and stored at −80 °C. CSF samples were analyzed by immunoassay for β-amyloid 1–42 (Aβ42), total tau, and tau phosphorylated at threonine 181 (p-tau) (INNOTEST, Fujirebio, Ghent, Belgium). The Institutional Review Boards of Washington University School of Medicine in St. Louis approved the study; research was performed in accordance with the approved protocols and participants provided informed consent.

For deep proteomics characterization in the CSF and plasma tissues, the levels of 1,305 proteins were quantified using a different methodological approach from that used for the Wisconsin cohorts: the SOMAscan assay, a multiplexed, aptamer-based platform(*120*). The assay covers a dynamic range of 10^8^ and measures all three major categories: secreted, membrane, and intracellular proteins. The proteins cover a wide range of molecular functions and include proteins known to be relevant to human disease. As previously described by Gold et al.(*120*), modified single-stranded DNA aptamers are used to bind specific protein targets that are then quantified by a DNA microarray. Protein concentrations are quantified as relative fluorescent units (RFU). Aliquots of 150 μL of tissue were sent to the Genome Technology Access Center at Washington University in St. Louis for protein measurement.

Quality control was performed at the sample and aptamer levels using control aptamers (positive and negative controls) and calibrator samples. At the sample level, hybridization controls on each plate were used to correct for systematic variability in hybridization. The median signal over all aptamers was used to correct for within-run technical variability. This median signal was assigned to different dilution sets within each tissue. For CSF samples, a 20% dilution rate was used. For plasma samples, three different dilution sets (40%, 1%, and 0.005%) were used.

As described in detail(*121*), additional quality control was performed by identifying and removing protein and analyte outliers by applying four criteria: 1) Minimum detection filtering. If the analyte for a given sample was less than the limit of detection (LOD), the sample was deemed an outlier. Collectively, if the number of outliers given an analyte was less than 15% of the total sample size, the analyte was kept. 2) Flagging analytes based on the scale factor difference. 3) Coefficient of variation (CV) of calibrators lower than 0.15, where the CV for each aptamer was calculated as the standard deviation divided by the mean of each calibrator at the raw protein level. 4) Interquartile range (IQR) strategy.

Outliers were identified if the subject was located 1.5-fold of the IQR outside of either end of the distribution given the log10-transformation of the protein level. Analytes were kept after passing all the criteria above for the downstream statistical analysis. An orthogonal approach was used to call subject outliers based on IQR. After this second removal of analytes, subject outliers were examined and removed again.

To obtain the proteomic signatures of sporadic AD status, CSF Aβ42, ptau/Aβ42, and ptau, differential abundance analysis was performed by using linear regression of the log-transformed protein levels. In each tissue, we performed surrogate variable analysis while protecting status and age to correct for unmeasured heterogeneity(*122*). Age at death or at measurement, gender, and the resulting surrogate variables were included as covariates.

The Knight ADRC analyses were used as a replication data set for the top findings from the main analyses performed in the Wisconsin ADRC and WRAP, focusing on the significantly associated proteins from the top implicated biological pathway from the protein-AT category and protein-biomarker analyses. The association of these proteins were compared to the results from the Knight ADRC association tests conducted in the CSF and plasma to see if their associations and directions of effect were replicated using a significance threshold corrected for the number of tested proteins in the replication (P = 0.05 / 9 = 0.0056) (Table 3, Supplementary Table 10).

To provide replication using a more similar proteomics methodology to what was used for discovery analysis here, the top findings were assessed against the published results from Higginbotham et al. 2020(*26*) and Johnson et al. 2020(*24*). Both of these data sets used a mass spectrometry-based approach for proteomics and reported differential protein expression results based on clinical diagnoses. Higginbotham et al. reported differences in CSF protein levels between AD and control participants (n = 20) while Johnson et al. reported differences in brain protein levels between healthy controls, asymptomatic AD, and AD participants (n = 453). A nominal P value threshold (P = 0.05) was used for replication.

### Secondary analysis of insulin-related proteins

Based on the results of the AT category and biomarker associations and the pathway analysis suggesting a relationship with glucose regulation (described below), the set of proteins excluded during the QC process due to low sample size was examined for proteins related to insulin signaling pathways, including any of the GLUT proteins (SLC2A family), insulin (INS), insulin receptor (INSR), insulin-like growth factor 1 (IGF1), IGF-1 receptor (IGF1R), insulin receptor substrate 1 (IRS1), IRS 2 (IRS2), phosphoinositide 3-kinase (PI3K), RAC-alpha serine/threonine-protein kinase (AKT1), mechanistic target of rapamycin (mTOR), and glycogen synthase kinase 3 (GSK3A). Proteins that failed the missingness threshold of 33% but were present for 50% or more of samples were investigated further but without the use of imputed data points. The relationships between the proteins and AT category (Supplementary Figure 14) and the CSF biomarkers (Supplementary Figure 15) were plotted, with ANCOVA and linear regression analyses to test for association between the proteins and AT category and the CSF biomarkers performed as previously (Supplementary Table 11).

### Secondary analysis of glycolysis and TCA cycle metabolites

In order to see if the implicated pathways observed from the proteomics analyses were also implicated within the metabolomics data, a second form of validation of the main proteomics findings around glucose metabolism was performed using the CSF metabolomics data available in the WRAP and WI ADRC cohorts (described above). Focusing on the 10 available metabolites from the “Glycolysis, gluconeogenesis, and pyruvate metabolism” and “TCA cycle” superpathways (namely, 1,5-anhydroglucitol (1,5-AG), alpha-ketoglutarate, citrate, glucose, glycerate, isocitrate, lactate, malate, pyruvate, and succinylcarnitine (C4-DC)) (Supplementary Table 12), the same AT category and CSF biomarker association analyses performed for the proteomics data were performed again but with these 10 metabolites instead of the proteins. The discovery phase of this analysis was performed using the 136 participant CSF samples from the proteomics analysis that also had CSF metabolomic data available from the same matched CSF samples. The results of the analysis were subjected to a Bonferroni-corrected P value threshold based on the number of metabolites tested (P = 0.05 / 10 metabolites = 0.005 for the metabolite-AT category association analyses; P = 0.05 / 10 metabolites / 9 biomarkers = 0.00056 for the metabolite-biomarker association analyses). Significantly associated metabolites were visualized with box plots (Supplementary Figure 16, Supplementary Table 13, Supplementary Table 14).

Because the group of participants in the WRAP and ADRC cohorts with CSF proteomics data generated here was a subset of all of the participants with previously generated CSF metabolomics data, there were an additional 363 unique participants with CSF metabolomics data whose samples were not included in the main proteomics work here (Supplementary Table 15). These participants’ CSF metabolomics data were used as an independent replication data set for this secondary metabolomics analysis. The same AT category and CSF biomarker association analyses were repeated, using the same Bonferroni-corrected P value thresholds as in the metabolomics discovery analyses (Supplementary Figure 16, Supplementary Table 13, Supplementary Table 14).

### Multiomic prediction of amyloid and tau

With the conceptual shift toward defining AD with amyloid and tau biomarkers in a research context(*11*), understanding exactly what other aspects of biology correspond to amyloid and tau, especially in the CSF, is critical, but the relative connections between the different biological omes and amyloid and tau is unclear. Our fourth main objective was to investigate these relationships. We conducted a separate and joint predictive analysis of amyloid and tau categories using CSF proteomics, CSF metabolomics, genomics, and demographic information. The CSF proteomic data set was combined with the CSF metabolomic, genomic, and demographic (age at sample and sex) data sets. After the quality control steps described previously for each ome, 136 of the 137 CSF samples had values for all of the multiomic features (915 proteins, 390 metabolites, 38,652 SNPs coded as dosages plus the *APOE* ε4 allele count, and 2 demographic features) (one sample was excluded for lacking CSF metabolomics data). This multiomic data set was then used to predict different biomarker positivity states(*82*): Aβ42/Aβ40-positive, ptau-positive, and ptau/Aβ42-positive. Each ome (CSF proteome, CSF metabolome, genome, and demographics) was used individually along with a fifth multiomics predictor set (comprising all omes) to predict each outcome with an elastic net(*123*) model (R package glmnet(*124*), version 4.0-2; alpha parameter = 0.5). When different CSF-based measurements were used for an individual (e.g., MS-derived CSF protein level, CSF biomarkers from the NTK platform, CSF metabolite levels, etc.), those measurements were all performed on or refer to the same underlying CSF sample; there was no sample date discrepancy between CSF measurements for a given participant.

For each biomarker and predictor pair, the procedure was the same. First, one-third of the data was held out as a testing set and the remaining two-thirds used as the training set. Within the training set data, 100-iteration, 3-fold cross-validation was used to select the best lambda value (11 possible values ranging from 10^-5^ to 1) according to AUC using the tidymodels(*125*) R package (version 0.1.3). The best-performing model was then run on the entire training data set using the chosen lambda and used to predict the outcome on the held-out testing data set. The performances of the different omic models were then compared with ROC curves and 2D histograms showing the raw biomarker levels against the predicted classifications for each biomarker for each subject (Figure 4). The mean model metrics across each of the 4,000 folds were calculated (Supplementary Table 16).

## List of Supplementary Materials

Supplementary Figure 1: Data and analytical overview

Supplementary Figure 2: Pilot study power analysis

Supplementary Figure 3: Protein missingness overall

Supplementary Figure 4: Protein missingness by AT category

Supplementary Figure 5: Protein imputation examples

Supplementary Figure 6: CSF proteomics PCA scree plot

Supplementary Figure 7: CSF proteomics PC plots

Supplementary Figure 8: Clustering BIC values

Supplementary Figure 9: Overlap of ANCOVA and APOE-controlled results

Supplementary Figure 10: Overlap of ANCOVA and logistic regression results

Supplementary Figure 11: Significantly associated proteins with A+T+ vs A-T-

Supplementary Figure 12: Protein-biomarker association Q-Q plot

Supplementary Figure 13: Overlap in the proteins associated among the biomarkers

Supplementary Figure 14: Distribution of IGF-1 by AT category

Supplementary Figure 15: Relationships between IGF-1 and CSF biomarkers

Supplementary Figure 16: Distribution of succinylcarnitine (C4-DC) by AT category

Supplementary Table 1: Protein quality control steps

Supplementary Table 2: Protein information

Supplementary Table 3: Nominally significant pairwise protein correlations

Supplementary Table 4: Significantly enriched pathways among the CSF proteome compared to the human proteome

Supplementary Table 5: Significantly enriched pathways across CSF proteome clusters

Supplementary Table 6: Protein-AT category association test results

Supplementary Table 7: Significantly enriched pathways among proteins associated with AT category

Supplementary Table 8: Protein-biomarker association test results

Supplementary Table 9: Significantly enriched pathways among proteins significantly associated with biomarkers

Supplementary Table 10: Associations of plasma levels of glucose metabolism-related proteins with AD biomarkers in the Knight ADRC

Supplementary Table 11: IGF-1-biomarker association test results

Supplementary Table 12: Metabolite information

Supplementary Table 13: Metabolite-AT category association test results

Supplementary Table 14: Metabolite-biomarker association test results

Supplementary Table 15: Summary of metabolomics replication sample

Supplementary Table 16: Summary of mean multiomic biomarker prediction model performance

## Data Availability

The data sets generated and analyzed in this study from the Wisconsin ADRC may be requested at https://www.adrc.wisc.edu/apply-resources. The Knight-ADRC proteomic data is available at NIAGADS: NG00102 collection and can be interactively explored at http://ngi.pub:3838/ONTIME_Proteomics/.

http://ngi.pub:3838/ONTIME_Proteomics/

## Acknowledgments

We would like to thank WRAP and ADRC participants and the Wisconsin Alzheimer’s Institute (WAI) and ADRC staff for their contributions to the WRAP and ADRC studies. Without their efforts this research would not be possible.

## Author contributions

Authors DJP, JM, JJC, and CDE conceived the study design and conducted the initial pilot study. Author DJP conducted the quality control, data integration, and analyses of the various Wisconsin data sets; prepared the figures and tables; and led the writing of the manuscript. Authors JM and JJC generated the Wisconsin proteomics data. Authors YKD, ARM, GEE, and BBB contributed to the secondary analyses and interpretations of the main proteomics findings. Author EMJ contributed to the statistical design of the study. Author CAVH contributed to the use and development of the biomarker-based categorizations. Authors CC, CY, YS, and MA conducted the replication analyses in the Knight ADRC data. Authors HZ, KB, GK, IS, and AB contributed to the generation of the NTK CSF biomarker data set. Authors CDE, CMC, SCJ, BBB, SA, JJC, HZ, and KB contributed resources or funding. Authors CMC, SCJ, BBB, SA, and CDE contributed to cohort sample or data collection. Authors CMC, SCJ, and SA oversaw the Wisconsin Alzheimer’s research cohort studies. All authors contributed to and critically reviewed the manuscript.

## Data and materials availability

Version information of the software used in this study is provided in the Methods.

## Tables

Table 1: Summary of sample demographics and CSF biomarkers

Table 2: Top 3 significantly associated proteins for each biomarker

Table 3: Replication of implicated glucose metabolism-related proteins in the Knight ADRC

## Figures

Figure 1: CSF proteomics descriptive analyses

Figure 2: Proteins associated with AT category

Figure 3: Network analysis of protein-biomarker associations

Figure 4: Multiomic predictive model performance for amyloid and tau

## References and Notes

1. Alzheimer’s Disease International, World Alzheimer Report 2019: Attitudes to dementia (Alzheimer’s Disease International, London, 2019), p. 160.

2. 2019 Alzheimer’s disease facts and figures. Alzheimers Dement. 15, 321–387 (2019).

3. A. Kumar, A. Singh, Ekavali, A review on Alzheimer’s disease pathophysiology and its management: an update. Pharmacol. Rep. 67, 195–203 (2015).

4. H. V. Vinters, Emerging Concepts in Alzheimer’s Disease. Annu. Rev. Pathol. Mech. Dis. 10, 291–319 (2015).

5. K. Blennow, H. Hampel, M. Weiner, H. Zetterberg, Cerebrospinal fluid and plasma biomarkers in Alzheimer disease. Nat. Rev. Neurol. 6, 131–144 (2010).

6. G. McKhann, D. Drachman, M. Folstein, R. Katzman, D. Price, E. M. Stadlan, Clinical diagnosis of Alzheimer’s disease: report of the NINCDS-ADRDA Work Group under the auspices of Department of Health and Human Services Task Force on Alzheimer’s Disease. Neurology 34, 939–944 (1984).

7. C. R. Jack, M. Albert, D. S. Knopman, G. M. McKhann, R. A. Sperling, M. Carillo, W. Thies, C. H. Phelps, Introduction to Revised Criteria for the Diagnosis of Alzheimer’s Disease: National Institute on Aging and the Alzheimer Association Workgroups. Alzheimers Dement. 7, 257–262 (2011).

8. R. A. Sperling, P. S. Aisen, L. A. Beckett, D. A. Bennett, S. Craft, A. M. Fagan, T. Iwatsubo, C. R. Jack, J. Kaye, T. J. Montine, D. C. Park, E. M. Reiman, C. C. Rowe, E. Siemers, Y. Stern, K. Yaffe, M. C. Carrillo, B. Thies, M. Morrison-Bogorad, M. V. Wagster, C. H. Phelps, Toward defining the preclinical stages of Alzheimer’s disease: Recommendations from the National Institute on Aging-Alzheimer’s Association workgroups on diagnostic guidelines for Alzheimer’s disease. Alzheimers Dement. 7, 280–292 (2011).

9. M. S. Albert, S. T. DeKosky, D. Dickson, B. Dubois, H. H. Feldman, N. C. Fox, A. Gamst, D. M. Holtzman, W. J. Jagust, R. C. Petersen, P. J. Snyder, M. C. Carrillo, B. Thies, C. H. Phelps, The diagnosis of mild cognitive impairment due to Alzheimer’s disease: Recommendations from the National Institute on Aging-Alzheimer’s Association workgroups on diagnostic guidelines for Alzheimer’s disease. Alzheimers Dement. 7, 270–279 (2011).

10. G. M. McKhann, D. S. Knopman, H. Chertkow, B. T. Hyman, C. R. Jack, C. H. Kawas, W. E. Klunk, W. J. Koroshetz, J. J. Manly, R. Mayeux, R. C. Mohs, J. C. Morris, M. N. Rossor, P. Scheltens, M. C. Carrillo, B. Thies, S. Weintraub, C. H. Phelps, The diagnosis of dementia due to Alzheimer’s disease: Recommendations from the National Institute on Aging-Alzheimer’s Association workgroups on diagnostic guidelines for Alzheimer’s disease. Alzheimers Dement. 7, 263–269 (2011).

11. C. R. Jack, D. A. Bennett, K. Blennow, M. C. Carrillo, B. Dunn, S. B. Haeberlein, D. M. Holtzman, W. Jagust, F. Jessen, J. Karlawish, E. Liu, J. L. Molinuevo, T. Montine, C. Phelps, K. P. Rankin, C. C. Rowe, P. Scheltens, E. Siemers, H. M. Snyder, R. Sperling, NIA-AA Research Framework: Toward a biological definition of Alzheimer’s disease. Alzheimers Dement. 14, 535–562 (2018).

12. K. J. Karczewski, M. P. Snyder, Integrative omics for health and disease. Nat. Rev. Genet. 19, 299–310 (2018).

13. S. M. Neuner, J. Tcw, A. M. Goate, Genetic architecture of Alzheimer’s disease. Neurobiol. Dis. 143, 104976 (2020).

14. B. W. Kunkle, B. Grenier-Boley, R. Sims, J. C. Bis, V. Damotte, A. C. Naj, A. Boland, M. Vronskaya, S. J. van der Lee, A. Amlie-Wolf, C. Bellenguez, A. Frizatti, V. Chouraki, E. R. Martin, K. Sleegers, N. Badarinarayan, J. Jakobsdottir, K. L. Hamilton-Nelson, S. Moreno-Grau, R. Olaso, R. Raybould, Y. Chen, A. B. Kuzma, M. Hiltunen, T. Morgan, S. Ahmad, B. N. Vardarajan, J. Epelbaum, P. Hoffmann, M. Boada, G. W. Beecham, J.-G. Garnier, D. Harold, A. L. Fitzpatrick, O. Valladares, M.-L. Moutet, A. Gerrish, A. V. Smith, L. Qu, D. Bacq, N. Denning, X. Jian, Y. Zhao, M. D. Zompo, N. C. Fox, S.-H. Choi, I. Mateo, J. T. Hughes, H. H. Adams, J. Malamon, F. Sanchez-Garcia, Y. Patel, J. A. Brody, B. A. Dombroski, M. C. D. Naranjo, M. Daniilidou, G. Eiriksdottir, S. Mukherjee, D. Wallon, J. Uphill, T. Aspelund, L. B. Cantwell, F. Garzia, D. Galimberti, E. Hofer, M. Butkiewicz, B. Fin, E. Scarpini, C. Sarnowski, W. S. Bush, S. Meslage, J. Kornhuber, C. C. White, Y. Song, R. C. Barber, S. Engelborghs, S. Sordon, D. Voijnovic, P. M. Adams, R. Vandenberghe, M. Mayhaus, L. A. Cupples, M. S. Albert, P. P. D. Deyn, W. Gu, J. J. Himali, D. Beekly, A. Squassina, A. M. Hartmann, A. Orellana, D. Blacker, E. Rodriguez-Rodriguez, S. Lovestone, M. E. Garcia, R. S. Doody, C. Munoz-Fernadez, R. Sussams, H. Lin, T. J. Fairchild, Y. A. Benito, C. Holmes, H. Karamujić-Čomić, M. P. Frosch, H. Thonberg, W. Maier, G. Roschupkin, B. Ghetti, V. Giedraitis, A. Kawalia, S. Li, R. M. Huebinger, L. Kilander, S. Moebus, I. Hernández, M. I. Kamboh, R. Brundin, J. Turton, Q. Yang, M. J. Katz, L. Concari, J. Lord, A. S. Beiser, C. D. Keene, S. Helisalmi, I. Kloszewska, W. A. Kukull, A. M. Koivisto, A. Lynch, L. Tarraga, E. B. Larson, A. Haapasalo, B. Lawlor, T. H. Mosley, R. B. Lipton, V. Solfrizzi, M. Gill, W. T. Longstreth, T. J. Montine, V. Frisardi, M. Diez-Fairen, F. Rivadeneira, R. C. Petersen, V. Deramecourt, I. Alvarez, F. Salani, A. Ciaramella, E. Boerwinkle, E. M. Reiman, N. Fievet, J. I. Rotter, J. S. Reisch, O. Hanon, C. Cupidi, A. G. A. Uitterlinden, D. R. Royall, C. Dufouil, R. G. Maletta, I. de Rojas, M. Sano, A. Brice, R. Cecchetti, P. S. George-Hyslop, K. Ritchie, M. Tsolaki, D. W. Tsuang, B. Dubois, D. Craig, C.-K. Wu, H. Soininen, D. Avramidou, R. L. Albin, L. Fratiglioni, A. Germanou, L. G. Apostolova, L. Keller, M. Koutroumani, S. E. Arnold, F. Panza, O. Gkatzima, S. Asthana, D. Hannequin, P. Whitehead, C. S. Atwood, P. Caffarra, H. Hampel, I. Quintela, Á. Carracedo, L. Lannfelt, D. C. Rubinsztein, L. L. Barnes, F. Pasquier, L. Frölich, S. Barral, B. McGuinness, T. G. Beach, J. A. Johnston, J. T. Becker, P. Passmore, E. H. Bigio, J. M. Schott, T. D. Bird, J. D. Warren, B. F. Boeve, M. K. Lupton, J. D. Bowen, P. Proitsi, A. Boxer, J. F. Powell, J. R. Burke, J. S. K. Kauwe, J. M. Burns, M. Mancuso, J. D. Buxbaum, U. Bonuccelli, N. J. Cairns, McQuillin, C. Cao, G. Livingston, C. S. Carlson, N. J. Bass, C. M. Carlsson, J. Hardy, R. M. Carney, J. Bras, M. M. Carrasquillo, R. Guerreiro, M. Allen, H. C. Chui, E. Fisher, C. Masullo, E. A. Crocco, C. DeCarli, G. Bisceglio, M. Dick, L. Ma, R. Duara, N. R. Graff-Radford, D. A. Evans, A. Hodges, K. M. Faber, M. Scherer, K. B. Fallon, M. Riemenschneider, D. W. Fardo, R. Heun, M. R. Farlow, H. Kölsch, S. Ferris, M. Leber, T. M. Foroud, I. Heuser, D. R. Galasko, I. Giegling, M. Gearing, M. Hüll, D. H. Geschwind, J. R. Gilbert, J. Morris, R. C. Green, K. Mayo, J. H. Growdon, T. Feulner, R. L. Hamilton, L. E. Harrell, D. Drichel, L. S. Honig, T. D. Cushion, M. J. Huentelman, P. Hollingworth, C. M. Hulette, B. T. Hyman, R. Marshall, G. P. Jarvik, A. Meggy, E. Abner, G. E. Menzies, L.-W. Jin, G. Leonenko, L. M. Real, G. R. Jun, C. T. Baldwin, D. Grozeva, A. Karydas, G. Russo, J. A. Kaye, R. Kim, F. Jessen, N. W. Kowall, B. Vellas, J. H. Kramer, E. Vardy, F. M. LaFerla, K.-H. Jöckel, J. J. Lah, M. Dichgans, J. B. Leverenz, D. Mann, A. I. Levey, S. Pickering-Brown, A. P. Lieberman, N. Klopp, K. L. Lunetta, H.-E. Wichmann, C. G. Lyketsos, K. Morgan, D. C. Marson, K. Brown, F. Martiniuk, C. Medway, D. C. Mash, M. M. Nöthen, E. Masliah, N. M. Hooper, W. C. McCormick, A. Daniele, S. M. McCurry, A. Bayer, A. N. McDavid, J. Gallacher, A. C. McKee, H. van den Bussche, M. Mesulam, C. Brayne, B. L. Miller, S. Riedel-Heller, C. A. Miller, J. W. Miller, A. Al-Chalabi, J. C. Morris, C. E. Shaw, A. J. Myers, J. Wiltfang, S. O’Bryant, J. M. Olichney, V. Alvarez, J. E. Parisi, A. B. Singleton, H. L. Paulson, J. Collinge, W. R. Perry, S. Mead, E. Peskind, D. H. Cribbs, M. Rossor, A. Pierce, N. S. Ryan, W. W. Poon, B. Nacmias, H. Potter, S. Sorbi, J. F. Quinn, E. Sacchinelli, A. Raj, G. Spalletta, M. Raskind, C. Caltagirone, P. Bossù, M. D. Orfei, B. Reisberg, R. Clarke, C. Reitz, A. D. Smith, J. M. Ringman, D. Warden, E. D. Roberson, G. Wilcock, E. Rogaeva, A. C. Bruni, H. J. Rosen, M. Gallo, R. N. Rosenberg, Y. Ben-Shlomo, M. A. Sager, P. Mecocci, A. J. Saykin, P. Pastor, M. L. Cuccaro, J. M. Vance, J. A. Schneider, L. S. Schneider, S. Slifer, W. W. Seeley, A. G. Smith, J. A. Sonnen, S. Spina, R. A. Stern, R. H. Swerdlow, M. Tang, R. E. Tanzi, J. Q. Trojanowski, J. C. Troncoso, V. M. V. Deerlin, L. J. V. Eldik, H. V. Vinters, J. P. Vonsattel, S. Weintraub, K. A. Welsh-Bohmer, K. C. Wilhelmsen, J. Williamson, T. S. Wingo, R. L. Woltjer, C. B. Wright, C.-E. Yu, L. Yu, Y. Saba, A. Pilotto, M. J. Bullido, O. Peters, P. K. Crane, D. Bennett, P. Bosco, E. Coto, V. Boccardi, P. L. D. Jager, A. Lleo, N. Warner, O. L. Lopez, M. Ingelsson, P. Deloukas, C. Cruchaga, C. Graff, R. Gwilliam, M. Fornage, A. M. Goate, P. Sanchez-Juan, P. G. Kehoe, N. Amin, N. Ertekin-Taner, C. Berr, S. Debette, S. Love, L. J. Launer, S. G. Younkin, J.-F. Dartigues, C. Corcoran, M. A. Ikram, D. W. Dickson, G. Nicolas, D. Campion, J. Tschanz, H. Schmidt, H. Hakonarson, J. Clarimon, R. Munger, R. Schmidt, L. A. Farrer, C. V. Broeckhoven, M. C. O’Donovan, A. L. DeStefano, L. Jones, J. L. Haines, J.-F. Deleuze, M. J. Owen, V. Gudnason, R. Mayeux, V. Escott-Price, B. M. Psaty, A. Ramirez, L.-S. Wang, A. Ruiz, C. M. van Duijn, P. A. Holmans, S. Seshadri, J. Williams, P. Amouyel, G. D. Schellenberg, J.-C. Lambert, M. A. Pericak-Vance, Genetic meta-analysis of diagnosed Alzheimer’s disease identifies new risk loci and implicates Aβ, tau, immunity and lipid processing. Nat. Genet. 51, 414 (2019).

15. I. E. Jansen, J. E. Savage, K. Watanabe, J. Bryois, D. M. Williams, S. Steinberg, J. Sealock, I. K. Karlsson, S. Hägg, L. Athanasiu, N. Voyle, P. Proitsi, A. Witoelar, S. Stringer, D. Aarsland, I. S. Almdahl, F. Andersen, S. Bergh, F. Bettella, S. Bjornsson, A. Brækhus, G. Bråthen, C. de Leeuw, R. S. Desikan, S. Djurovic, L. Dumitrescu, T. Fladby, T. J. Hohman, P. V. Jonsson, S. J. Kiddle, A. Rongve, I. Saltvedt, S. B. Sando, G. Selbæk, M. Shoai, N. G. Skene, J. Snaedal, E. Stordal, I. D. Ulstein, Y. Wang, L. R. White, J. Hardy, J. Hjerling-Leffler, P. F. Sullivan, W. M. van der Flier, R. Dobson, L. K. Davis, H. Stefansson, K. Stefansson, N. L. Pedersen, S. Ripke, O. A. Andreassen, D. Posthuma, Genome-wide meta-analysis identifies new loci and functional pathways influencing Alzheimer’s disease risk. Nat. Genet. 51, 404–413 (2019).

16. C. Bellenguez, F. Küçükali, I. E. Jansen, L. Kleineidam, S. Moreno-Grau, N. Amin, A. C. Naj, R. Campos-Martin, B. Grenier-Boley, V. Andrade, P. A. Holmans, A. Boland, V. Damotte, S. J. van der Lee, M. R. Costa, T. Kuulasmaa, Q. Yang, I. de Rojas, J. C. Bis, A. Yaqub, I. Prokic, J. Chapuis, S. Ahmad, V. Giedraitis, D. Aarsland, P. Garcia-Gonzalez, C. Abdelnour, E. Alarcón-Martín, D. Alcolea, M. Alegret, I. Alvarez, V. Álvarez, N. J. Armstrong, A. Tsolaki, C. Antúnez, I. Appollonio, M. Arcaro, S. Archetti, A. A. Pastor, B. Arosio, L. Athanasiu, H. Bailly, N. Banaj, M. Baquero, S. Barral, A. Beiser, A. B. Pastor, J. E. Below, P. Benchek, L. Benussi, C. Berr, C. Besse, V. Bessi, G. Binetti, A. Bizarro, R. Blesa, M. Boada, E. Boerwinkle, B. Borroni, S. Boschi, P. Bossù, G. Bråthen, J. Bressler, C. Bresner, H. Brodaty, K. J. Brookes, L. I. Brusco, D. Buiza-Rueda, K. Bûrger, V. Burholt, W. S. Bush, M. Calero, L. B. Cantwell, G. Chene, J. Chung, M. L. Cuccaro, Á. Carracedo, R. Cecchetti, L. Cervera-Carles, C. Charbonnier, H.-H. Chen, C. Chillotti, S. Ciccone, J. A. H. R. Claassen, C. Clark, E. Conti, A. Corma-Gómez, E. Costantini, C. Custodero, D. Daian, M. C. Dalmasso, A. Daniele, E. Dardiotis, J.-F. Dartigues, P. P. de Deyn, K. de Paiva Lopes, L. D. de Witte, S. Debette, J. Deckert, T. del Ser, N. Denning, A. DeStefano, M. Dichgans, J. Diehl-Schmid, M. Diez-Fairen, P. D. Rossi, S. Djurovic, E. Duron, E. Düzel, C. Dufouil, G. Eiriksdottir, S. Engelborghs, V. Escott-Price, A. Espinosa, M. Ewers, K. M. Faber, T. Fabrizio, S. F. Nielsen, D. W. Fardo, L. Farotti, C. Fenoglio, M. Fernández-Fuertes, R. Ferrari, C. B. Ferreira, E. Ferri, B. Fin, P. Fischer, T. Fladby, K. Fließbach, B. Fongang, M. Fornage, J. Fortea, T. M. Foroud, S. Fostinelli, N. C. Fox, E. Franco-Macías, M. J. Bullido, A. Frank-García, L. Froelich, B. Fulton-Howard, D. Galimberti, J. M. García-Alberca, P. García-González, S. Garcia-Madrona, G. Garcia-Ribas, R. Ghidoni, I. Giegling, G. Giorgio, A. M. Goate, O. Goldhardt, D. Gomez-Fonseca, A. González-Pérez, C. Graff, G. Grande, E. Green, T. Grimmer, E. Grünblatt, M. Grunin, V. Gudnason, T. Guetta-Baranes, A. Haapasalo, G. Hadjigeorgiou, J. L. Haines, K. L. Hamilton-Nelson, H. Hampel, O. Hanon, J. Hardy, A. M. Hartmann, L. Hausner, J. Harwood, S. Heilmann-Heimbach, S. Helisalmi, M. T. Heneka, I. Hernández, M. J. Herrmann, P. Hoffmann, C. Holmes, H. Holstege, R. H. Vilas, M. Hulsman, J. Humphrey, G. J. Biessels, X. Jian, C. Johansson, G. R. Jun, Y. Kastumata, J. Kauwe, P. G. Kehoe, L. Kilander, A. K. Ståhlbom, M. Kivipelto, A. Koivisto, J. Kornhuber, M. H. Kosmidis, W. A. Kukull, P. P. Kuksa, B. W. Kunkle, A. B. Kuzma, C. Lage, E. J. Laukka, L. Launer, A. Lauria, C.-Y. Lee, J. Lehtisalo, O. Lerch, A. Lleó, W. Longstreth, O. Lopez, A. L. de Munain, S. Love, M. Löwemark, L. Luckcuck, K. L. Lunetta, Y. Ma, J. Macías, C. A. MacLeod, W. Maier, F. Mangialasche, M. Spallazzi, M. Marquié, R. Marshall, E. R. Martin, A. M. Montes, C. M. Rodríguez, C. Masullo, R. Mayeux, S. Mead, P. Mecocci, M. Medina, A. Meggy, S. Mehrabian, S. Mendoza, M. Menéndez-González, P. Mir, S. Moebus, M. Mol, L. Molina-Porcel, L. Montrreal, L. Morelli, F. Moreno, K. Morgan, T. Mosley, M. M. Nöthen, C. Muchnik, S. Mukherjee, B. Nacmias, T. Ngandu, G. Nicolas, B. G. Nordestgaard, R. Olaso, A. Orellana, M. Orsini, G. Ortega, A. Padovani, C. Paolo, G. Papenberg, L. Parnetti, F. Pasquier, P. Pastor, G. Peloso, A. Pérez-Cordón, J. Pérez-Tur, P. Pericard, O. Peters, Y. A. L. Pijnenburg, J. A. Pineda, G. Piñol-Ripoll, C. Pisanu, T. Polak, J. Popp, D. Posthuma, J. Priller, R. Puerta, O. Quenez, I. Quintela, J. Q. Thomassen, A. Rábano, I. Rainero, F. Rajabli, I. Ramakers, L. M. Real, M. J. T. Reinders, C. Reitz, D. Reyes-Dumeyer, P. Ridge, S. Riedel-Heller, P. Riederer, N. Roberto, E. Rodriguez-Rodriguez, A. Rongve, I. R. Allende, M. Rosende-Roca, J. L. Royo, E. Rubino, D. Rujescu, M. E. Sáez, P. Sakka, I. Saltvedt, Á. Sanabria, M. B. Sánchez-Arjona, F. Sanchez-Garcia, P. S. Juan, R. Sánchez-Valle, S. B. Sando, C. Sarnowski, C. L. Satizabal, M. Scamosci, N. Scarmeas, E. Scarpini, P. Scheltens, N. Scherbaum, M. Scherer, M. Schmid, A. Schneider, J. M. Schott, G. Selbæk, D. Seripa, M. Serrano, J. Sha, A. A. Shadrin, O. Skrobot, S. Slifer, G. J. L. Snijders, H. Soininen, V. Solfrizzi, A. Solomon, Y. Song, S. Sorbi, O. Sotolongo-Grau, G. Spalletta, A. Spottke, A. Squassina, E. Stordal, J. P. Tartan, L. Tárraga, N. Tesí, A. Thalamuthu, T. Thomas, G. Tosto, L. Traykov, L. Tremolizzo, A. Tybjærg-Hansen, A. Uitterlinden, A. Ullgren, I. Ulstein, S. Valero, O. Valladares, C. V. Broeckhoven, J. Vance, B. N. Vardarajan, A. van der Lugt, J. V. Dongen, J. van Rooij, J. van Swieten, R. Vandenberghe, F. Verhey, J.-S. Vidal, J. Vogelgsang, M. Vyhnalek, M. Wagner, D. Wallon, L.-S. Wang, R. Wang, L. Weinhold, J. Wiltfang, G. Windle, B. Woods, M. Yannakoulia, H. Zare, Y. Zhao, X. Zhang, C. Zhu, M. Zulaica, L. A. Farrer, B. M. Psaty, M. Ghanbari, T. Raj, P. Sachdev, K. Mather, F. Jessen, M. A. Ikram, A. de Mendonça, J. Hort, M. Tsolaki, M. A. Pericak-Vance, P. Amouyel, J. Williams, R. Frikke-Schmidt, J. Clarimon, J.-F. Deleuze, G. Rossi, S. Seshadri, O. A. Andreassen, M. Ingelsson, M. Hiltunen, K. Sleegers, G. D. Schellenberg, C. M. van Duijn, R. Sims, W. M. van der Flier, A. Ruiz, A. Ramirez, J.-C. Lambert, New insights into the genetic etiology of Alzheimer’s disease and related dementias. Nat. Genet., 1–25 (2022).

17. R. Kaddurah-Daouk, S. Rozen, W. Matson, X. Han, C. M. Hulette, J. R. Burke, P. M. Doraiswamy, K. A. Welsh-Bohmer, Metabolomic changes in autopsy-confirmed Alzheimer’s disease. Alzheimers Dement. 7, 309–317 (2011).

18. E. Trushina, T. Dutta, X.-M. T. Persson, M. M. Mielke, R. C. Petersen, Identification of Altered Metabolic Pathways in Plasma and CSF in Mild Cognitive Impairment and Alzheimer’s Disease Using Metabolomics. PLOS ONE 8, e63644 (2013).

19. P. E. Khoonsari, A. Häggmark, M. Lönnberg, M. Mikus, L. Kilander, L. Lannfelt, J. Bergquist, M. Ingelsson, P. Nilsson, K. Kultima, G. Shevchenko, Analysis of the Cerebrospinal Fluid Proteome in Alzheimer’s Disease. PLOS ONE 11, e0150672 (2016).

20. R. W. Paterson, W. E. Heywood, A. J. Heslegrave, N. K. Magdalinou, U. Andreasson, E. Sirka, E. Bliss, C. F. Slattery, J. Toombs, J. Svensson, P. Johansson, N. C. Fox, H. Zetterberg, K. Mills, J. M. Schott, A targeted proteomic multiplex CSF assay identifies increased malate dehydrogenase and other neurodegenerative biomarkers in individuals with Alzheimer’s disease pathology. Transl. Psychiatry 6, e952 (2016).

21. D. S. Spellman, K. R. Wildsmith, L. A. Honigberg, M. Tuefferd, D. Baker, N. Raghavan, A. C. Nairn, P. Croteau, M. Schirm, R. Allard, J. Lamontagne, D. Chelsky, S. Hoffmann, W. Z. Potter, Development and Evaluation of a Multiplexed Mass Spectrometry-Based Assay for Measuring Candidate Peptide Biomarkers in Alzheimer’s Disease Neuroimaging Initiative (ADNI) CSF. Proteomics Clin. Appl. 9, 715–731 (2015).

22. K. R. Wildsmith, S. P. Schauer, A. M. Smith, D. Arnott, Y. Zhu, J. Haznedar, S. Kaur, W. R. Mathews, L. A. Honigberg, Identification of longitudinally dynamic biomarkers in Alzheimer’s disease cerebrospinal fluid by targeted proteomics. Mol. Neurodegener. 9, 22 (2014).

23. C. D. Whelan, N. Mattsson, M. W. Nagle, S. Vijayaraghavan, C. Hyde, S. Janelidze, E. Stomrud, J. Lee, L. Fitz, T. A. Samad, G. Ramaswamy, R. A. Margolin, A. Malarstig, O. Hansson, Multiplex proteomics identifies novel CSF and plasma biomarkers of early Alzheimer’s disease. Acta Neuropathol. Commun. 7, 169 (2019).

24. E. C. B. Johnson, E. B. Dammer, D. M. Duong, L. Ping, M. Zhou, L. Yin, L. A. Higginbotham, A. Guajardo, B. White, J. C. Troncoso, M. Thambisetty, T. J. Montine, E. B. Lee, J. Q. Trojanowski, T. G. Beach, E. M. Reiman, V. Haroutunian, M. Wang, E. Schadt, B. Zhang, D. W. Dickson, N. Ertekin-Taner, T. E. Golde, V. A. Petyuk, P. L. De Jager, D. A. Bennett, T. S. Wingo, S. Rangaraju, I. Hajjar, J. M. Shulman, J. J. Lah, A. I. Levey, N. T. Seyfried, Large-scale proteomic analysis of Alzheimer’s disease brain and cerebrospinal fluid reveals early changes in energy metabolism associated with microglia and astrocyte activation. Nat. Med. 26, 769–780 (2020).

25. J. McKetney, D. J. Panyard, S. C. Johnson, C. Carlsson, C. D. Engelman, J. J. Coon, Pilot proteomic analysis of cerebrospinal fluid in Alzheimer’s disease. Proteomics Clin. Appl., e2000072 (2021).

26. L. Higginbotham, L. Ping, E. B. Dammer, D. M. Duong, M. Zhou, M. Gearing, C. Hurst, J. D. Glass, S. A. Factor, E. C. B. Johnson, I. Hajjar, J. J. Lah, A. I. Levey, N. T. Seyfried, Integrated proteomics reveals brain-based cerebrospinal fluid biomarkers in asymptomatic and symptomatic Alzheimer’s disease. Sci. Adv. 6, eaaz9360 (2020).

27. S. Pan, D. Zhu, J. F. Quinn, E. R. Peskind, T. J. Montine, B. Lin, D. R. Goodlett, G. Taylor, J. Eng, J. Zhang, A combined dataset of human cerebrospinal fluid proteins identified by multi-dimensional chromatography and tandem mass spectrometry. PROTEOMICS 7, 469–473 (2007).

28. A. Zougman, B. Pilch, A. Podtelejnikov, M. Kiehntopf, C. Schnabel, C. Kumar, M. Mann, Integrated Analysis of the Cerebrospinal Fluid Peptidome and Proteome. J. Proteome Res. 7, 386–399 (2008).

29. S. E. Schutzer, T. Liu, B. H. Natelson, T. E. Angel, A. A. Schepmoes, S. O. Purvine, K. K. Hixson, M. S. Lipton, D. G. Camp, P. K. Coyle, R. D. Smith, J. Bergquist, Establishing the Proteome of Normal Human Cerebrospinal Fluid. PLoS ONE 5 (2010), doi:10.1371/journal.pone.0010980.

30. Y. Zhang, Z. Guo, L. Zou, Y. Yang, L. Zhang, N. Ji, C. Shao, W. Sun, Y. Wang, A comprehensive map and functional annotation of the normal human cerebrospinal fluid proteome. J. Proteomics 119, 90–99 (2015).

31. I. Begcevic, D. Brinc, A. P. Drabovich, I. Batruch, E. P. Diamandis, Identification of brain-enriched proteins in the cerebrospinal fluid proteome by LC-MS/MS profiling and mining of the Human Protein Atlas. Clin. Proteomics 13 (2016), doi:10.1186/s12014-016-9111-3.

32. C. Macron, R. Lavigne, A. Núñez Galindo, M. Affolter, C. Pineau, L. Dayon, Exploration of human cerebrospinal fluid: A large proteome dataset revealed by trapped ion mobility time-of-flight mass spectrometry. Data Brief 31 (2020), doi:10.1016/j.dib.2020.105704.

33. H. Wang, K. K. Dey, P.-C. Chen, Y. Li, M. Niu, J.-H. Cho, X. Wang, B. Bai, Y. Jiao, S. R. Chepyala, V. Haroutunian, B. Zhang, T. G. Beach, J. Peng, Integrated analysis of ultra-deep proteomes in cortex, cerebrospinal fluid and serum reveals a mitochondrial signature in Alzheimer’s disease. Mol. Neurodegener. 15, 43 (2020).

34. L. Dayon, A. Núñez Galindo, J. Wojcik, O. Cominetti, J. Corthésy, A. Oikonomidi, H. Henry, M. Kussmann, E. Migliavacca, I. Severin, G. L. Bowman, J. Popp, Alzheimer disease pathology and the cerebrospinal fluid proteome. Alzheimers Res. Ther. 10, 66 (2018).

35. B. Bai, X. Wang, Y. Li, P.-C. Chen, K. Yu, K. K. Dey, J. M. Yarbro, X. Han, B. M. Lutz, S. Rao, Y. Jiao, J. M. Sifford, J. Han, M. Wang, H. Tan, T. I. Shaw, J.-H. Cho, S. Zhou, H. Wang, M. Niu, A. Mancieri, K. A. Messler, X. Sun, Z. Wu, V. Pagala, A. A. High, W. Bi, H. Zhang, H. Chi, V. Haroutunian, B. Zhang, T. G. Beach, G. Yu, J. Peng, Deep Multilayer Brain Proteomics Identifies Molecular Networks in Alzheimer’s Disease Progression. Neuron 105, 975–991.e7 (2020).

36. M. A. Korolainen, T. A. Nyman, T. Aittokallio, T. Pirttilä, An update on clinical proteomics in Alzheimer’s research. J. Neurochem. 112, 1386–1414 (2010).

37. E. Portelius, G. Brinkmalm, J. Pannee, H. Zetterberg, K. Blennow, R. Dahlén, A. Brinkmalm, J. Gobom, Proteomic studies of cerebrospinal fluid biomarkers of Alzheimer’s disease: an update. Expert Rev. Proteomics 14, 1007–1020 (2017).

38. N. A. O’Leary, M. W. Wright, J. R. Brister, S. Ciufo, D. Haddad, R. McVeigh, B. Rajput, B. Robbertse, B. Smith-White, D. Ako-Adjei, A. Astashyn, A. Badretdin, Y. Bao, O. Blinkova, V. Brover, V. Chetvernin, J. Choi, E. Cox, O. Ermolaeva, C. M. Farrell, T. Goldfarb, T. Gupta, D. Haft, E. Hatcher, W. Hlavina, V. S. Joardar, V. K. Kodali, W. Li, D. Maglott, P. Masterson, K. M. McGarvey, M. R. Murphy, K. O’Neill, S. Pujar, S. H. Rangwala, D. Rausch, L. D. Riddick, C. Schoch, A. Shkeda, S. S. Storz, H. Sun, F. Thibaud-Nissen, I. Tolstoy, R. E. Tully, A. R. Vatsan, C. Wallin, D. Webb, W. Wu, M. J. Landrum, A. Kimchi, T. Tatusova, M. DiCuccio, P. Kitts, T. D. Murphy, K. D. Pruitt, Reference sequence (RefSeq) database at NCBI: current status, taxonomic expansion, and functional annotation. Nucleic Acids Res. 44, D733–D745 (2016).

39. M. Comabella, M. Fernández, R. Martin, S. Rivera-Vallvé, E. Borrás, C. Chiva, E. Julià, A. Rovira, E. Cantó, J. C. Alvarez-Cermeño, L. M. Villar, M. Tintoré, X. Montalban, Cerebrospinal fluid chitinase 3-like 1 levels are associated with conversion to multiple sclerosis. Brain 133, 1082–1093 (2010).

40. L. J. Britton, K. Bridle, L.-A. Jaskowski, J. He, C. Ng, J. E. Ruelcke, A. Mohamed, J. Reiling, N. Santrampurwala, M. M. Hill, J. P. Whitehead, V. N. Subramaniam, D. H. G. Crawford, Iron Inhibits the Secretion of Apolipoprotein E in Cultured Human Adipocytes. Cell. Mol. Gastroenterol. Hepatol. 6, 215–217.e8 (2018).

41. D. Herebian, B. Alhaddad, A. Seibt, T. Schwarzmayr, K. Danhauser, D. Klee, S. Harmsen, T. Meitinger, T. M. Strom, A. Schulz, E. Mayatepek, T. B. Haack, F. Distelmaier, Coexisting variants in OSTM1 and MANEAL cause a complex neurodegenerative disorder with NBIA-like brain abnormalities. Eur. J. Hum. Genet. 25, 1092–1095 (2017).

42. N. N. Nalivaeva, N. D. Belyaev, I. A. Zhuravin, A. J. Turner, The Alzheimer’s Amyloid-Degrading Peptidase, Neprilysin: Can We Control It? Int. J. Alzheimers Dis. 2012, e383796 (2012).

43. O. Goldhardt, I. Warnhoff, I. Yakushev, I. Begcevic, H. Förstl, V. Magdolen, A. Soosaipillai, E. Diamandis, P. Alexopoulos, T. Grimmer, Kallikrein-related peptidases 6 and 10 are elevated in cerebrospinal fluid of patients with Alzheimer’s disease and associated with CSF-TAU and FDG-PET. Transl. Neurodegener. 8, 25 (2019).

44. B. M. Tijms, J. Gobom, L. Reus, I. Jansen, S. Hong, V. Dobricic, F. Kilpert, M. ten Kate, F. Barkhof, M. Tsolaki, F. R. J. Verhey, J. Popp, P. Martinez-Lage, R. Vandenberghe, A. Lleó, J. L. Molinuevo, S. Engelborghs, L. Bertram, S. Lovestone, J. Streffer, S. Vos, I. Bos, The Alzheimer’s Disease Neuroimaging Initiative (ADNI), K. Blennow, P. Scheltens, C. E. Teunissen, H. Zetterberg, P. J. Visser, Pathophysiological subtypes of Alzheimer’s disease based on cerebrospinal fluid proteomics. Brain 143, 3776–3792 (2020).

45. J. Remnestål, S. Bergström, J. Olofsson, E. Sjöstedt, M. Uhlén, K. Blennow, H. Zetterberg, A. Zettergren, S. Kern, I. Skoog, P. Nilsson, A. Månberg, Association of CSF proteins with tau and amyloid β levels in asymptomatic 70-year-olds. Alzheimers Res. Ther. 13, 54 (2021).

46. O. Hansson, S. Lehmann, M. Otto, H. Zetterberg, P. Lewczuk, Advantages and disadvantages of the use of the CSF Amyloid β (Aβ) 42/40 ratio in the diagnosis of Alzheimer’s Disease. Alzheimers Res. Ther. 11, 34 (2019).

47. A. Ng, W. W. Tam, M. W. Zhang, C. S. Ho, S. F. Husain, R. S. McIntyre, R. C. Ho, IL-1β, IL-6, TNF-α and CRP in Elderly Patients with Depression or Alzheimer’s disease: Systematic Review and Meta-Analysis. Sci. Rep. 8, 12050 (2018).

48. K.-C. Sonntag, W.-I. Ryu, K. M. Amirault, R. A. Healy, A. J. Siegel, D. L. McPhie, B. Forester, B. M. Cohen, Late-onset Alzheimer’s disease is associated with inherent changes in bioenergetics profiles. Sci. Rep. 7, 1–13 (2017).

49. S. Canchi, B. Raao, D. Masliah, S. B. Rosenthal, R. Sasik, K. M. Fisch, P. L. De Jager, D. A. Bennett, R. A. Rissman, Integrating Gene and Protein Expression Reveals Perturbed Functional Networks in Alzheimer’s Disease. Cell Rep. 28, 1103–1116.e4 (2019).

50. W.-I. Ryu, M. K. Bormann, M. Shen, D. Kim, B. Forester, Y. Park, J. So, H. Seo, K.-C. Sonntag, B. M. Cohen, Brain cells derived from Alzheimer’s disease patients have multiple specific innate abnormalities in energy metabolism. Mol. Psychiatry, 1–13 (2021).

51. M. Berger, M. Cooter, A. S. Roesler, S. Chung, J. Park, J. L. Modliszewski, K. W. VanDusen, J. W. Thompson, A. Moseley, M. J. Devinney, S. Smani, A. Hall, V. Cai, J. N. Browndyke, M. W. Lutz, D. L. Corcoran, and A. D. N. Initiative, APOE4 Copy Number-Dependent Proteomic Changes in the Cerebrospinal Fluid. J. Alzheimers Dis. 79, 511–530 (2021).

52. D. A. Butterfield, B. Halliwell, Oxidative stress, dysfunctional glucose metabolism and Alzheimer disease. Nat. Rev. Neurosci. 20, 148–160 (2019).

53. D. Bosco, A. Fava, M. Plastino, T. Montalcini, A. Pujia, Possible implications of insulin resistance and glucose metabolism in Alzheimer’s disease pathogenesis. J. Cell. Mol. Med. 15, 1807–1821 (2011).

54. E. M. Reiman, R. J. Caselli, L. S. Yun, K. Chen, D. Bandy, S. Minoshima, S. N. Thibodeau, D. Osborne, Preclinical evidence of Alzheimer’s disease in persons homozygous for the epsilon 4 allele for apolipoprotein E. N. Engl. J. Med. 334, 752–758 (1996).

55. E. M. Reiman, K. Chen, G. E. Alexander, R. J. Caselli, D. Bandy, D. Osborne, A. M. Saunders, J. Hardy, Functional brain abnormalities in young adults at genetic risk for late-onset Alzheimer’s dementia. Proc. Natl. Acad. Sci. U. S. A. 101, 284–289 (2004).

56. M. Perkins, A. B. Wolf, B. Chavira, D. Shonebarger, J. P. Meckel, L. Leung, L. Ballina, S. Ly, A. Saini, T. Jones, J. Vallejo, G. Jentarra, J. Valla, Altered Energy Metabolism Pathways in the Posterior Cingulate in Young Adult Apolipoprotein E ɛ4 Carriers. J. Alzheimers Dis. JAD 53, 95–106 (2016).

57. E. Iori, R. Millioni, L. Puricelli, G. Arrigoni, L. Lenzini, R. Trevisan, P. James, G. P. Rossi, L. A. Pinna, P. Tessari, Glycolytic enzyme expression and pyruvate kinase activity in cultured fibroblasts from type 1 diabetic patients with and without nephropathy. Biochim. Biophys. Acta BBA - Mol. Basis Dis. 1782, 627– 633 (2008).

58. S. E. Arnold, Z. Arvanitakis, S. L. Macauley-Rambach, A. M. Koenig, H.-Y. Wang, R. S. Ahima, S. Craft, S. Gandy, C. Buettner, L. E. Stoeckel, D. M. Holtzman, D. M. Nathan, Brain insulin resistance in type 2 diabetes and Alzheimer disease: concepts and conundrums. Nat. Rev. Neurol. 14, 168–181 (2018).

59. E. Steen, B. M. Terry, E. J. Rivera, J. L. Cannon, T. R. Neely, R. Tavares, X. J. Xu, J. R. Wands, S. M. de la Monte, Impaired insulin and insulin-like growth factor expression and signaling mechanisms in Alzheimer’s disease – is this type 3 diabetes? J. Alzheimers Dis. 7, 63–80 (2005).

60. E. J. Rivera, A. Goldin, N. Fulmer, R. Tavares, J. R. Wands, S. M. de la Monte, Insulin and insulin-like growth factor expression and function deteriorate with progression of Alzheimer’s disease: Link to brain reductions in acetylcholine. J. Alzheimers Dis. 8, 247–268 (2005).

61. K. Talbot, H.-Y. Wang, H. Kazi, L.-Y. Han, K. P. Bakshi, A. Stucky, R. L. Fuino, K. R. Kawaguchi, A. J. Samoyedny, R. S. Wilson, Z. Arvanitakis, J. A. Schneider, B. A. Wolf, D. A. Bennett, J. Q. Trojanowski, S. E. Arnold, Demonstrated brain insulin resistance in Alzheimer’s disease patients is associated with IGF-1 resistance, IRS-1 dysregulation, and cognitive decline. J. Clin. Invest. 122, 1316–1338 (2012).

62. H. Y. Agbemenyah, R. C. Agis-Balboa, S. Burkhardt, I. Delalle, A. Fischer, Insulin growth factor binding protein 7 is a novel target to treat dementia. Neurobiol. Dis. 62, 135–143 (2014).

63. D. S. Wishart, Y. D. Feunang, A. Marcu, A. C. Guo, K. Liang, R. Vázquez-Fresno, T. Sajed, D. Johnson, C. Li, N. Karu, Z. Sayeeda, E. Lo, N. Assempour, M. Berjanskii, S. Singhal, D. Arndt, Y. Liang, H. Badran, J. Grant, A. Serra-Cayuela, Y. Liu, R. Mandal, V. Neveu, A. Pon, C. Knox, M. Wilson, C. Manach, A. Scalbert, HMDB 4.0: the human metabolome database for 2018. Nucleic Acids Res. 46, D608–D617 (2018).

64. S. Trefely, C. D. Lovell, N. W. Snyder, K. E. Wellen, Compartmentalised acyl-CoA metabolism and roles in chromatin regulation. Mol. Metab. 38, 100941 (2020).

65. Y. Dong, G. J. Brewer, Global Metabolic Shifts in Age and Alzheimer’s Disease Mouse Brains Pivot at NAD+/NADH Redox Sites. J. Alzheimers Dis. 71, 119–140 (2019).

66. A. Ramirez, W. M. van der Flier, C. Herold, D. Ramonet, S. Heilmann, P. Lewczuk, J. Popp, A. Lacour, D. Drichel, E. Louwersheimer, M. P. Kummer, C. Cruchaga, P. Hoffmann, C. Teunissen, H. Holstege, J. Kornhuber, O. Peters, A. C. Naj, V. Chouraki, C. Bellenguez, A. Gerrish, International Genomics of Alzheimer’s Project (IGAP), Alzheimer’s Disease Neuroimaging Initiative (ADNI), R. Heun, L. Frölich, M. Hüll, L. Buscemi, S. Herms, H. Kölsch, P. Scheltens, M. M. Breteler, E. Rüther, J. Wiltfang, A. Goate, F. Jessen, W. Maier, M. T. Heneka, T. Becker, M. M. Nöthen, SUCLG2 identified as both a determinator of CSF Aβ1-42 levels and an attenuator of cognitive decline in Alzheimer’s disease. Hum. Mol. Genet. 23, 6644–6658 (2014).

67. E. Horgusluoglu, R. Neff, W.-M. Song, M. Wang, Q. Wang, M. Arnold, J. Krumsiek, B. Galindo-Prieto, Ming, K. Nho, G. Kastenmüller, X. Han, R. Baillie, Q. Zeng, S. Andrews, H. Cheng, K. Hao, A. Goate, D. A. Bennett, A. J. Saykin, R. Kaddurah-Daouk, B. Zhang, for the A. D. N. Initiative (ADNI), the A. D. M. Consortium, Integrative metabolomics-genomics approach reveals key metabolic pathways and regulators of Alzheimer’s disease. Alzheimers Dement. **n/a** (2021), doi:10.1002/alz.12468.

68. S. H. Lee, S. Park, H.-S. Kim, B. H. Jung, Metabolomic approaches to the normal aging process. Metabolomics 10, 1268–1292 (2014).

69. Z. Yu, G. Zhai, P. Singmann, Y. He, T. Xu, C. Prehn, W. Römisch-Margl, E. Lattka, C. Gieger, N. Soranzo, J. Heinrich, M. Standl, E. Thiering, K. Mittelstraß, H.-E. Wichmann, A. Peters, K. Suhre, Y. Li, J. Adamski, T. Spector, T. Illig, R. Wang-Sattler, Human serum metabolic profiles are age dependent. Aging Cell 11, 960–967 (2012).

70. T. Morawe, C. Hiebel, A. Kern, C. Behl, Protein Homeostasis, Aging and Alzheimer’s Disease. Mol. Neurobiol. 46, 41–54 (2012).

71. F. Chiti, C. M. Dobson, Protein Misfolding, Amyloid Formation, and Human Disease: A Summary of Progress Over the Last Decade. Annu. Rev. Biochem. 86, 27–68 (2017).

72. A. Höhn, A. Tramutola, R. Cascella, Proteostasis Failure in Neurodegenerative Diseases: Focus on Oxidative Stress. Oxid. Med. Cell. Longev. 2020 (2020), doi:10.1155/2020/5497046.

73. B. P. Festa, A. D. Barbosa, M. Rob, D. C. Rubinsztein, The pleiotropic roles of autophagy in Alzheimer’s disease: From pathophysiology to therapy. Curr. Opin. Pharmacol. 60, 149–157 (2021).

74. D. A. Mahajan, M. J. Go, W. Zhang, J. E. Below, K. J. Gaulton, T. Ferreira, M. Horikoshi, A. D. Johnson, M. C. Y. Ng, I. Prokopenko, D. Saleheen, X. Wang, E. Zeggini, G. R. Abecasis, L. S. Adair, P. Almgren, M. Atalay, T. Aung, D. Baldassarre, B. Balkau, Y. Bao, A. H. Barnett, I. Barroso, A. Basit, L. F. Been, J. Beilby, G. I. Bell, R. Benediktsson, R. N. Bergman, B. O. Boehm, E. Boerwinkle, L. L. Bonnycastle, N. Burtt, Q. Cai, H. Campbell, J. Carey, S. Cauchi, M. Caulfield, J. C. N. Chan, L.-C. Chang, T.-J. Chang, Y.-C. Chang, G. Charpentier, C.-H. Chen, H. Chen, Y.-T. Chen, K.-S. Chia, M. Chidambaram, P. S. Chines, N. H. Cho, Y. M. Cho, L.-M. Chuang, F. S. Collins, M. C. Cornelis, D. J. Couper, A. T. Crenshaw, R. M. van Dam, J. Danesh, D. Das, U. de Faire, G. Dedoussis, P. Deloukas, A. S. Dimas, C. Dina, A. S. F. Doney, P. J. Donnelly, M. Dorkhan, C. van Duijn, J. Dupuis, S. Edkins, P. Elliott, V. Emilsson, R. Erbel, J. G. Eriksson, J. Escobedo, T. Esko, E. Eury, J. C. Florez, P. Fontanillas, N. G. Forouhi, T. Forsen, C. Fox, R. M. Fraser, T. M. Frayling, P. Froguel, P. Frossard, Y. Gao, K. Gertow, C. Gieger, B. Gigante, H. Grallert, G. B. Grant, L. C. Groop, C. J. Groves, E. Grundberg, C. Guiducci, A. Hamsten, B.-G. Han, K. Hara, N. Hassanali, A. T. Hattersley, C. Hayward, A. K. Hedman, C. Herder, A. Hofman, O. L. Holmen, K. Hovingh, A. B. Hreidarsson, C. Hu, F. B. Hu, J. Hui, S. E. Humphries, S. E. Hunt, D. J. Hunter, K. Hveem, Z. I. Hydrie, H. Ikegami, T. Illig, E. Ingelsson, M. Islam, B. Isomaa, A. U. Jackson, T. Jafar, A. James, W. Jia, K.-H. Jöckel, A. Jonsson, J. B. M. Jowett, T. Kadowaki, H. M. Kang, S. Kanoni, W. H. L. Kao, S. Kathiresan, N. Kato, P. Katulanda, S. M. Keinanen-Kiukaanniemi, A. M. Kelly, H. Khan, K.-T. Khaw, C.-C. Khor, H.-L. Kim, S. Kim, Y. J. Kim, L. Kinnunen, N. Klopp, A. Kong, E. Korpi-Hyövälti, S. Kowlessur, P. Kraft, J. Kravic, M. M. Kristensen, S. Krithika, A. Kumar, J. Kumate, J. Kuusisto, S. H. Kwak, M. Laakso, V. Lagou, T. A. Lakka, C. Langenberg, C. Langford, R. Lawrence, K. Leander, J.-M. Lee, N. R. Lee, M. Li, X. Li, Y. Li, J. Liang, S. Liju, W.-Y. Lim, L. Lind, C. M. Lindgren, E. Lindholm, C.-T. Liu, J. J. Liu, S. Lobbens, J. Long, R. J. F. Loos, W. Lu, J. Luan, V. Lyssenko, R. C. W. Ma, S. Maeda, R. Mägi, S. Männistö, D. R. Matthews, J. B. Meigs, O. Melander, A. Metspalu, J. Meyer, G. Mirza, E. Mihailov, S. Moebus, V. Mohan, K. L. Mohlke, A. D. Morris, T. W. Mühleisen, M. Müller-Nurasyid, B. Musk, J. Nakamura, E. Nakashima, P. Navarro, P.-K. Ng, A. C. Nica, P. M. Nilsson, I. Njølstad, M. M. Nöthen, K. Ohnaka, T. H. Ong, K. R. Owen, C. N. A. Palmer, J. S. Pankow, K. S. Park, M. Parkin, S. Pechlivanis, N. L. Pedersen, L. Peltonen, J. R. B. Perry, A. Peters, J. M. Pinidiyapathirage, C. G. P. Platou, S. Potter, J. F. Price, L. Qi, V. Radha, L. Rallidis, A. Rasheed, W. Rathmann, R. Rauramaa, S. Raychaudhuri, N. W. Rayner, S. D. Rees, E. Rehnberg, S. Ripatti, N. Robertson, M. Roden, E. J. Rossin, I. Rudan, D. Rybin, T. Saaristo, V. Salomaa, J. Saltevo, M. Samuel, D. K. Sanghera, J. Saramies, J. Scott, L. J. Scott, R. A. Scott, A. V. Segrè, J. Sehmi, B. Sennblad, N. Shah, S. Shah, A. S. Shera, X. O. Shu, A. R. Shuldiner, G. Sigurðsson, E. Sijbrands, A. Silveira, X. Sim, S. Sivapalaratnam, K. S. Small, W. Y. So, A. Stančáková, K. Stefansson, G. Steinbach, V. Steinthorsdottir, K. Stirrups, R. J. Strawbridge, H. M. Stringham, Q. Sun, C. Suo, A.-C. Syvänen, R. Takayanagi, F. Takeuchi, W. T. Tay, T. M. Teslovich, B. Thorand, G. Thorleifsson, U. Thorsteinsdottir, E. Tikkanen, J. Trakalo, E. Tremoli, M. D. Trip, F. J. Tsai, T. Tuomi, J. Tuomilehto, A. G. Uitterlinden, A. Valladares-Salgado, S. Vedantam, F. Veglia, B. F. Voight, C. Wang, N. J. Wareham, R. Wennauer, A. R. Wickremasinghe, T. Wilsgaard, J. F. Wilson, S. Wiltshire, W. Winckler, T. Y. Wong, A. R. Wood, J.-Y. Wu, Y. Wu, K. Yamamoto, T. Yamauchi, M. Yang, L. Yengo, M. Yokota, R. Young, D. Zabaneh, F. Zhang, R. Zhang, W. Zheng, P. Z. Zimmet, D. Altshuler, D. W. Bowden, Y. S. Cho, N. J. Cox, M. Cruz, C. L. Hanis, J. Kooner, J.-Y. Lee, M. Seielstad, Y. Y. Teo, M. Boehnke, E. J. Parra, J. C. Chambers, E. S. Tai, M. I. McCarthy, A. P. Morris, Genome-wide trans-ancestry meta-analysis provides insight into the genetic architecture of type 2 diabetes susceptibility. Nat. Genet. 46, 234–244 (2014).

75. Q. Zhang, C. Ma, M. Gearing, P. G. Wang, L.-S. Chin, L. Li, Integrated proteomics and network analysis identifies protein hubs and network alterations in Alzheimer’s disease. Acta Neuropathol. Commun. 6 (2018), doi:10.1186/s40478-018-0524-2.

76. P. E. Khoonsari, G. Shevchenko, S. Herman, J. Remnestål, V. Giedraitis, R. Brundin, M. Degerman Gunnarsson, L. Kilander, H. Zetterberg, P. Nilsson, L. Lannfelt, M. Ingelsson, K. Kultima, Improved Differential Diagnosis of Alzheimer’s Disease by Integrating ELISA and Mass Spectrometry-Based Cerebrospinal Fluid Biomarkers. J. Alzheimers Dis. 67, 639–651 (2019).

77. L. M. Raffield, H. Dang, K. A. Pratte, S. Jacobson, L. Gillenwater, E. Ampleford, I. Barjaktarevic, P. Basta, C. B. Clish, A. P. Comellas, E. Cornell, J. L. Curtis, C. Doerschuk, P. Durda, C. Emson, C. Freeman, X. Guo, A. T. Hastie, G. A. Hawkins, J. Herrera, W. C. Johnson, W. W. Labaki, Y. Liu, B. Masters, M. Miller, V. E. Ortega, G. Papanicolaou, S. Peters, K. D. Taylor, S. S. Rich, J. I. Rotter, P. Auer, A. P. Reiner, R. P. Tracy, D. Ngo, R. E. Gerszten, W. K. O’Neal, R. P. Bowler, Comparison of Proteomic Assessment Methods in Multiple Cohort Studies. Proteomics 20, e1900278 (2020).

78. M. Pietzner, E. Wheeler, J. Carrasco-Zanini, N. D. Kerrison, E. Oerton, M. Koprulu, J. Luan, A. D. Hingorani, S. A. Williams, N. J. Wareham, C. Langenberg, Synergistic insights into human health from aptamer- and antibody-based proteomic profiling. Nat. Commun. 12, 6822 (2021).

79. S. C. Johnson, R. L. Koscik, E. M. Jonaitis, L. R. Clark, K. D. Mueller, S. E. Berman, B. B. Bendlin, C. D. Engelman, O. C. Okonkwo, K. J. Hogan, S. Asthana, C. M. Carlsson, B. P. Hermann, M. A. Sager, The Wisconsin Registry for Alzheimer’s Prevention: A review of findings and current directions. Alzheimers Dement. Diagn. Assess. Dis. Monit. 10, 130–142 (2017).

80. K. E. Melah, S. Y.-F. Lu, S. M. Hoscheidt, A. L. Alexander, N. Adluru, D. J. Destiche, C. M. Carlsson, H. Zetterberg, K. Blennow, O. C. Okonkwo, C. E. Gleason, N. M. Dowling, L. C. Bratzke, H. A. Rowley, M. A. Sager, S. Asthana, S. C. Johnson, B. B. Bendlin, CSF markers of Alzheimer’s pathology and microglial activation are associated with altered white matter microstructure in asymptomatic adults at risk for Alzheimer’s disease. J. Alzheimers Dis. 50, 873–886 (2016).

81. E. von Elm, D. G. Altman, M. Egger, S. J. Pocock, P. C. Gøtzsche, J. P. Vandenbroucke, STROBE Initiative, The Strengthening the Reporting of Observational Studies in Epidemiology (STROBE) statement: guidelines for reporting observational studies. J. Clin. Epidemiol. 61, 344–349 (2008).

82. C. A. Van Hulle, E. M. Jonaitis, T. J. Betthauser, R. Batrla, N. Wild, G. Kollmorgen, U. Andreasson, O. Okonkwo, B. B. Bendlin, S. Asthana, C. M. Carlsson, S. C. Johnson, H. Zetterberg, K. Blennow, An examination of a novel multipanel of CSF biomarkers in the Alzheimer’s disease clinical and pathological continuum. Alzheimers Dement. 17, 431–445 (2021).

83. L. Komsta, F. Novomestky, moments: Moments, cumulants, skewness, kurtosis and related tests (2015; https://CRAN.R-project.org/package=moments).

84. MATLAB (The MathWorks Inc., Natick, MA).

85. J. Hilden, The Area under the ROC Curve and Its Competitors. Med. Decis. Making 11, 95–101 (1991).

86. B. F. Darst, R. L. Koscik, A. M. Racine, J. M. Oh, R. A. Krause, C. M. Carlsson, H. Zetterberg, K. Blennow, B. T. Christian, B. B. Bendlin, O. C. Okonkwo, K. J. Hogan, B. P. Hermann, M. A. Sager, S. Asthana, S. C. Johnson, C. D. Engelman, Pathway-Specific Polygenic Risk Scores as Predictors of Amyloid-β Deposition and Cognitive Function in a Sample at Increased Risk for Alzheimer’s Disease. J. Alzheimers Dis. 55, 473–484 (2017).

87. B. F. Darst, Q. Lu, S. C. Johnson, C. D. Engelman, Integrated analysis of genomics, longitudinal metabolomics, and Alzheimer’s risk factors among 1,111 cohort participants. Genet. Epidemiol. 43, 657– 674 (2019).

88. D. J. Panyard, K. M. Kim, B. F. Darst, Y. K. Deming, X. Zhong, Y. Wu, H. Kang, C. M. Carlsson, S. C. Johnson, S. Asthana, C. D. Engelman, Q. Lu, Cerebrospinal fluid metabolomics identifies 19 brain-related phenotype associations. *Commun*. Biol. 4, 1–11 (2021).

89. S. Das, L. Forer, S. Schönherr, C. Sidore, A. E. Locke, A. Kwong, S. I. Vrieze, E. Y. Chew, S. Levy, M. McGue, D. Schlessinger, D. Stambolian, P.-R. Loh, W. G. Iacono, A. Swaroop, L. J. Scott, F. Cucca, F. Kronenberg, M. Boehnke, G. R. Abecasis, C. Fuchsberger, Next-generation genotype imputation service and methods. Nat. Genet. 48, 1284–1287 (2016).

90. S. McCarthy, S. Das, W. Kretzschmar, O. Delaneau, A. R. Wood, A. Teumer, H. M. Kang, C. Fuchsberger, P. Danecek, K. Sharp, Y. Luo, C. Sidore, A. Kwong, N. Timpson, S. Koskinen, S. Vrieze, L. J. Scott, H. Zhang, A. Mahajan, J. Veldink, U. Peters, C. Pato, C. M. van Duijn, C. E. Gillies, I. Gandin, M. Mezzavilla, A. Gilly, M. Cocca, M. Traglia, A. Angius, J. C. Barrett, D. Boomsma, K. Branham, G. Breen, C. M. Brummett, F. Busonero, H. Campbell, A. Chan, S. Chen, E. Chew, F. S. Collins, L. J. Corbin, G. D. Smith, G. Dedoussis, M. Dorr, A.-E. Farmaki, L. Ferrucci, L. Forer, R. M. Fraser, S. Gabriel, S. Levy, L. Groop, T. Harrison, A. Hattersley, O. L. Holmen, K. Hveem, M. Kretzler, J. C. Lee, M. McGue, T. Meitinger, D. Melzer, J. L. Min, K. L. Mohlke, J. B. Vincent, M. Nauck, D. Nickerson, A. Palotie, M. Pato, N. Pirastu, M. McInnis, J. B. Richards, C. Sala, V. Salomaa, D. Schlessinger, S. Schoenherr, P. E. Slagboom, K. Small, T. Spector, D. Stambolian, M. Tuke, J. Tuomilehto, L. H. Van den Berg, W. Van Rheenen, U. Volker, C. Wijmenga, D. Toniolo, E. Zeggini, P. Gasparini, M. G. Sampson, J. F. Wilson, T. Frayling, P. I. W. de Bakker, M. A. Swertz, S. McCarroll, C. Kooperberg, A. Dekker, D. Altshuler, C. Willer, W. Iacono, S. Ripatti, N. Soranzo, K. Walter, A. Swaroop, F. Cucca, C. A. Anderson, R. M. Myers, M. Boehnke, M. I. McCarthy, R. Durbin, G. Abecasis, J. Marchini, the Haplotype Reference Consortium, A reference panel of 64,976 haplotypes for genotype imputation. Nat. Genet. 48, 1279–1283 (2016).

91. P.-R. Loh, P. Danecek, P. F. Palamara, C. Fuchsberger, Y. A. Reshef, H. K. Finucane, S. Schoenherr, L. Forer, S. McCarthy, G. R. Abecasis, R. Durbin, A. L. Price, Reference-based phasing using the Haplotype Reference Consortium panel. Nat. Genet. 48, 1443–1448 (2016).

92. A. Auton, G. R. Abecasis, D. M. Altshuler, R. M. Durbin, G. R. Abecasis, D. R. Bentley, A. Chakravarti, A. G. Clark, P. Donnelly, E. E. Eichler, P. Flicek, S. B. Gabriel, R. A. Gibbs, E. D. Green, M. E. Hurles, B. M. Knoppers, J. O. Korbel, E. S. Lander, C. Lee, H. Lehrach, E. R. Mardis, G. T. Marth, G. A. McVean, D. A. Nickerson, J. P. Schmidt, S. T. Sherry, J. Wang, R. K. Wilson, R. A. Gibbs, E. Boerwinkle, H. Doddapaneni, Y. Han, V. Korchina, C. Kovar, S. Lee, D. Muzny, J. G. Reid, Y. Zhu, J. Wang, Y. Chang, Q. Feng, X. Fang, X. Guo, M. Jian, H. Jiang, X. Jin, T. Lan, G. Li, J. Li, Y. Li, S. Liu, X. Liu, Y. Lu, X. Ma, M. Tang, B. Wang, G. Wang, H. Wu, R. Wu, X. Xu, Y. Yin, D. Zhang, W. Zhang, J. Zhao, M. Zhao, X. Zheng, E. S. Lander, D. M. Altshuler, S. B. Gabriel, N. Gupta, N. Gharani, L. H. Toji, N. P. Gerry, A. M. Resch, P. Flicek, J. Barker, L. Clarke, L. Gil, S. E. Hunt, G. Kelman, E. Kulesha, R. Leinonen, W. M. McLaren, R. Radhakrishnan, A. Roa, D. Smirnov, R. E. Smith, I. Streeter, A. Thormann, I. Toneva, B. Vaughan, X. Zheng-Bradley, D. R. Bentley, R. Grocock, S. Humphray, T. James, Z. Kingsbury, H. Lehrach, R. Sudbrak, M. W. Albrecht, V. S. Amstislavskiy, T. A. Borodina, M. Lienhard, F. Mertes, M. Sultan, B. Timmermann, M.-L. Yaspo, E. R. Mardis, R. K. Wilson, L. Fulton, R. Fulton, S. T. Sherry, V. Ananiev, Z. Belaia, D. Beloslyudtsev, N. Bouk, C. Chen, D. Church, R. Cohen, C. Cook, J. Garner, T. Hefferon, M. Kimelman, C. Liu, J. Lopez, P. Meric, C. O’Sullivan, Y. Ostapchuk, L. Phan, S. Ponomarov, V. Schneider, E. Shekhtman, K. Sirotkin, D. Slotta, H. Zhang, G. A. McVean, R. M. Durbin, S. Balasubramaniam, J. Burton, P. Danecek, T. M. Keane, A. Kolb-Kokocinski, S. McCarthy, J. Stalker, M. Quail, J. P. Schmidt, C. J. Davies, J. Gollub, T. Webster, B. Wong, Y. Zhan, A. Auton, C. L. Campbell, Y. Kong, A. Marcketta, R. A. Gibbs, F. Yu, L. Antunes, M. Bainbridge, D. Muzny, A. Sabo, Z. Huang, J. Wang, L. J. M. Coin, L. Fang, X. Guo, X. Jin, G. Li, Q. Li, Y. Li, Z. Li, H. Lin, B. Liu, R. Luo, H. Shao, Y. Xie, C. Ye, C. Yu, F. Zhang, H. Zheng, H. Zhu, C. Alkan, E. Dal, F. Kahveci, G. T. Marth, E. P. Garrison, D. Kural, W.-P. Lee, W. Fung Leong, M. Stromberg, A. N. Ward, J. Wu, M. Zhang, M. J. Daly, M. A. DePristo, R. E. Handsaker, D. M. Altshuler, E. Banks, G. Bhatia, G. del Angel, S. B. Gabriel, G. Genovese, N. Gupta, H. Li, S. Kashin, E. S. Lander, S. A. McCarroll, J. C. Nemesh, R. E. Poplin, S. C. Yoon, J. Lihm, V. Makarov, A. G. Clark, S. Gottipati, A. Keinan, J. L. Rodriguez-Flores, J. O. Korbel, T. Rausch, M. H. Fritz, A. M. Stütz, P. Flicek, K. Beal, L. Clarke, A. Datta, J. Herrero, W. M. McLaren, G. R. S. Ritchie, R. E. Smith, D. Zerbino, X. Zheng-Bradley, P. C. Sabeti, I. Shlyakhter, S. F. Schaffner, J. Vitti, D. N. Cooper, E. V. Ball, P. D. Stenson, D. R. Bentley, B. Barnes, M. Bauer, R. Keira Cheetham, A. Cox, M. Eberle, S. Humphray, S. Kahn, L. Murray, J. Peden, R. Shaw, E. E. Kenny, M. A. Batzer, M. K. Konkel, J. A. Walker, D. G. MacArthur, M. Lek, R. Sudbrak, V. S. Amstislavskiy, R. Herwig, E. R. Mardis, L. Ding, D. C. Koboldt, D. Larson, K. Ye, S. Gravel, The 1000 Genomes Project Consortium, Corresponding authors, Steering committee, Production group, Baylor College of Medicine, BGI-Shenzhen, Broad Institute of MIT and Harvard, Coriell Institute for Medical Research, E. B. I. European Molecular Biology Laboratory, Illumina, Max Planck Institute for Molecular Genetics, McDonnell Genome Institute at Washington University, US National Institutes of Health, University of Oxford, Wellcome Trust Sanger Institute, Analysis group, Affymetrix, Albert Einstein College of Medicine, Bilkent University, Boston College, Cold Spring Harbor Laboratory, Cornell University, European Molecular Biology Laboratory, Harvard University, Human Gene Mutation Database, Icahn School of Medicine at Mount Sinai, Louisiana State University, Massachusetts General Hospital, McGill University, N. National Eye Institute, A global reference for human genetic variation. Nature 526, 68–74 (2015).

93. S. Purcell, B. Neale, K. Todd-Brown, L. Thomas, M. A. R. Ferreira, D. Bender, J. Maller, P. Sklar, P. I. W. de Bakker, M. J. Daly, P. C. Sham, PLINK: a tool set for whole-genome association and population-based linkage analyses. Am. J. Hum. Genet. 81, 559–575 (2007).

94. S. Champely, pwr: Basic Functions for Power Analysis (2020; https://CRAN.R-project.org/package=pwr).

95. E. Shishkova, A. S. Hebert, M. S. Westphall, J. J. Coon, Ultra-High Pressure (>30,000 psi) Packing of Capillary Columns Enhancing Depth of Shotgun Proteomic Analyses. Anal. Chem. 90, 11503–11508 (2018).

96. J. Cox, M. Mann, MaxQuant enables high peptide identification rates, individualized p.p.b.-range mass accuracies and proteome-wide protein quantification. Nat. Biotechnol. 26, 1367–1372 (2008).

97. S. Tyanova, T. Temu, J. Cox, The MaxQuant computational platform for mass spectrometry-based shotgun proteomics. Nat. Protoc. 11, 2301–2319 (2016).

98. M. Carlson, org.Hs.eg.db: Genome wide annotation for Human (2020).

99. The UniProt Consortium, UniProt: a worldwide hub of protein knowledge. Nucleic Acids Res. 47, D506–D515 (2019).

100. A. Frankish, M. Diekhans, A.-M. Ferreira, R. Johnson, I. Jungreis, J. Loveland, J. M. Mudge, C. Sisu, J. Wright, J. Armstrong, I. Barnes, A. Berry, A. Bignell, S. Carbonell Sala, J. Chrast, F. Cunningham, T. Di Domenico, S. Donaldson, I. T. Fiddes, C. García Girón, J. M. Gonzalez, T. Grego, M. Hardy, T. Hourlier, T. Hunt, O. G. Izuogu, J. Lagarde, F. J. Martin, L. Martínez, S. Mohanan, P. Muir, F. C. P. Navarro, A. Parker, Pei, F. Pozo, M. Ruffier, B. M. Schmitt, E. Stapleton, M.-M. Suner, I. Sycheva, B. Uszczynska-Ratajczak, J. Xu, A. Yates, D. Zerbino, Y. Zhang, B. Aken, J. S. Choudhary, M. Gerstein, R. Guigó, T. J. P. Hubbard, M. Kellis, B. Paten, A. Reymond, M. L. Tress, P. Flicek, GENCODE reference annotation for the human and mouse genomes. Nucleic Acids Res. 47, D766–D773 (2019).

101. S. Tweedie, B. Braschi, K. Gray, T. E. M. Jones, R. L. Seal, B. Yates, E. A. Bruford, Genenames.org: the HGNC and VGNC resources in 2021. Nucleic Acids Res. 49, D939–D946 (2021).

102. Z. Gu, R. Eils, M. Schlesner, Complex heatmaps reveal patterns and correlations in multidimensional genomic data. Bioinformatics 32, 2847–2849 (2016).

103. A. Kassambara, F. Mundt, factoextra: Extract and Visualize the Results of Multivariate Data Analyses (2020; https://CRAN.R-project.org/package=factoextra).

104. The Gene Ontology Consortium, The Gene Ontology resource: enriching a GOld mine. Nucleic Acids Res. 49, D325–D334 (2020).

105. M. Kanehisa, S. Goto, KEGG: Kyoto Encyclopedia of Genes and Genomes. Nucleic Acids Res. 28, 27 (2000).

106. L. M. Schriml, E. Mitraka, J. Munro, B. Tauber, M. Schor, L. Nickle, V. Felix, L. Jeng, C. Bearer, R. Lichenstein, K. Bisordi, N. Campion, B. Hyman, D. Kurland, C. P. Oates, S. Kibbey, P. Sreekumar, C. Le, M. Giglio, C. Greene, Human Disease Ontology 2018 update: classification, content and workflow expansion. Nucleic Acids Res. 47, D955–D962 (2019).

107. G. Yu, L.-G. Wang, Y. Han, Q.-Y. He, clusterProfiler: an R package for comparing biological themes among gene clusters. Omics J. Integr. Biol. 16, 284–287 (2012).

108. G. Yu, L.-G. Wang, G.-R. Yan, Q.-Y. He, DOSE: an R/Bioconductor package for disease ontology semantic and enrichment analysis. Bioinformatics 31, 608–609 (2015).

109. G. Yu, Q.-Y. He, ReactomePA: an R/Bioconductor package for reactome pathway analysis and visualization. Mol. Biosyst. 12, 477–479 (2016).

110. L. Scrucca, M. Fop, T. B. Murphy, A. E. Raftery, mclust 5: Clustering, Classification and Density Estimation Using Gaussian Finite Mixture Models. R J. 8, 289–317 (2016).

111. C.-H. Gao, ggVennDiagram: A “ggplot2” Implement of Venn Diagram (2019).

112. T. L. Pedersen, tidygraph: A Tidy API for Graph Manipulation (; https://CRAN.R-project.org/package=tidygraph).

113. A. Clauset, M. E. J. Newman, C. Moore, Finding community structure in very large networks. *Phys*. Rev. E 70, 066111 (2004).

114. G. Csardi, T. Nepusz, The igraph software package for complex network research (2006; http://igraph.org).

115. N. Gehlenborg, UpSetR: A More Scalable Alternative to Venn and Euler Diagrams for Visualizing Intersecting Sets (2019; https://CRAN.R-project.org/package=UpSetR).

116. J. C. Morris, The Clinical Dementia Rating (CDR): current version and scoring rules. Neurology 43, 2412–2414 (1993).

117. W. T. Hu, D. M. Holtzman, A. M. Fagan, L. M. Shaw, R. Perrin, S. E. Arnold, M. Grossman, C. Xiong, R. Craig-Schapiro, C. M. Clark, E. Pickering, M. Kuhn, Y. Chen, V. M. V. Deerlin, L. McCluskey, L. Elman, J. Karlawish, A. Chen-Plotkin, H. I. Hurtig, A. Siderowf, F. Swenson, V. M.-Y. Lee, J. C. Morris, J. Q. Trojanowski, H. Soares, Plasma multianalyte profiling in mild cognitive impairment and Alzheimer disease. Neurology 79, 897–905 (2012).

118. A. M. Fagan, M. A. Mintun, R. H. Mach, S.-Y. Lee, C. S. Dence, A. R. Shah, G. N. LaRossa, M. L. Spinner, W. E. Klunk, C. A. Mathis, S. T. DeKosky, J. C. Morris, D. M. Holtzman, Inverse relation between in vivo amyloid imaging load and cerebrospinal fluid Aβ42 in humans. Ann. Neurol. 59, 512–519 (2006).

119. Y. Deming, F. Filipello, F. Cignarella, C. Cantoni, S. Hsu, R. Mikesell, Z. Li, J. L. Del-Aguila, U. Dube, F. G. Farias, J. Bradley, J. Budde, L. Ibanez, M. V. Fernandez, K. Blennow, H. Zetterberg, A. Heslegrave, P. M. Johansson, J. Svensson, B. Nellgård, A. Lleo, D. Alcolea, J. Clarimon, L. Rami, J. L. Molinuevo, M. Suárez-Calvet, E. Morenas-Rodríguez, G. Kleinberger, M. Ewers, O. Harari, C. Haass, T. J. Brett, B. A. Benitez, C. M. Karch, L. Piccio, C. Cruchaga, The MS4A gene cluster is a key modulator of soluble TREM2 and Alzheimer’s disease risk. Sci. Transl. Med. 11 (2019), doi:10.1126/scitranslmed.aau2291.

120. L. Gold, D. Ayers, J. Bertino, C. Bock, A. Bock, E. N. Brody, J. Carter, A. B. Dalby, B. E. Eaton, T. Fitzwater, D. Flather, A. Forbes, T. Foreman, C. Fowler, B. Gawande, M. Goss, M. Gunn, S. Gupta, D. Halladay, J. Heil, J. Heilig, B. Hicke, G. Husar, N. Janjic, T. Jarvis, S. Jennings, E. Katilius, T. R. Keeney, N. Kim, T. H. Koch, S. Kraemer, L. Kroiss, N. Le, D. Levine, W. Lindsey, B. Lollo, W. Mayfield, M. Mehan, R. Mehler, S. K. Nelson, M. Nelson, D. Nieuwlandt, M. Nikrad, U. Ochsner, R. M. Ostroff, M. Otis, T. Parker, S. Pietrasiewicz, D. I. Resnicow, J. Rohloff, G. Sanders, S. Sattin, D. Schneider, B. Singer, M. Stanton, A. Sterkel, A. Stewart, S. Stratford, J. D. Vaught, M. Vrkljan, J. J. Walker, M. Watrobka, S. Waugh, A. Weiss, S. K. Wilcox, A. Wolfson, S. K. Wolk, C. Zhang, D. Zichi, Aptamer-Based Multiplexed Proteomic Technology for Biomarker Discovery. PLOS ONE 5, e15004 (2010).

121. C. Yang, F. H. G. Farias, L. Ibanez, A. Suhy, B. Sadler, M. V. Fernandez, F. Wang, J. L. Bradley, B. Eiffert, J. A. Bahena, J. P. Budde, Z. Li, U. Dube, Y. J. Sung, K. A. Mihindukulasuriya, J. C. Morris, A. M. Fagan, R. J. Perrin, B. A. Benitez, H. Rhinn, O. Harari, C. Cruchaga, Genomic atlas of the proteome from brain, CSF and plasma prioritizes proteins implicated in neurological disorders. Nat. Neurosci., 1–11 (2021).

122. J. T. Leek, W. E. Johnson, H. S. Parker, A. E. Jaffe, J. D. Storey, The sva package for removing batch effects and other unwanted variation in high-throughput experiments. Bioinformatics 28, 882–883 (2012).

123. H. Zou, T. Hastie, Regularization and variable selection via the elastic net. J. R. Stat. Soc. Ser. B Stat. Methodol. 67, 301–320 (2005).

124. J. Friedman, T. Hastie, R. Tibshirani, Regularization Paths for Generalized Linear Models via Coordinate Descent. J. Stat. Softw. 33, 1–22 (2010).

125. M. Kuhn, H. Wickham, Tidymodels: a collection of packages for modeling and machine learning using tidyverse principles (2020; https://www.tidymodels.org).

