## Supplementary Figures for "Large-scale proteome and metabolome analysis of CSF implicates altered glucose metabolism and succinylcarnitine in Alzheimer’s disease"

### Table of Contents

Supplementary Figure 1. Data and analytical overview ... 2
Supplementary Figure 2. Pilot study power analysis ... 3
Supplementary Figure 3. Protein missingness overall ... 4
Supplementary Figure 4. Protein missingness by AT category ... 5
Supplementary Figure 5. Protein imputation examples ... 6
Supplementary Figure 6. CSF proteomics PCA scree plot ... 7
Supplementary Figure 7. CSF proteomics PC plots ... 8
Supplementary Figure 8. Clustering BIC values ... 9
Supplementary Figure 9. Overlap of ANCOVA and *APOE*-controlled results ... 10
Supplementary Figure 10. Overlap of ANCOVA and logistic regression results ... 11
Supplementary Figure 11. Significantly associated proteins with A+T+ vs A-T- ... 12
Supplementary Figure 12. Protein-biomarker association Q-Q plot ... 13
Supplementary Figure 13. Overlap in the proteins associated among the biomarkers ... 14
Supplementary Figure 14. Distribution of IGF-1 by AT category ... 15
Supplementary Figure 15. Relationships between IGF-1 and CSF biomarkers ... 16
Supplementary Figure 16. Distribution of succinylcarnitine (C4-DC) by AT category ... 17

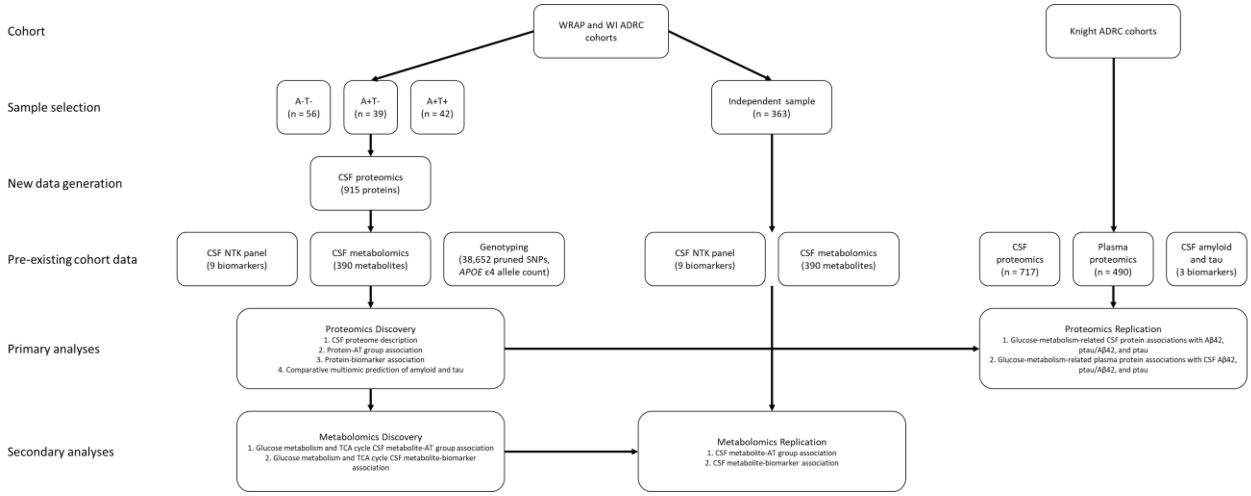


**Supplementary Figure 1. Data and analytical overview**

A schematic representing the primary cohorts, new and previously existing data sets, and the discovery/replication analyses is shown. Note that the previously existing CSF NTK biomarker, metabolomic, and genotyping data in the WRAP and WI ADRC cohorts were generated for the same participants and (for the biomarker and metabolomic data) the same CSF samples as those used to generate the new CSF proteomics data set. Of the 137 WRAP and WI ADRC participants used in the proteomics discovery analyses, one participant lacked CSF metabolomics data and was excluded from the comparative multiomic prediction metabolomics discovery analyses, which consequently only had a sample size of 136.

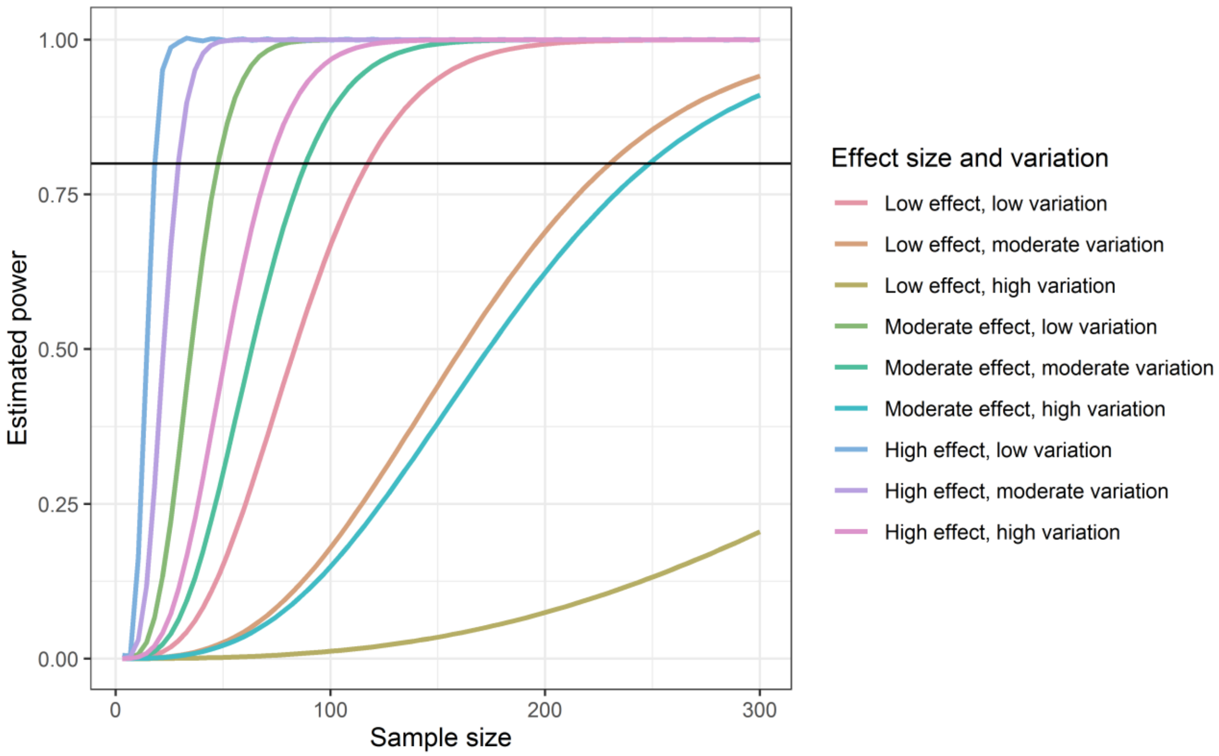


**Supplementary Figure 2. Pilot study power analysis**

The estimated power at various sample sizes for a case-control analysis based on the results of the previously published pilot analysis is shown. Each curve represents a different combination of the effect size (difference in protein levels) and protein variation, based on the observed effect sizes and variances in the pilot study. The horizontal black line represents an estimated power of 0.8.

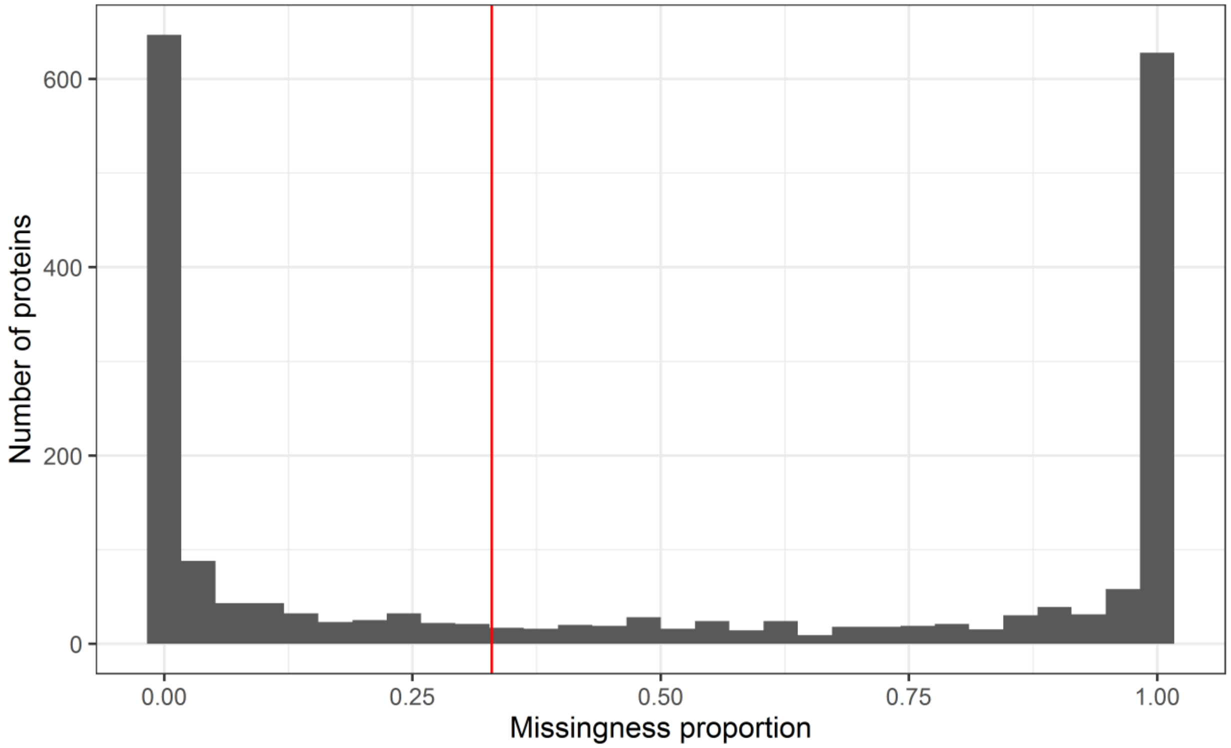


**Supplementary Figure 3. Protein missingness overall**

The distribution of protein missingness for all 2,040 identified proteins across the full data set (n = 137 participants) is shown. The red vertical line indicates the 33% missingness threshold used to exclude highly missing proteins. A strong, bimodal distribution was observed, with most proteins either being fully present or absent.

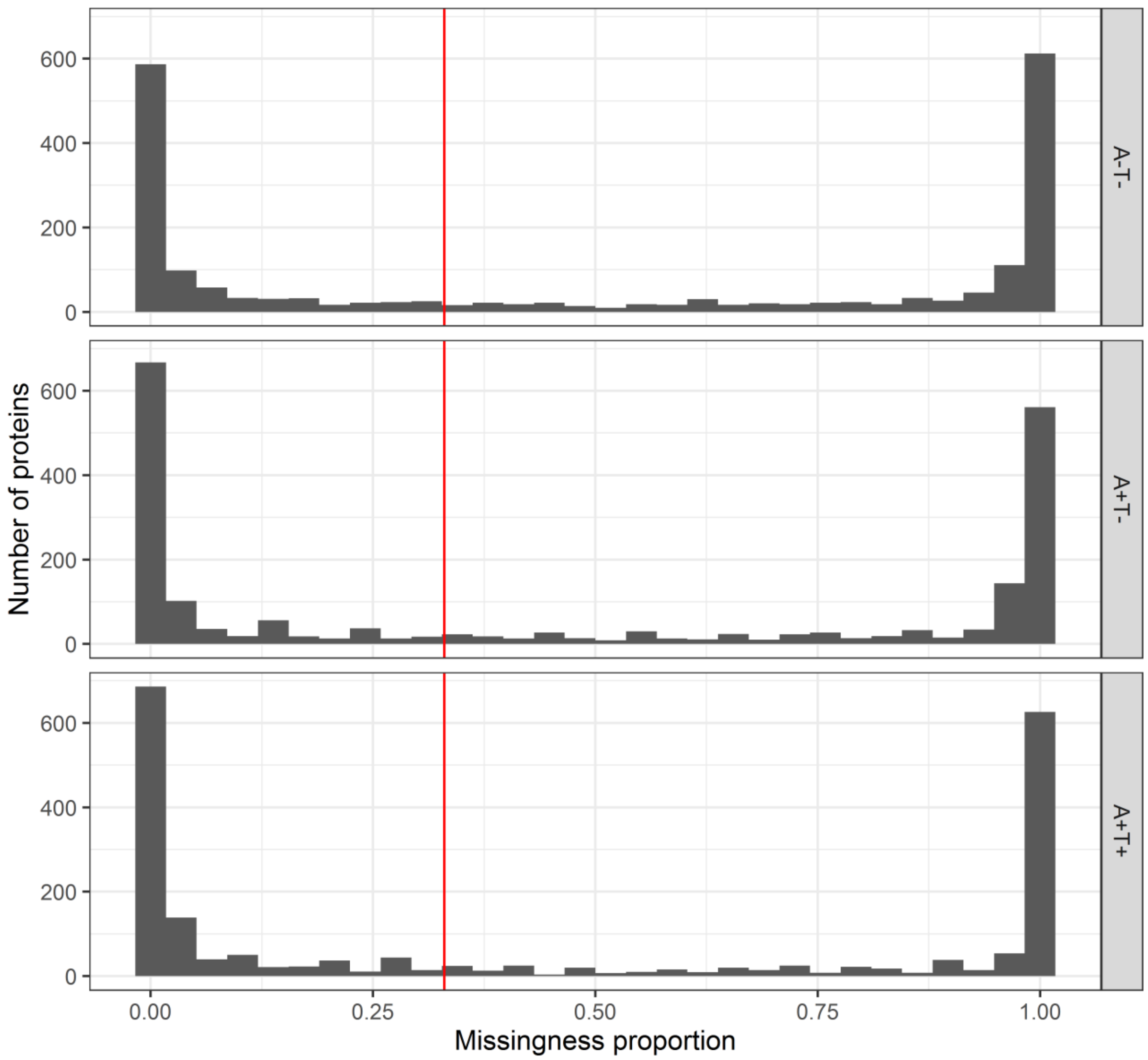


**Supplementary Figure 4. Protein missingness by AT category**

The distribution of protein missingness for all 2,040 identified proteins across the full data set (n = 137 participants), stratified by AT category, is shown. The red vertical line indicates the 33% missingness threshold used to exclude highly missing proteins. A strong, bimodal distribution was observed, with most proteins either being fully present or absent. Little difference was observed between the AT categories or in comparison to the overall missingness distribution.

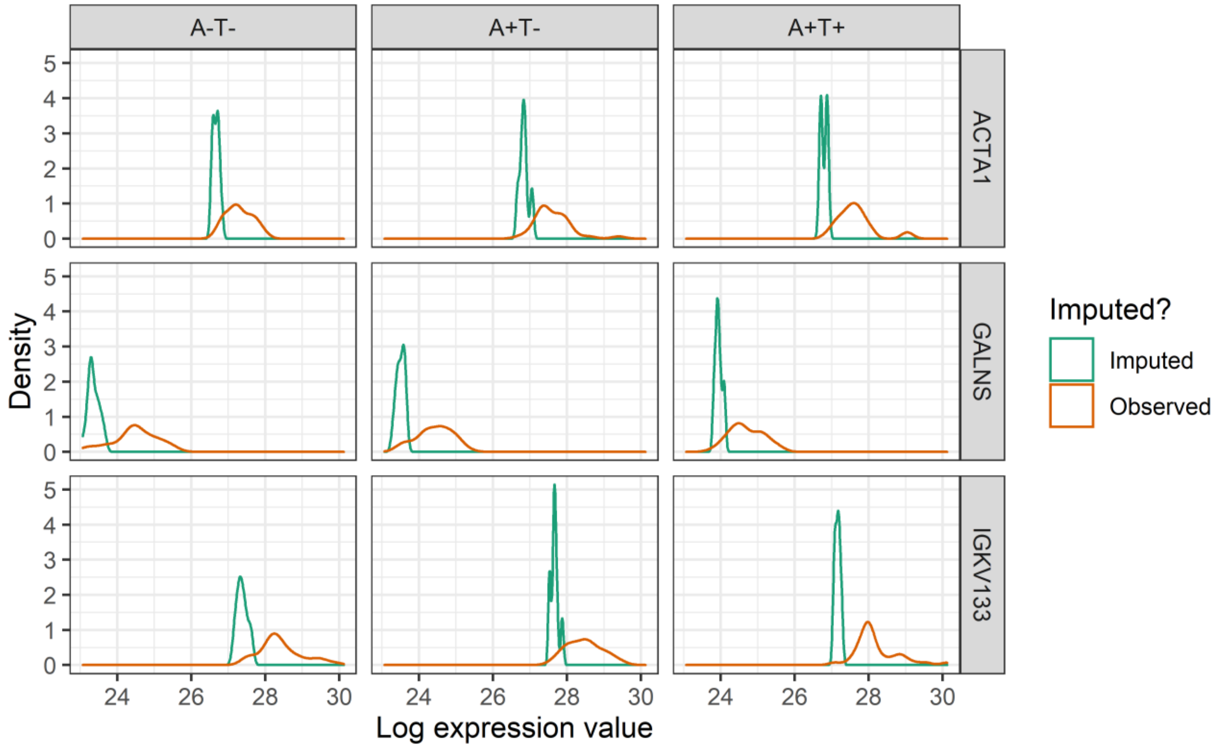


**Supplementary Figure 5. Protein imputation examples**

Examples of the imputation procedure are shown. For the three most-imputed proteins (rows), the observed distribution of values is shown in orange for each AT category (columns). The distribution of imputed values is shown in green.

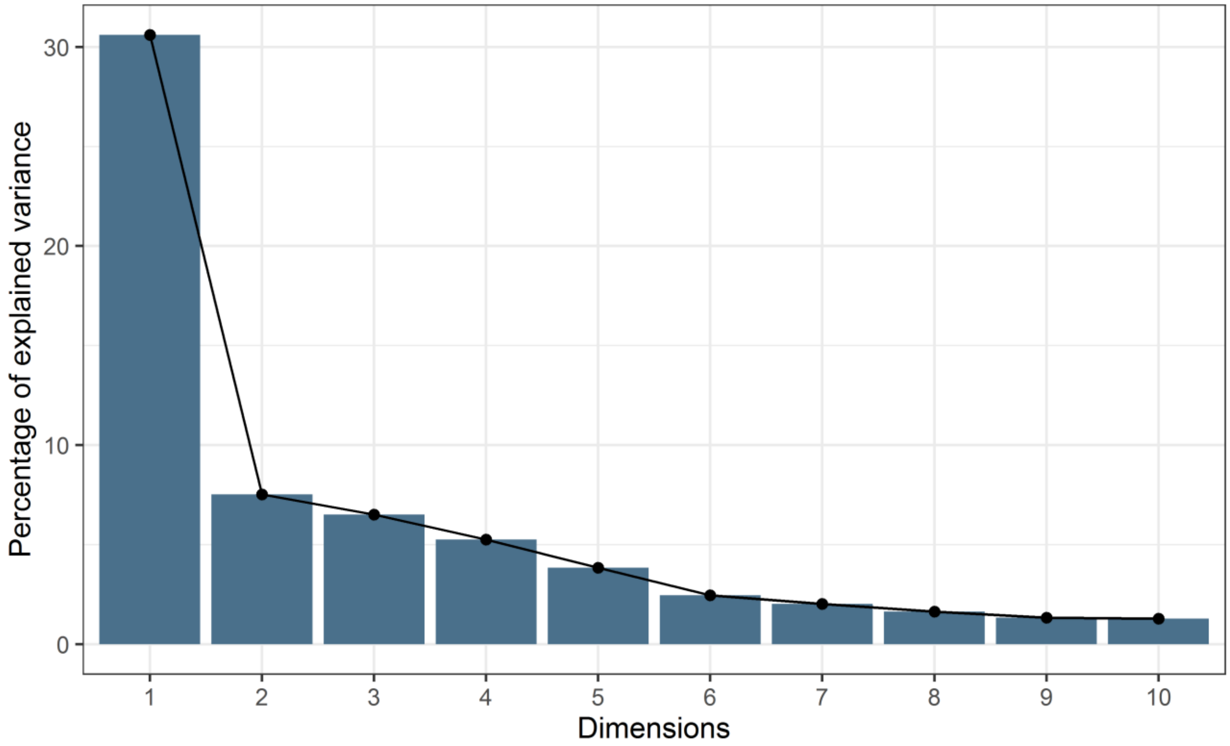


**Supplementary Figure 6. CSF proteomics PCA scree plot**

The scree plot of the first 10 PCs of the CSF proteomics data is shown. The first PC captured substantially more variance (30.6%) than subsequent PCs, but the complexity of the CSF proteome was highlighted by the low collective variance explained by the top PCs. The first 4 PCs only explained a total of 49.89% (30.6%, 7.52%, 6.52%, and 5.26%, respectively) of the total variance.

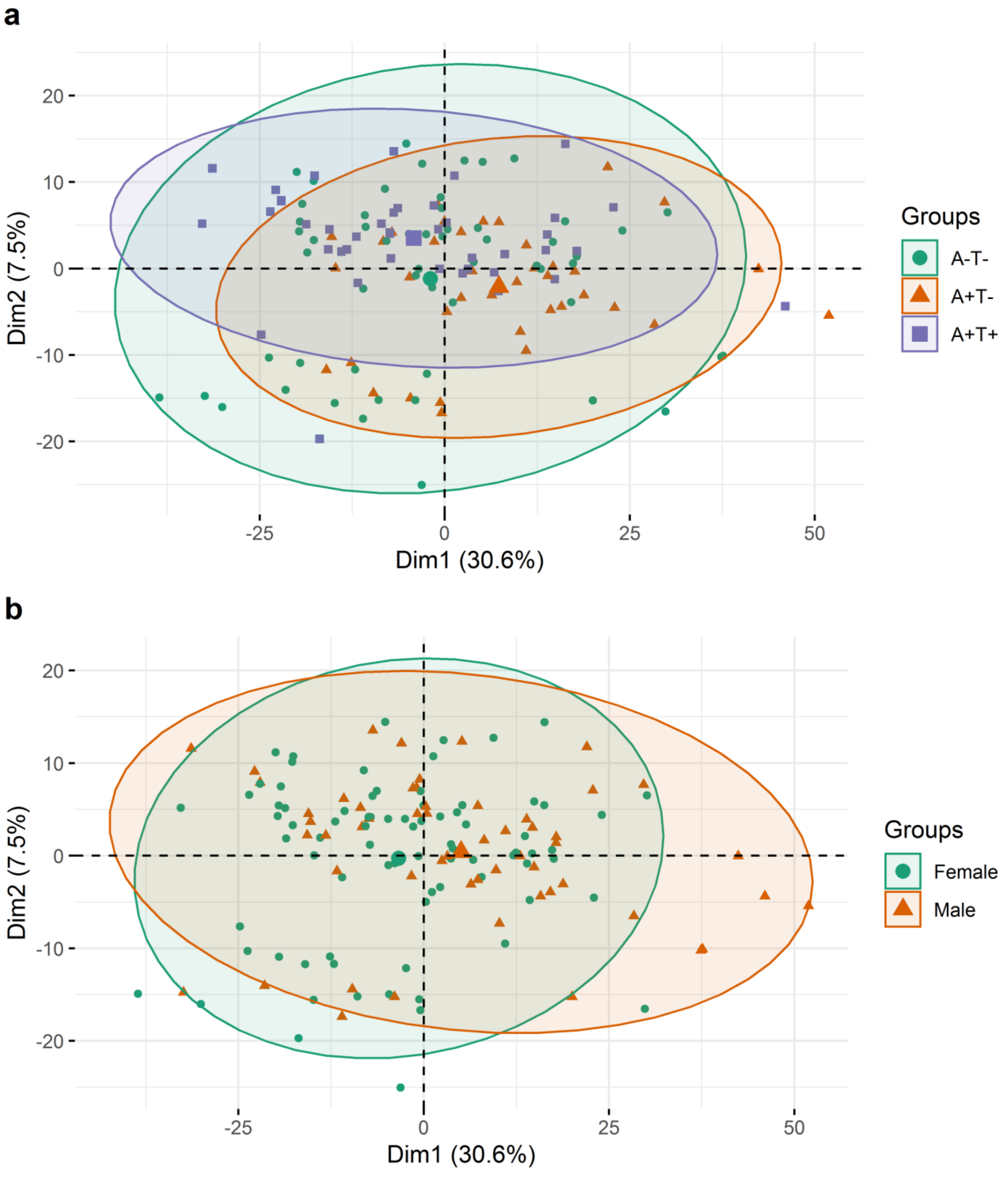


**Supplementary Figure 7. CSF proteomics PC plots**

The plots of the top 2 PCs from the PCA are shown. Little difference across the PCs was seen by either AT category (**a**) or sex (**b**).

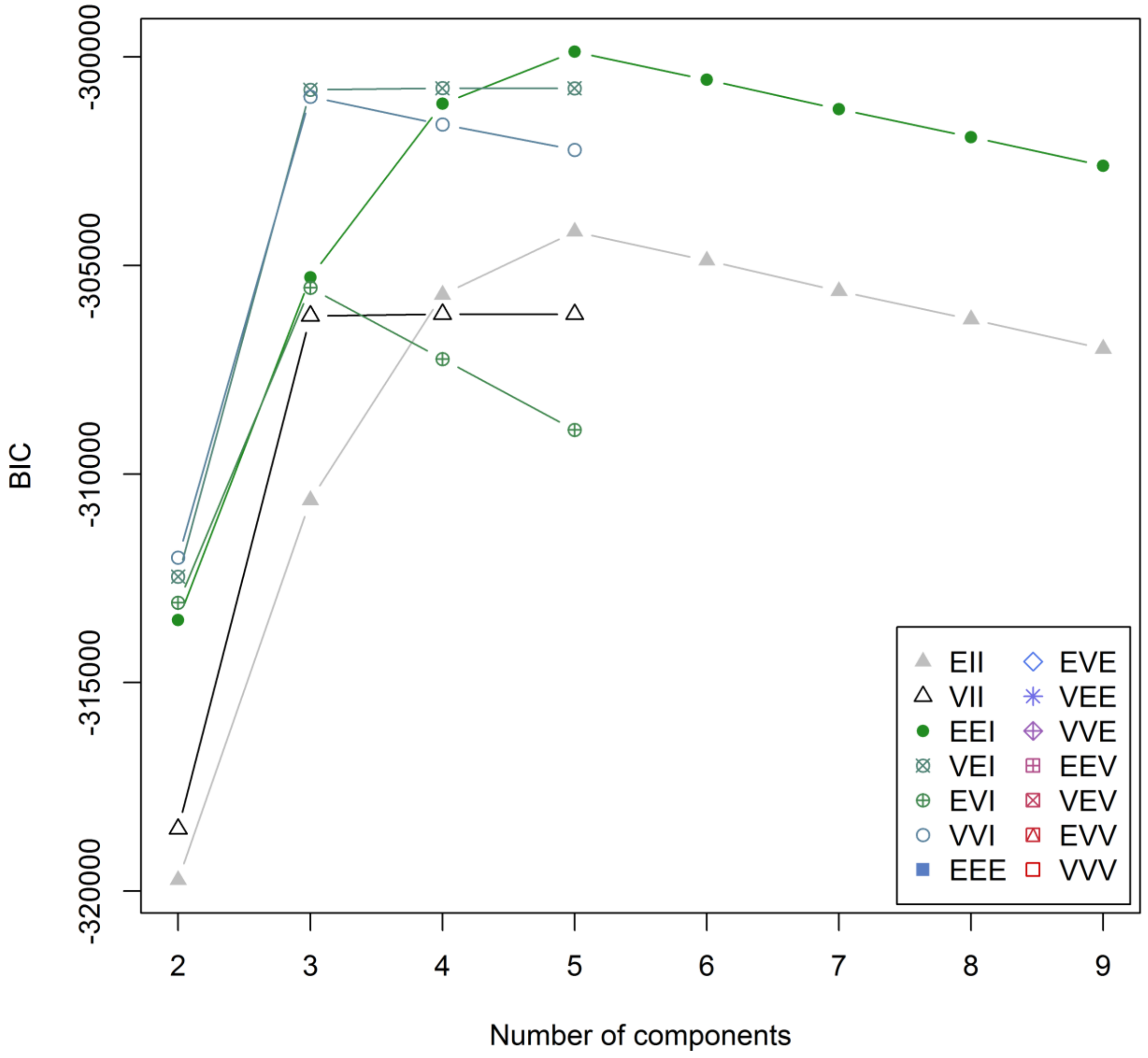


**Supplementary Figure 8. Clustering BIC values**

The Bayesian Information Criterion (BIC) values for different kinds of clustering models and different numbers of clusters (x-axis) are shown. The number of clusters for this analysis (3) was chosen based on the elbow inflection point on the curves.

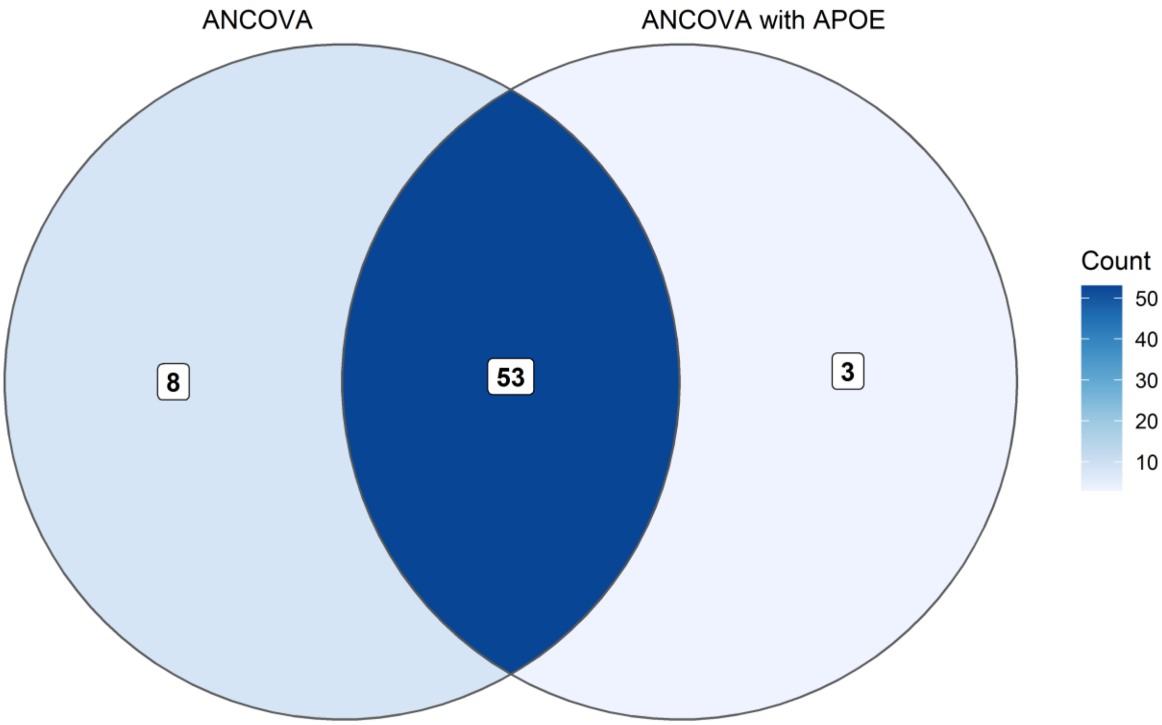


**Supplementary Figure 9. Overlap of ANCOVA and *APOE*-controlled results**

The overlap of the proteins significantly associated with AT category according to the ANCOVA models when the *APOE* ε4 allele count was and was not controlled for is shown above. The majority of the proteins that were significantly associated with AT in the first model remained significant when the *APOE* variable was added, indicating that *APOE* status was unlikely to be confounding the protein-AT associations.

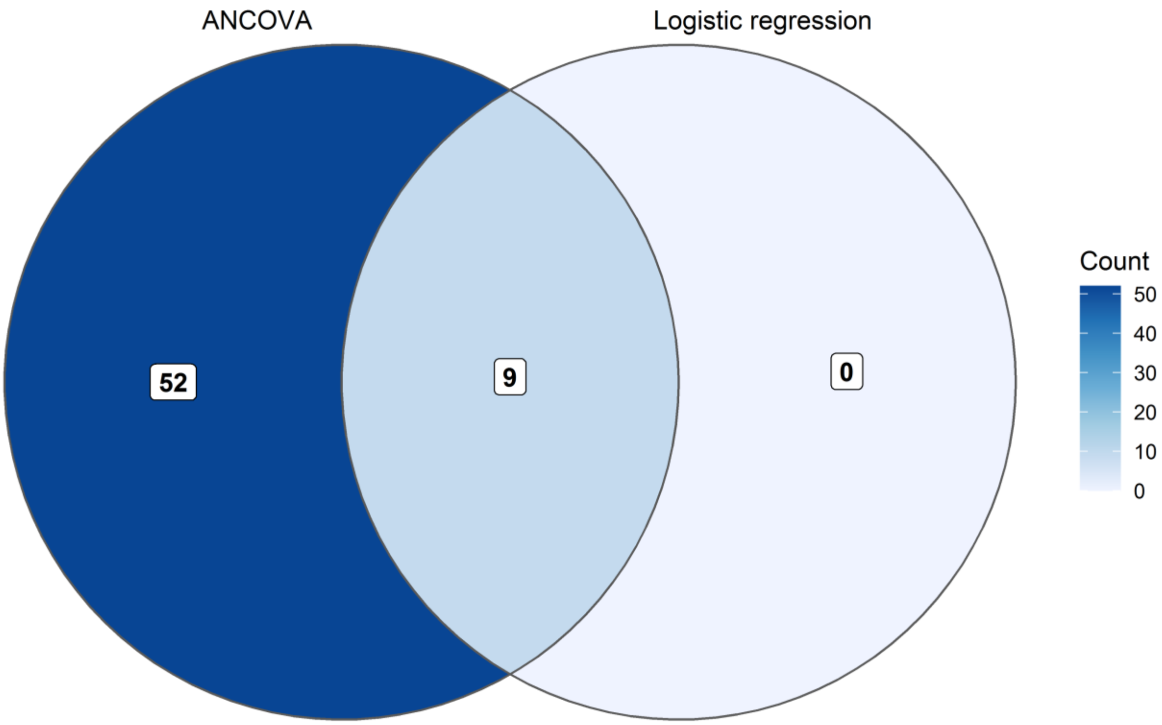


**Supplementary Figure 10. Overlap of ANCOVA and logistic regression results**

The overlap of the proteins significantly associated with AT category according to the ANCOVA models and the proteins associated with A+T+ vs A-T- is shown above. Only 9 proteins remained significant in the logistic regression analysis compared to the ANCOVA, which was likely due to the smaller sample size in the logistic regression models, which excluded the intermediate category of A+T-.

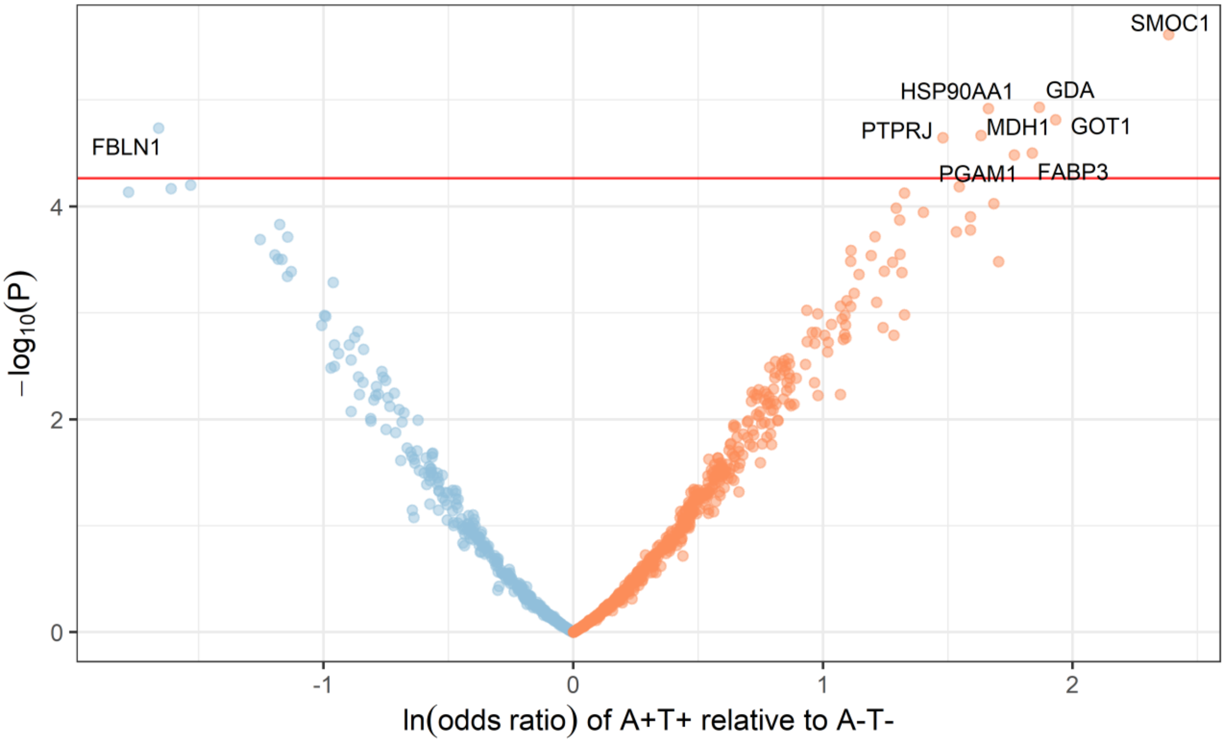


**Supplementary Figure 11. Significantly associated proteins with A+T+ vs A-T-**

A volcano plot of the proteins significantly associated with A+T+ (vs A-T-) in the logistic regression is shown. The horizontal red line shows the Bonferroni-corrected P value threshold of 5.46 x $10^{-5}$. The significantly associated proteins are labeled on the plot. All but 1 of the 9 associated proteins after multiple testing correction were increased (orange) in A+T+ relative to A-T- rather than decreased (blue).

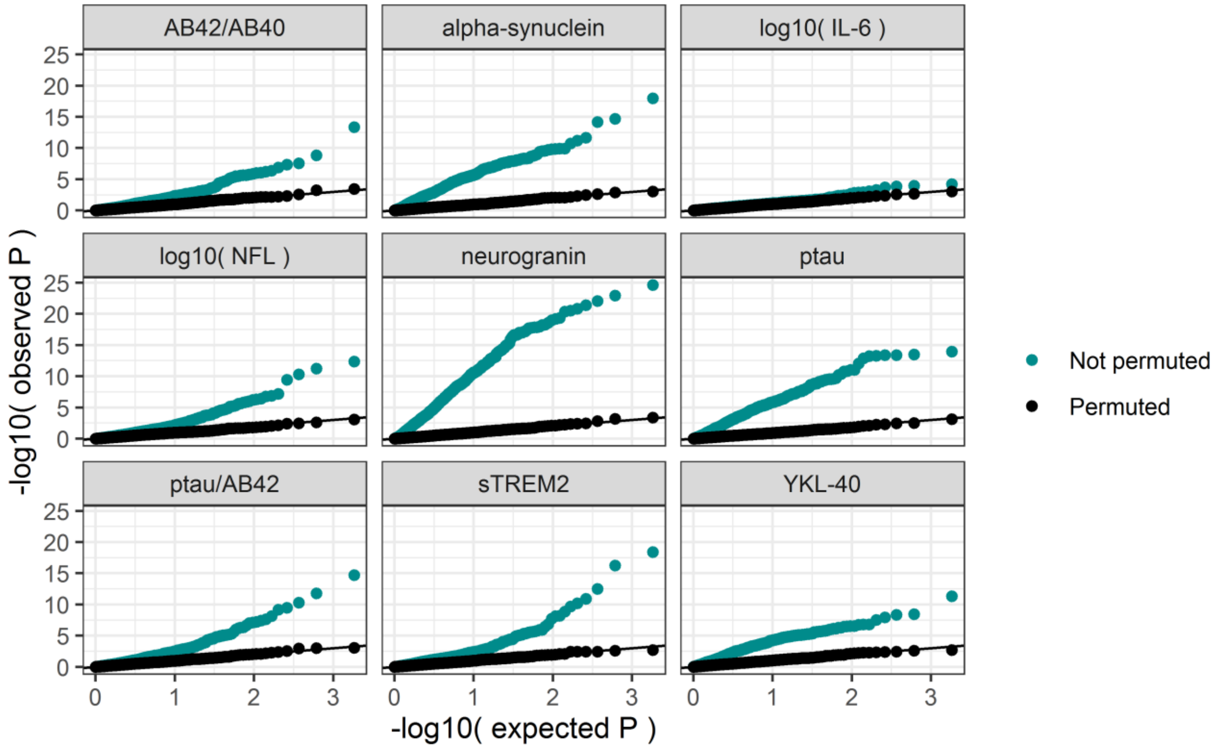


**Supplementary Figure 12. Protein-biomarker association Q-Q plot**

The quantile-quantile (Q-Q) plots of the protein-biomarker linear regression association tests are shown above, with the distribution of P-values shown separately for the original ("Not permuted") and permuted data sets. Substantial signal enrichment was seen across the CSF proteome similar to the protein-AT ANCOVA models, with the exception of IL-6, which showed little association with the CSF proteome. The strongest proteome-wide deviation was seen with neurogranin.

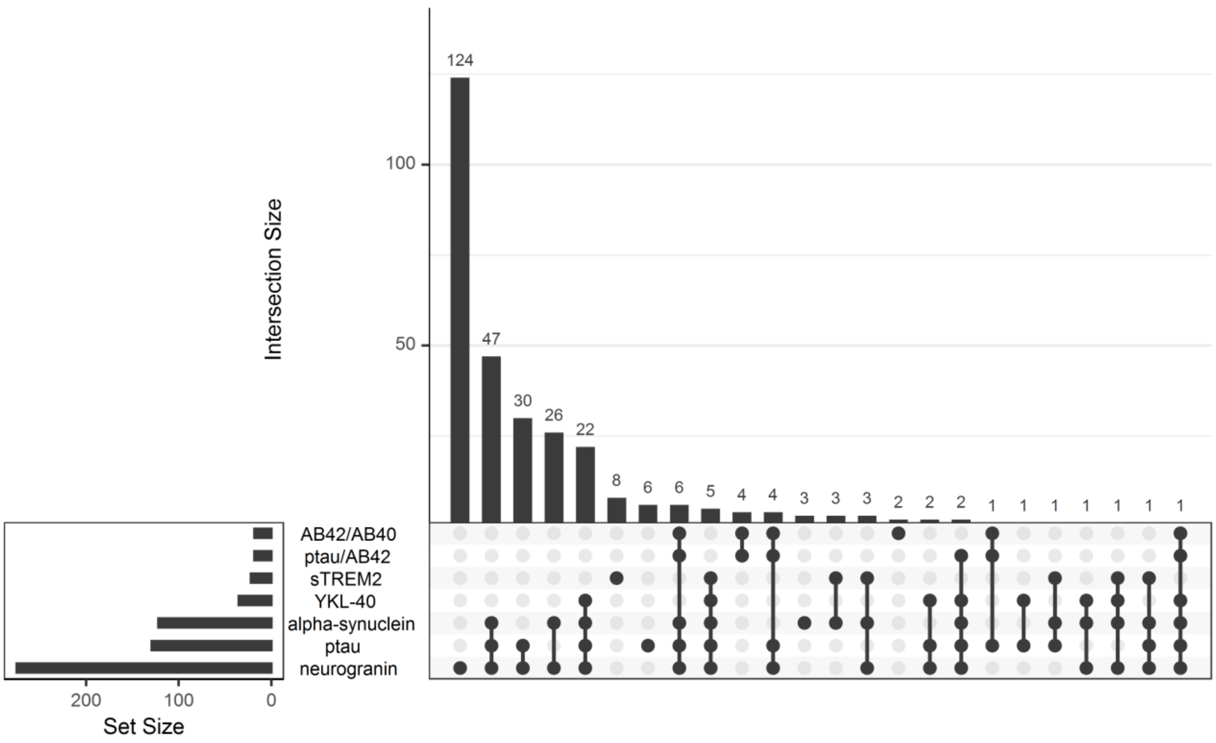


**Supplementary Figure 13. Overlap in the proteins associated among the biomarkers**

An upset plot showing the overlap among the significantly associated proteins for each CSF biomarker is shown for the 7 biomarkers with the greatest number of significant associations. Neurogranin had the greatest number of proteins uniquely associated with it at 124. The largest overlap of associated proteins included neurogranin, ptau, and alpha-synuclein with 103 overlapping proteins.

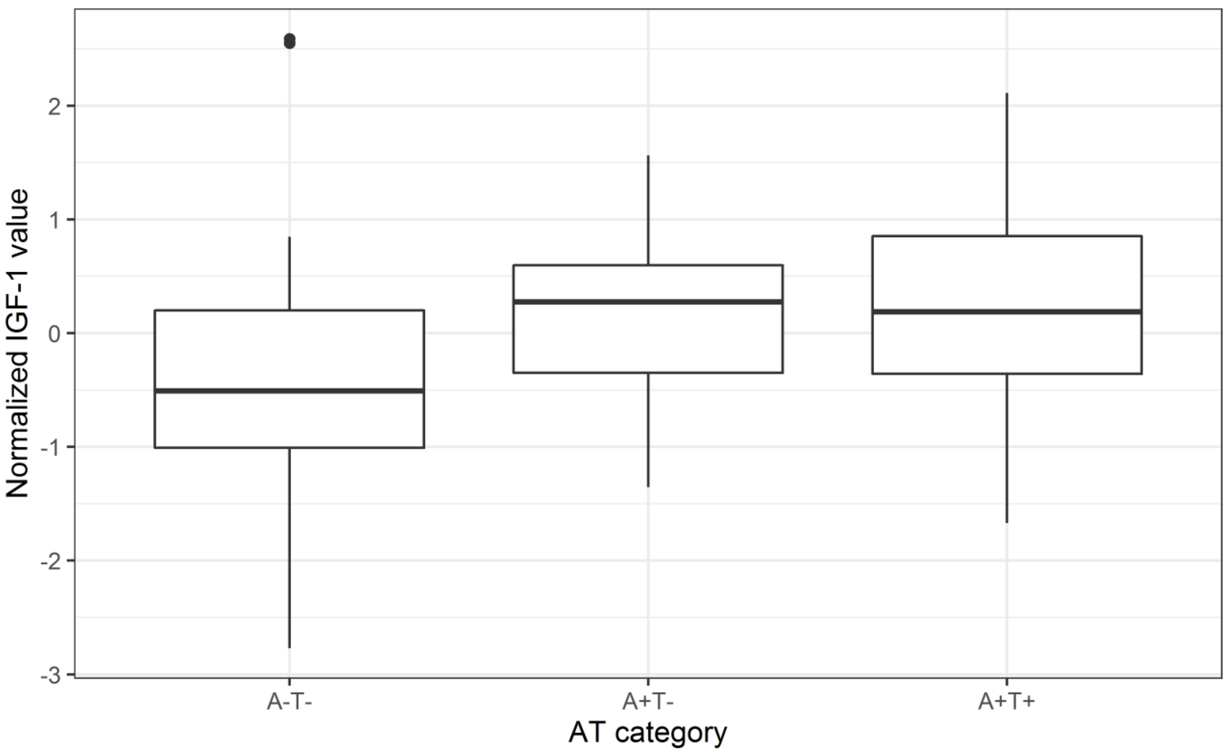


**Supplementary Figure 14. Distribution of IGF-1 by AT category**

A series of box plots show the distribution of IGF-1 across AT category. Due to the higher missingness of IGF-1 in the original proteomics data set, no imputed values for IGF-1 were used. The sample size was roughly similar across categories: A-T- = 31, A+T- = 23, A+T+ = 28, total n = 82. An increase in IGF-1 between the amyloid negative and amyloid positive categories can be seen, with little difference between the A+T- and A+T+ categories.

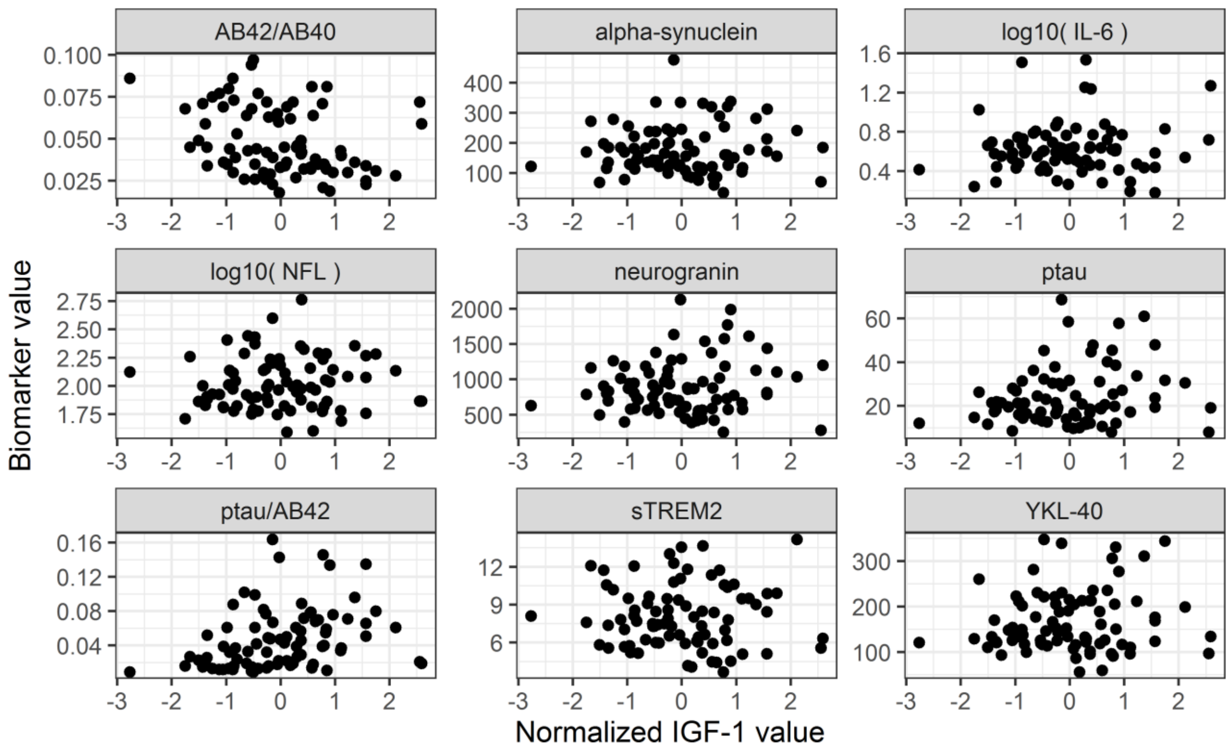


**Supplementary Figure 15. Relationships between IGF-1 and CSF biomarkers**

A set of scatter plots show the relationships between IGF-1 (unimputed, n = 82) and each of the CSF biomarkers is shown. A nominally statistically significant negative relationship with Aβ42/Aβ40 and positive relationship with ptau/Aβ42 was observed.

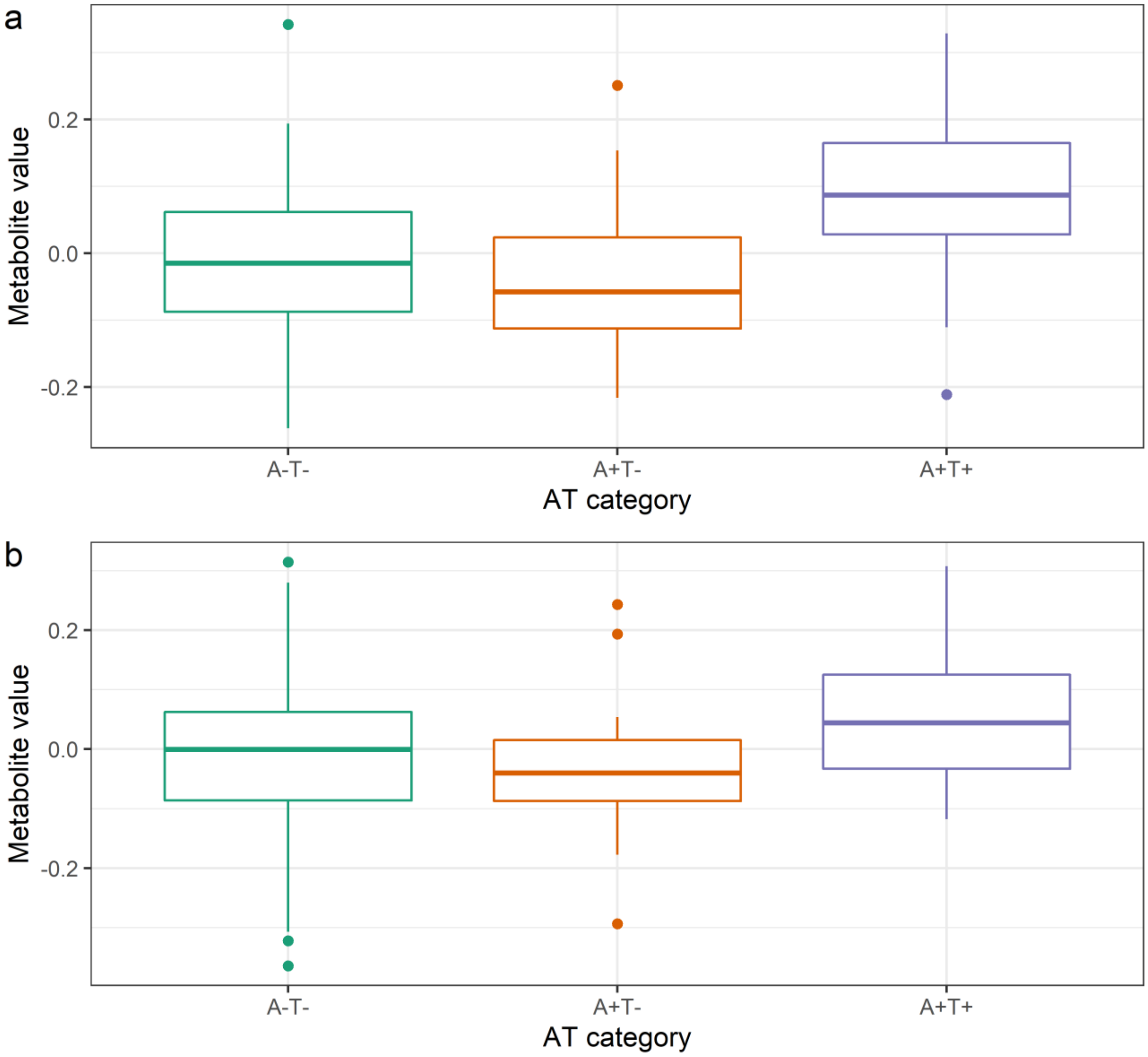


**Supplementary Figure 16. Distribution of succinylcarnitine (C4-DC) by AT category**

Box plots showing the distribution of CSF levels of succinylcarnitine (C4-DC) across AT categories among participants in the proteomics discovery cohort (n = 136) **(a)** and among an independent group of WRAP and WI ADRC participants (n = 363) **(b)**.
